# Personalized Circulating Tumor DNA (ctDNA) Profiling Enables Superior and Universal Measurable Residual Disease (MRD) Detection in Acute Myeloid Leukemia (AML)

**DOI:** 10.64898/2026.01.28.26344873

**Authors:** Ruwan Gunaratne, Crystal Zhou, Sanjeeth Rajaram, Jesse W. Tai, Kailee Tanaka, Charu Tiwari, Emily Yang, Sky Kim, Grace Gao, Raymond Yin, Mia Carleton, Matthew S. Alkaitis, Matthew Schwede, Brian J. Sworder, Gabriel N. Mannis, Michael S. Khodadoust, Ravindra Majeti, David M. Kurtz, Tian Yi Zhang

**Author notes:** Corresponding Author: Tian Yi Zhang, MD, PhD Stanford University School of Medicine Division of Hematology 269 Campus Drive Room 1155 CCSR Bldg MC 5156 Stanford, CA 94305. These authors contributed equally (Crystal Zhou, Sanjeeth Rajaram).

## Abstract

Relapsed and/or refractory disease remains the leading cause of death in AML, highlighting the need for broadly applicable, high-sensitivity approaches to MRD detection. We developed AML-CAPP-Seq (<u>Ca</u>ncer <u>P</u>ersonalized <u>P</u>rofiling by Deep <u>Seq</u>uencing), a personalized hybrid-capture assay that tracks both canonical AML drivers and patient-specific variants identified by whole-exome sequencing. In 56 patients with longitudinal plasma and matched peripheral blood and bone marrow samples, AML-CAPP-Seq enabled universal MRD assessment and resolution of clonal dynamics using a median of 30.5 variants per patient. Plasma ctDNA outperformed cellular compartments for MRD detection and more strongly predicted relapse-free (HR 17.8, p<0.0001) and overall survival (HR 17.0, p<0.0001) than standard-of-care MRD methods. Among 29 allogeneic transplant recipients, peri-transplant ctDNA-MRD dynamics markedly improved relapse risk stratification (HR 36.0, p=0.0009). Together, these results establish personalized ctDNA profiling as a minimally invasive, highly sensitive, and generalizable platform for enhanced clinical MRD detection and clonal surveillance in AML.

**Significance Statement:** We present a personalized blood test for acute myeloid leukemia that tracks patient-specific circulating tumor DNA, enabling sensitive, universal, noninvasive detection of residual disease. It outperforms standard-of-care marrow and cell-based methods for predicting relapse and survival, including after transplant, reveals clonal dynamics, and supports individualized disease monitoring and risk-adapted treatment.

## Introduction

AML is a genomically heterogenous malignancy defined by serially acquired somatic mutations that impair differentiation and promote clonal proliferation^1–3^. Despite therapeutic advances with most patients achieving clinical remission, the majority succumb to relapsed and/or refractory (R/R) disease, driven by persistence of measurable residual disease (MRD) and emergence of treatment resistance^4,5^.

Standard-of-care (SOC) MRD assessment in AML relies on multiparametric flow cytometry (MFC) or molecular assays targeting AML-specific variants but these methods face major limitations^5–8^. Both approaches require invasive bone marrow (BM) sampling, limiting frequent monitoring and introducing sampling error due to anatomic disease heterogeneity^7^. MFC suffers from limited sensitivity, variability in leukemia-associated immunophenotype, and poor harmonization across institutions^6^. While qPCR or next generation sequencing (NGS) methods offer sensitive detection, these are only applicable to ∼30-40% of AML patients with specific mutations (*FLT3-ITD*, *NPM1c*, or core-binding factors) and cannot capture the genomic diversity needed to interrogate clonal evolution and understand mechanisms of R/R AML^5–7^. Circulating tumor DNA (ctDNA) offers a noninvasive alternative for serial MRD assessment that has been successfully applied in solid and hematologic cancers^9,10^. However, owing to the relative mutational sparsity and genomic heterogeneity of AML^2^, fixed-panel strategies commonly used in ctDNA MRD monitoring offer limited sensitivity and are not generalizable to all AML patients^6,11^.

To address this unmet need for sensitive and broadly applicable quantification of AML tumor burden, we developed AML-CAPP-Seq (<u>Ca</u>ncer <u>P</u>ersonalized <u>P</u>rofiling by Deep <u>Seq</u>uencing), a personalized hybrid capture–based ctDNA assay tailored for the genetic complexity of each AML patient^11,12^. AML-CAPP-Seq integrates whole exome sequencing (WES) of diagnostic tumor samples to identify patient-specific somatically acquired single nucleotide variants (SNVs), combined with a panel of known ‘canonical’ driver mutations in AML. This composite approach enables universal and highly sensitive MRD detection in all patients. We show that ctDNA is a superior biomarker for AML MRD detection and genome characterization (**Figure 1A**) compared to traditional marrow-based methods and demonstrate its clinical utility for relapse detection and survival prediction in patients with and without allogeneic bone marrow transplantation (alloHSCT), donor chimerism surveillance, and extramedullary disease detection. These findings support ctDNA as a novel biomarker for noninvasive personalized disease monitoring in AML.

**Figure 1.**
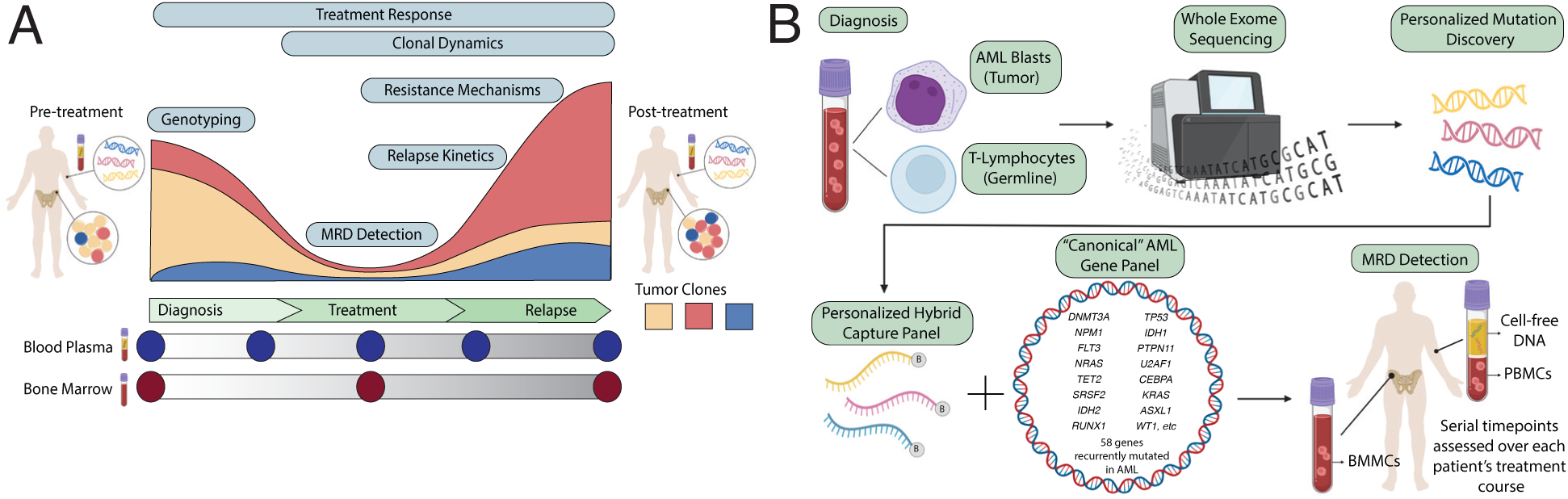
Study design and AML-CAPP-Seq workflow. (A) Overview of clinical applications for ctDNA monitoring in AML, including disease characterization, treatment response assessment, MRD and early relapse detection, as well as evaluation of clonal dynamics, relapse kinetics, and resistance mechanisms. (B) Schematic of the AML-CAPP-Seq assay, which integrates WES of diagnostic samples to identify patient-specific somatic mutations with a fixed 58-gene canonical AML panel to enable personalized hybrid capture–based profiling of serial plasma and cellular samples.

## Results

### AML-CAPP-Seq Design for Personalized MRD Tracking

To enable comprehensive ctDNA-based genomic monitoring in AML, we first designed a canonical 58-gene hybrid capture selector based on recurrently mutated genes identified from established public WES datasets (TCGA^2^ and BeatAML^13^) (**Table S1**). To complement this canonical panel, we also generated personalized hybrid capture selectors derived from patient-specific (‘noncanonical’) variants identified by WES as a composite and customized assay (AML-CAPP-Seq) for MRD detection and disease tracking in each patient (**Figure 1B**). WES was performed on flow-sorted blasts or whole blood from diagnostic samples (**Figure S1**).

### Patient Cohort Overview

We applied AML-CAPP-Seq to 56 AML patients with longitudinal sampling from plasma, BMMCs, and PBMCs (**Table 1**, **Figure 2A, Tables S2-S5**). The cohort included primarily newly diagnosed *de novo* AML (n=42) with a median age of 60.5 years (range 31-85). A median of 5 timepoints (interquartile range, IQR 4–7) over a median follow-up of 25.8 months (IQR 9.8–33.6) were sampled. Clinical NGS assessment at diagnosis revealed a median of 4 genomic alterations per patient (range 1–8), including SNVs, indels, and fusions with the most frequently mutated genes matching established literature (**Figure 2A, Table S4-S5**). All patients were risk stratified by European LeukemiaNet (ELN) 2022 criteria (**Table 1, Tables S2-S5**). Most patients (**73.2%, n=41**) received high-intensity induction therapy, while the remainder received low-intensity or targeted therapies (**Table S2-S3)**. 51.8% (n=29) underwent alloHSCT with transplant conditioning regimens and post-HSCT maintenance therapies detailed in **Table S6**. Standard-of-care (SOC) clinical MRD testing by MFC was performed for all patients, with a subset (32.1%, n=18) having a genomic feature amenable to SOC single-variant molecular MRD testing (*FLT3-ITD*, *NPM1c*, *CBFB-MYH11*, or *RUNX1-RUNX1T1)*. 89.3% (n=50) of patients achieved an initial composite clinical remission (cCR1), of whom 68% (n=34) were MRD^−^ by combined SOC testing (MFC ± molecular) at best response. Among cCR1 responders, 64% (n=32) relapsed, including 20% (n=10) post-HSCT and 10% (n=5) with extramedullary relapse.

**Figure 2.**
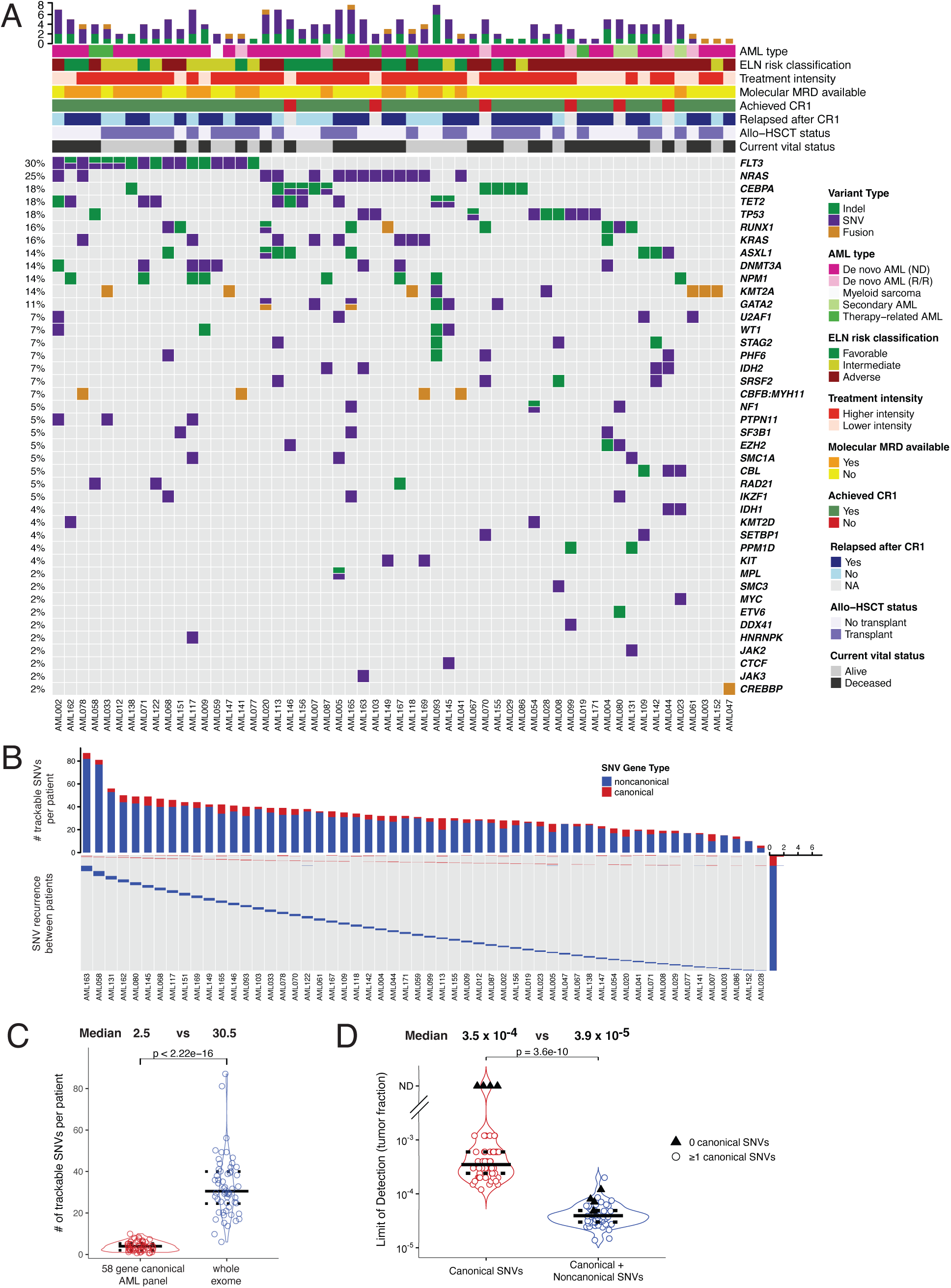
WES–informed personalization expands trackable mutations for sensitive MRD detection. (A) Oncoprint summarizing clinical characteristics and molecular features of the 56-patient cohort. (B) **Top:** Bar plot showing the number of trackable SNVs per patient identified by WES, stratified by canonical vs noncanonical genes. Asterisks denote patients lacking canonical SNVs, whose disease was trackable only using noncanonical variants. **Bottom:** Heatmap illustrating interpatient variant specificity. (C) Violin plot comparing the number of trackable SNVs per patient using a canonical-only panel versus a WES–integrated personalized approach. (D) Violin plot showing the theoretical per-patient limit of detection (LOD) for tumor fraction based on the number of trackable variants using canonical-only versus personalized mutation sets. Black triangles indicate patients without trackable canonical variants. ND, not detectable.

**Table 1:**
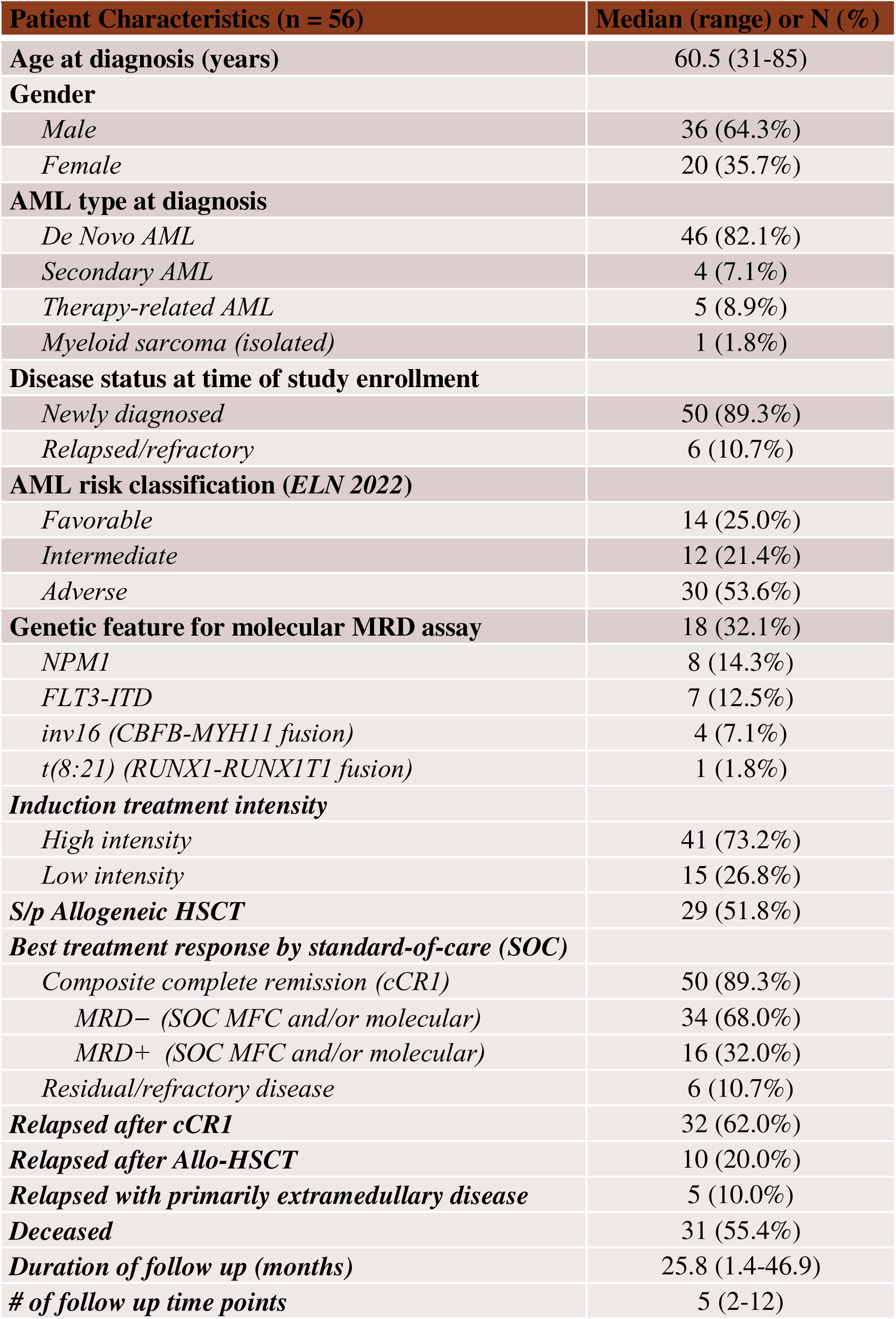
Demographic and clinical characteristics of AML patients including in the study.

### Personalized Variant Detection Enables Universal Molecular Disease Monitoring

Focusing on SNVs for robust NGS quantitation, WES identified a median of 30.5 somatic SNVs per patient (range: 6–87), allowing molecular MRD monitoring in 100% of cohort patients (**Figure 2B-C**). We observed a median of 2.5 (range 0-4) canonical AML-associated SNVs per patient compared to a median of 28 (range 4-82) noncanonical SNVs (p < 0.001, **Figure 2C**). Notably, 7.1% (4/56) of patients had zero canonical AML-associated SNVs available for SOC MRD tracking (**Figure 2B-2C**).

We found 99.9% of all noncanonical variants were uniquely specific to each individual patient (**Figure 2B**, heatmap), consistent with their role as personalized ‘passenger’ variants^14^. To further characterize them, we used multiple informatic tools, including phylogenetic conservation scores (PHAST^15^ and Zoonomia^16^), protein function impact predictions (SIFT^17^ and ESM^18^), and composite Combined Annotation Dependent Depletion (CADD)^19,20^ analyses, to assess predicted pathogenicity (**Figure S2**). In all analyses, we observed that noncanonical variants were significantly less conserved and less likely to affect protein function and/or pathogenicity (**Figure S2**), supporting their role as patient-specific passenger mutations which nonetheless function as effective biomarkers for personalized MRD monitoring. Noncanonical variants had higher median variant allele frequency (VAF) than canonical variants (39.3% vs. 25.6%, p < 0.001, **Figure S3**).

Because sensitivity in personalized MRD assays scales with the number of independently trackable tumor variants per patient, we next evaluated the theoretical limit of detection (LOD) for each patient using canonical (driver) variants alone versus incorporating both canonical/driver and noncanonical/passenger variants derived by initial WES. Using this patient-specific model which assumes 2500× sequencing depth (**Methods**), we found the median LOD (3.9 × 10 ) of the combined all-variant approach to be an order of magnitude lower than using only canonical/driver variants alone (3.5 × 10 ). These findings demonstrate the addition of noncanonical/passenger variants potentially enhances sensitivity and enables universal MRD monitoring.

### ctDNA Enhances Mutation Detection Compared to Cellular Compartments

To evaluate the utility of ctDNA as a biomarker for quantifying AML leukemic burden, we first compared variant to variant detection in paired plasma (from peripheral blood and bone marrow aspirate) and cellular (BMMC and PBMC) compartments. Among canonical AML variants identified at diagnosis by clinical NGS with a VAF > 1%, qualitative variant detection rates were highly concordant across all compartments, including 100% concordance between PB-ctDNA and BMMCs (**Figure S4**). Expanding the analysis to include all paired timepoints and VAF ranges, we observed significant quantitative correlations in VAF between ctDNA and cellular compartments across both canonical and noncanonical variants (**Figures 3A–3C**, PB-ctDNA vs BMMCs: ρ = 0.89; PB-ctDNA vs PBMCs: ρ = 0.84; BMMCs vs PBMCs: ρ = 0.88). All results were validated against our institutional CLIA-approved clinical NGS assay (Stanford HemeSTAMP^21^).

**Figure 3.**
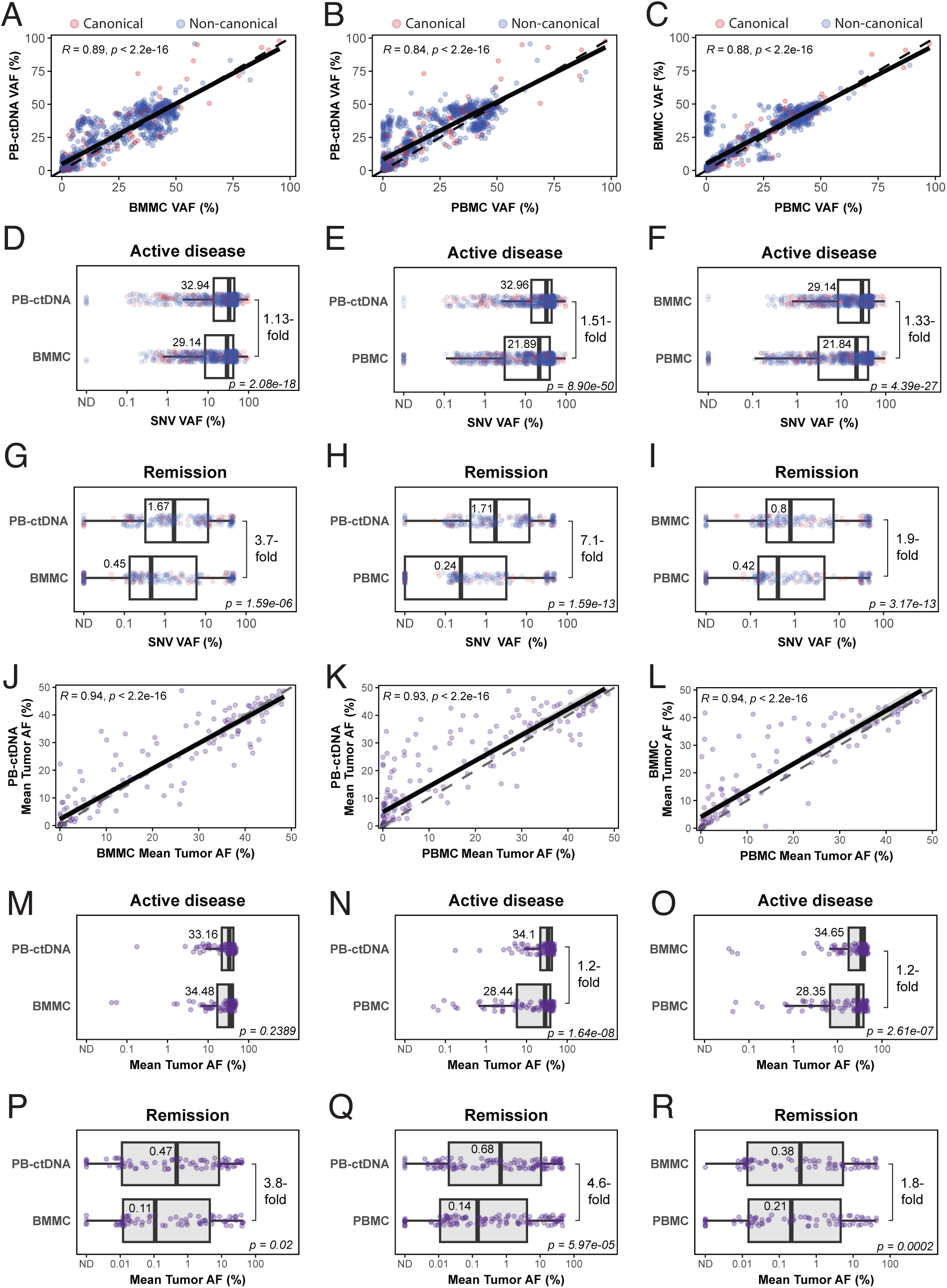
Comparison of ctDNA with cellular compartments for mutation detection and MRD quantification. (A–C) Correlation of SNV variant allele frequencies (VAFs) across all patients and timepoints between paired compartments: PB-ctDNA vs BMMC (A), PB-ctDNA vs PBMC (B), and BMMC vs PBMC (C). Box plots showing distribution of VAFs for shared SNVs across paired compartments during active disease (D–F) or during remission or MRD assessment timepoints (G–I). (J–L) Correlation of mean tumor allele fraction (AF) at the sample level across compartments: PB-ctDNA vs BMMC (J), PB-ctDNA vs PBMC (K), and BMMC vs PBMC (L). Box plots comparing distribution of mean sample tumor AFs in paired compartments during active disease (M–O) or remission/MRD timepoints (P–R). In all box plots, central bars indicate medians; statistical comparisons were performed using paired t-test with corresponding p-values shown.

To assess the performance of ctDNA as an analyte for MRD monitoring, we stratified samples based on standard remission criteria. During active disease (>5% blasts, **Figures 3D–3F**), PB-ctDNA exhibited modest enrichment in median SNV VAF compared to both cellular compartments (PB-ctDNA vs BMMCs: 1.13-fold, *p* < 0.0001; PB-ctDNA vs PBMCs: 1.51-fold, *p* < 0.0001); BMMCs vs PBMCs: 1.33-fold, *p* < 0.0001). During MRD assessment/remission states (**Figures 3G–3I**), ctDNA was markedly more sensitive and outperformed BMMCs (3.7-fold, *p* < 0.0001) and PBMCs (7.1-fold, *p* < 0.0001). Similarly, BMMCs also outperformed PBMCs (1.9-fold, *p* <0.0001). In addition, among paired samples in which the corresponding SNV VAF in BMMCs or PBMCs was <1% (**Figure S5A–S5B**), PB-ctDNA again demonstrated superior sensitivity compared to both BMMCs (2.1-fold, *p* < 0.0001) and PBMCs (5.5-fold, *p* < 0.0001), while BMMCs yielded a 3.1-fold enrichment over PBMCs (*p* < 0.0001, **Figure S5C**).

We next performed sample-level disease detection analyses between paired ctDNA and cellular compartments across all timepoints, by incorporating all individual patient-specific SNVs to reflect the aggregate tumor burden or mean tumor allele fraction (AF) for each sample (**Methods**) ^11,12,22^. First, we again observed strong correlation in the mean tumor AF compartments (PB-ctDNA vs BMMC: ρ = 0.94; PB-ctDNA vs PBMC: ρ = 0.93; BMMC vs PBMC: ρ = 0.94, **Figures 3J-3L**). Second, while PB-ctDNA and BMMCs had comparable AFs during active disease states (**Figures 3M-3O**), PB-ctDNA again demonstrated significantly increased sensitivity for MRD detection during remission states (**Figures 3P-3Q**) compared to both BMMCs (3.8-fold, *p* = 0.02) and PBMCs (4.6-fold, *p* < 0.0001). BMMCs remained superior to PBMCs (1.8-fold, *p* = 0.0002, **Figure 3R**). Third, at timepoints when the mean tumor AF in BMMCs or PBMCs was <1%, PB-ctDNA remained the most sensitive for MRD detection compared to both BMMCs (1.6-fold, *p* = 0.006) and PBMCs (3.5-fold, *p* < 0.0001, **Figure S6A-S6B**). BMMCs yielded a 1.4-fold enrichment over PBMCs (p = 0.0003, **Figure S6C**). Taken together, our results show ctDNA to be a superior biomarker with greater sensitivity for capturing AML disease burden.

Lastly, we also compared variant detection and composite MRD signal between PB-ctDNA and BM-ctDNA in a subset of patients (n=23). Paired plasma ctDNA samples showed nearly identical quantitative mutation profiles at both the variant and sample levels, spanning active disease and remission timepoints (**Figures S7A-S7B**), suggesting general equivalence of these cell-free compartments in representing systemic disease burden in AML. Thus, the remainder of the analyses reported here were conducted with PB-ctDNA.

### ctDNA-MRD Correlates with Clinical Outcomes and Outperforms Current Standard-of-Care MRD

To assess the clinical utility of ctDNA for outcome prediction, we compared AML-CAPP-Seq–derived MRD results with SOC MRD assays as applied in routine clinical practice. **Figure 4A** summarizes the clinical trajectories, disease status, and MRD assessments over time for all cohort patients (n=56).

**Figure 4.**
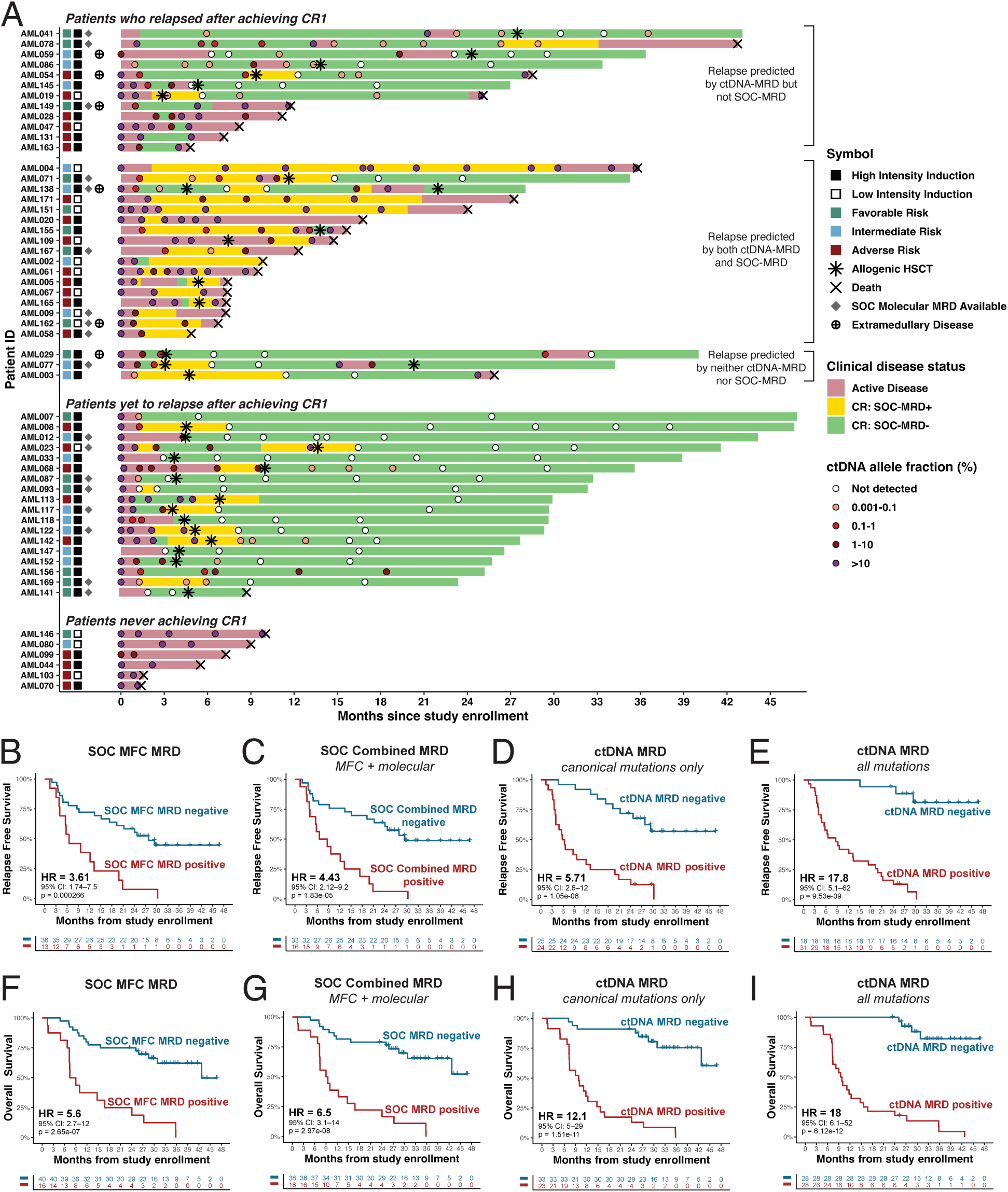
Comparison of ctDNA-MRD and SOC-MRD for clinical outcome prediction. (A) Swimmer plots showing disease course and MRD status for all AML patients (n=56) in the cohort. Bars represent time on study, annotated by standard-of-care (SOC) MRD status (MFC and/or single variant molecular testing when applicable), with superimposed dots indicating plasma ctDNA levels measured by AML-CAPP-Seq. (B-E) Kaplan Meier curves depicting relapse free survival (RFS) among patients who achieved initial composite CR (n=49), stratified based on SOC MFC MRD alone (B), SOC combined MRD (MFC + molecular, C), ctDNA-MRD using canonical AML gene mutations only (D), or ctDNA-MRD using all WES-derived personalized tumor variants (E). (F-I) Kaplan-Meier curves showing overall survival (OS) for all patients (n=56) stratified by achieving MRD negativity at any timepoint using SOC MFC MRD alone (F), SOC combined MRD (G), ctDNA-MRD using only canonical variants (H), or ctDNA-MRD using all personalized tumor variants (I). Hazard ratios (HR), 95% confidence intervals (CI), and p-values from log-rank tests are indicated for each comparison.

Among all cCR samples deemed to be MRD**^+^** by SOC methods, ctDNA-MRD was detectable in 96.7% (58/60), including 100% (40/40) by MFC and 93.1% (27/29) by single-variant molecular assays (**Figure S8**).

Conversely, in samples deemed to be MRD^−^ by SOC methods, ctDNA-MRD was notably detectable in 46.5% (67/144), including 47.8% (63/132) by MFC and 28% (7/25) by molecular testing (**Figure S8)**. These 67 SOC-MRD^−^/ctDNA-MRD^+^ samples corresponded to 28 unique patients who would have been classified as MRD positive by ctDNA at least once. ctDNA upstaging of MRD status occurred in 15 of 16 (93.8%) post-induction samples, 17 of 22 (77.3%) post-consolidation samples, and 8 of 14 (57.1%) pre-transplant samples that were SOC-MRD^−^ (**Figure S8**). In these samples where MRD status was upstaged by ctDNA, AML-CAPP-Seq accurately predicted clinical relapse in 50.7% (34/67) of patients who ultimately relapsed. The remainder 40.3% (27/67) patients later achieved ctDNA-MRD negative status after additional therapy and remain relapse-free (**Figure S8**). We note only two patients who currently remain ctDNA-MRD^+^ after completing therapy and have yet to relapse (AML156 and AML041; **Figure 4A**). Importantly, restricting ctDNA-MRD assessment to canonical AML variants markedly reduced sensitivity, detecting MRD in only 80% (48/60) of SOC-MRD^+^ samples and 20.8% (30/144) of SOC-MRD^−^ samples.

Among patients who achieved a cCR1 (n=50), ctDNA-MRD was positive in 90.3% (28/31) of patients prior to relapse, compared to 51.6% (16/31) detected by combined SOC MRD methods or 41.9% (13/31) detected by MFC MRD alone (**Figure 4A**). Of the 12 SOC MRD^−^ patients who had detectable ctDNA that correctly anticipated relapse, ctDNA-MRD became positive a median of 160 days (∼5.3 months; IQR, 49–367 days, **Figure 4A**, top panel) prior to clinical relapse. This group included 3 patients (AML086, AML047, and AML145) who lacked or lost trackable canonical molecular markers, but in whom personalized ctDNA-MRD identified persistent molecular disease (**Figure 4A**). For the only 3 patients (AML003, AML029, and AML077) who relapsed (all after alloHSCT) despite negative ctDNA-MRD and SOC-MRD status, no plasma samples for MRD analysis were collected within 168-583 days immediately preceding relapse (**Figure 4A**). Notably, AML-CAPP-Seq detected ctDNA-MRD and anticipated relapse in 5 of 6 patients (83.3%) who relapsed at primarily extramedullary sites (AML054, AML059, AML138, AML149, and AML162), compared to only 1 of 6 (16.7%) using only SOC MRD methods (**Figure 4A**).

In relapse-free survival (RFS) analysis of cCR1 patients stratified by MRD status at either the last pre-relapse timepoint (for relapsed patients, n=32) or the last available follow-up (for non-relapsed patients, n=17), MRD positivity by SOC MFC alone predicted inferior RFS (hazard ratio [HR] = 3.6, *p* = 0.0003), which was improved when combined with available SOC molecular MRD assays (HR = 4.4, *p* < 0.0001) reflecting real-world clinical practice (**Figures 4B-4C, Methods**). However, ctDNA-MRD outperformed SOC methods with superior prediction of relapse, even when relying only on canonical variants (HR=5.7, *p* < 0.0001) (**Figure 4D**). Strikingly, ctDNA-MRD incorporating personalized noncanonical variants discovered by WES was the most prognostic of RFS (HR=17.8, *p* < 0.0001, **Figure 4E**) with significant incremental predictive value beyond SOC and canonical-only MRD methods as demonstrated by nested likelihood ratio testing and improved model fit (**Table S7**, **Methods**).

Similar trends were observed for overall survival (OS) analysis across all cohort patients (n = 56, **Figure 4F-I, Table S7**) with personalized ctDNA-MRD most strongly predicting OS (HR 18.0, *p* < 0.0001) compared to SOC MFC (HR = 5.6, *p* < 0.0001), SOC combined MRD (HR = 6.5, *p* < 0.0001), or ctDNA-MRD with canonical variants only (HR = 12.1, *p* < 0.0001). Only 3 deaths (AML002, AML058, and AML141) were due to non-relapse mortality. We note 6 (of 56) patients with R/R disease were included in our study (**Tables S3/S5**), 3 of whom subsequently achieved cCR and were included in the primary RFS and OS analyses (**Figure 4)**. The same analyses using only newly diagnosed AML patients (n=50) yielded highly concordant results with minimal changes in effect sizes (**Figure S9**).

To clarify the clinical context in which personalized ctDNA-MRD assessment may provide greatest benefit, we performed subgroup analyses in patients with or without trackable driver mutations using SOC single-variant molecular MRD assays (*FLT3-ITD*, *NPM1c*, *CBFB-MYH11*, or *RUNX1-RUNX1T1)*. Among the majority of patients (70%, n=38) without a trackable variant for whom MFC is the only guideline recommended MRD modality, personalized ctDNA-MRD markedly improved RFS prediction compared to SOC MFC (HR 13.8, p < 0.0001 vs HR 2.6, p = 0.02, **Figure S10A-C**). Among patients with SOC single-variant molecular assays (n=18, **Figure S10D-F, Figure S11**), ctDNA-MRD remained strongly prognostic and outperformed SOC single-variant plus MFC MRD (HR 21.5, p = 0.0001 vs HR 9.1, p = 0.0004). Although in the latter, sample size was limited and molecular testing was done in a ‘real-world’ non-uniform manner.

Likewise, adverse-risk AML is a genomic subgroup in which MFC-based MRD assessment has lower prognostic utility and trackable driver mutants are typically unavailable^23^. In subgroup analyses performed for this group at high risk of relapse (n = 30, **Figure S12**), we found personalized ctDNA-MRD markedly outperformed SOC MRD approaches including MFC for both RFS (HR 13.1, p < 0.0001 vs HR 2.5, p = 0.04) and OS (HR 9.5, p < 0.0001 vs HR 2.5, p = 0.02).

Despite the growing interest and utilization of PBMCs for AML MRD detection, ctDNA-MRD integrating canonical and personalized SNVs also outperformed both PBMCs (HR = 17.6 vs 8.6, both *p* < 0.0001, n = 48) and BMMCs (HR = 28.2, *p* < 0.0001 vs HR = 7.0, *p* = 0.0006, n = 28) for RFS prediction (**Figure S13A–S13B**), and similarly for OS (HR = 17.0 vs 13.8 vs 11.7 for ctDNA, BMMCs, and PBMCs respectively, *p* < 0.0001 for all; **Figure S13C**) among patients with paired samples. These findings highlight the superior prognostic accuracy of our truly personalized approach for AML MRD detection using PB-ctDNA compared to current MRD assays.

### Association of ctDNA Clearance Kinetics with Relapse

We next evaluated the timing and kinetics of ctDNA-MRD clearance and their association with RFS. Among patients who achieved ctDNA negativity at any timepoint (n = 27), the median time to ctDNA clearance was 221 days (IQR, 178.5–406 days), substantially later than clearance by SOC MRD assays (median, 70 days; IQR, 35.5–177.25 days). To assess whether the kinetics of ctDNA clearance impacted outcome, patients were stratified by early versus late clearance (<180 vs ≥180 days). Although both early and late ctDNA-MRD clearance was associated with markedly improved RFS compared with patients who never achieved ctDNA-MRD negativity, the timing of clearance itself did not significantly differentiate RFS between these two groups (**Figure S14A**). In contrast, patients who maintained ctDNA negativity for ≥ 24 months (n = 7) had significantly superior RFS compared with those with shorter durations of ctDNA negativity (< 24 months; n = 20; HR 14.4; p = 0.006). Both groups with sustained ctDNA negative status experienced substantially better clinical outcomes compared to those who never achieved ctDNA clearance (n = 22). Our data suggest duration of ctDNA negative status maybe strongly associated with RFS (**Figure S14B**).

### ctDNA-MRD Dynamics Predict Outcomes in the Peri-Transplant Setting

MRD status in the peri-transplant setting is an important clinical evaluation landmark. We applied AML-CAPP-Seq to evaluate the prognostic utility of ctDNA-MRD assessment in the peri-transplant setting among patients in our cohort who underwent alloHSCT (n=29, **Figure 5A**). As expected, pre-HSCT MRD positivity was associated with inferior RFS post-transplant. While stratification by SOC MRD methods showed relatively modest prognostic separation (HR = 2.2, p = 0.22), ctDNA-MRD yielded a stronger quantitative association with RFS (HR = 6.1, p = 0.097) with all patients who achieved ctDNA-MRD^−^ status pre-transplant remaining relapse-free post-HSCT with persistently undetectable disease (**Figure 5B**).

**Figure 5.**
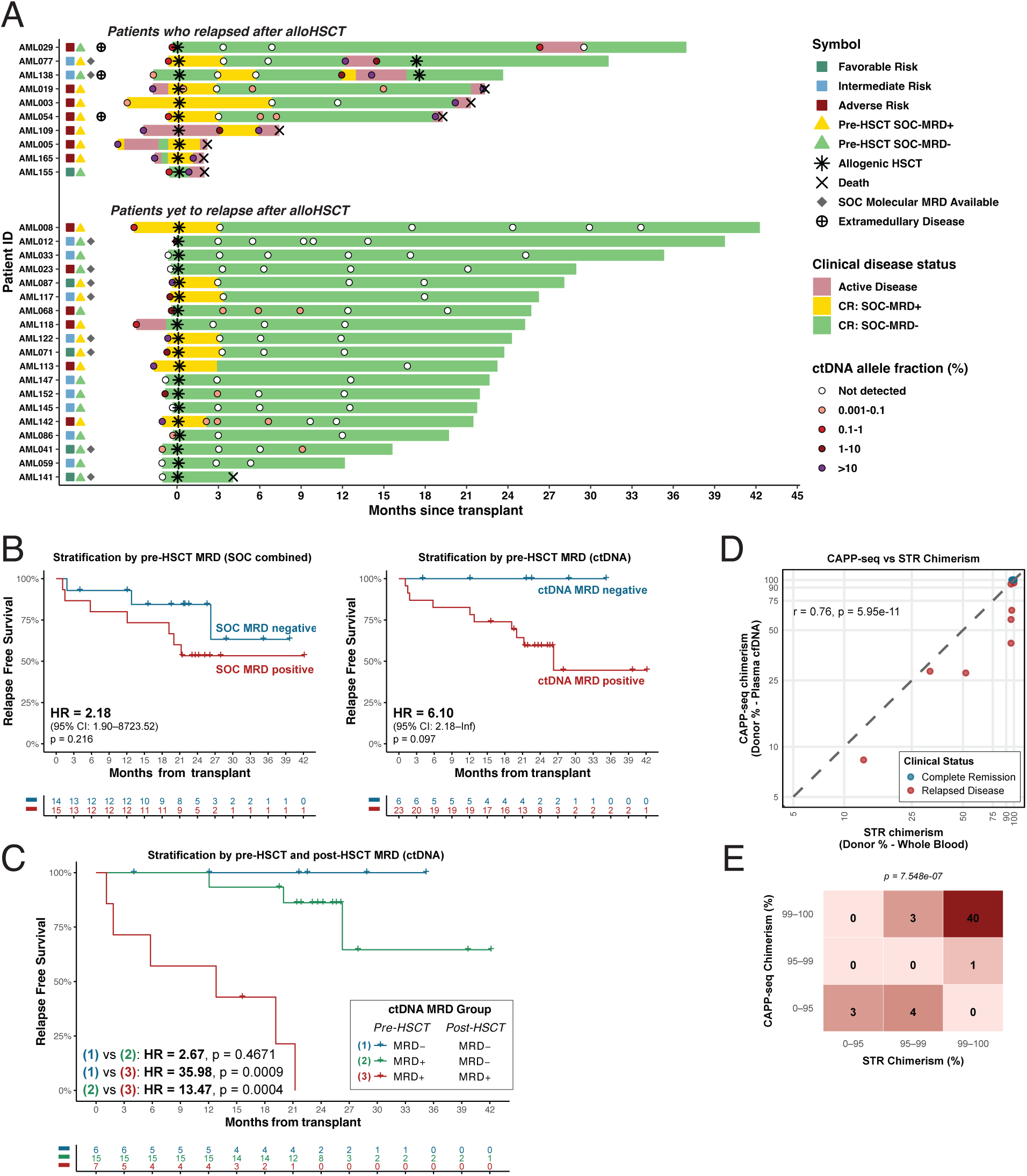
Clinical utility of cfDNA for peri-transplant MRD risk stratification and chimerism assessment. (A) Swimmer plots for AML patients who underwent allogeneic HSCT after achieving initial CR (n=29). Bars represent disease course and MRD status based on SOC combined MRD testing (MFC and/or molecular assays), with overlaid dots reflecting quantitative ctDNA levels measured by AML-CAPP-Seq. (B) Kaplan–Meier curves depicting post-HSCT relapse-free survival (RFS) stratified by MRD status at the most recent pre-transplant timepoint, using SOC combined MRD (left) or plasma ctDNA-MRD (right). (C) RFS stratified by dynamic ctDNA-MRD trajectories before and after transplant: patients who remained ctDNA-negative (Group 1), converted to negative (Group 2), or remained ctDNA-positive (Group 3) post-HSCT. (D) Top: Correlation plot comparing donor-recipient chimerism estimates from plasma cfDNA via AML-CAPP-Seq versus standard STR-based testing across paired post-transplant samples (n=51). Dots are colored by clinical disease status at the time of sample collection (remission vs relapse). Bottom: Chi-square contingency plot showing categorical concordance between cfDNA- and STR-based chimerism methods at clinically relevant thresholds (Fisher’s exact test p-value shown).

Notably, patients who remained ctDNA-MRD**^+^** post-HSCT (**Figure 5C**, *Group 3*) uniformly relapsed (HR = 36.0, p = 0.0009). This included 3 patients (AML019, AML054, AML138) who initially cleared their ctDNA-MRD after HSCT but reconverted to ctDNA-MRD^+^ months before relapse. In contrast, those who converted to durable ctDNA-MRD negative status post-HSCT (*Group 2*) experienced significantly improved RFS (HR = 13.5, p = 0.0004). The median time to post-transplant ctDNA clearance was 90 days (IQR, 85.5–137 days) among patients who achieved ctDNA-MRD negativity after alloHSCT (**Table S6**). 3 patients (AML068, AML142, and AML145) demonstrated ctDNA-MRD clearance beyond day 90 and have remained relapse-free with sustained ctDNA negativity during follow-up. As previously mentioned, ctDNA-MRD did not predict relapse in 3 patients (AML029, AML077 and AML003) due to lack of sampling in the > 5.5 months immediately preceding relapse.

For comparison, we also evaluated peri-HSCT MRD assessment using conventional bone marrow–based NGS performed as part of routine clinical care using our institutional NGS assay (Stanford HemeSTAMP). Among the 26 patients with known canonical driver mutations and evaluable clinical NGS result, stratification by positive or negative status pre- or post-HSCT did not significantly predict post-transplant RFS (**Figure S15A-B**). In contrast, ctDNA-MRD–based risk stratification outperformed standard clinical NGS based on achieving MRD negative status either before or after alloHSCT (**Figure S15A-B**). These observations further illustrate the prognostic value of peri-transplant ctDNA dynamics over SOC metrics for identifying patients at highest risk of relapse and windows where post-alloHSCT interventions may improve outcomes.

### AML-CAPP-Seq Allows Assessment of Donor-Recipient Chimerism Post-HSCT

Early monitoring of chimerism dynamics can inform post-HSCT decisions around immunosuppression, donor lymphocyte infusion (DLI), or maintenance treatment in preventing fulminant clinical relapse^24,25^. Current methods using short tandem repeat (STR) analysis using cellular based methods are limited by the number of informative loci per individual and infrequent testing intervals. Using donor-derived single-nucleotide polymorphisms (SNPs) captured in both the canonical and personalized selector regions, we observed that AML-CAPP-Seq can accurately and sensitively quantitate donor chimerism using PB-cell-free DNA (cfDNA). We identified a median of 8 donor SNPs (range: 1–16) per alloHSCT recipient (**Figure S16**), enabling robust, serial quantification of donor-derived cfDNA. We found strong correlation (ρ = 0.76, p < 0.0001) and concordance between PB-cfDNA and STR-derived chimerism at key clinical thresholds (e.g., >99% or <95% donor, p < 0.0001; **Figure 5D–E**). Notably, PB-cfDNA detection of chimerism loss was more prominent compared to STR-based analysis in 6 cases of post-alloHSCT relapse (**Figure 5D**).

### AML-CAPP-Seq Resolves Clonal Dynamics

To gain a better understanding of AML leukemogenesis, we analyzed changes in known recurrent driver mutations between diagnosis and relapse in those who experienced relapsed disease (**Figure S17,** n=31). Overall, mutations in epigenetic regulators (e.g., *DNMT3A*, *ASXL1*), myeloid transcription factors (e.g., *CEBPA*), spliceosome components (*U2AF1*, *SF3B1*), *NPM1*, and *TP53* were largely stable between diagnosis and relapse, consistent with their roles in early leukemogenesis and in contrast to dynamic changes observed in signal transduction mutations (*FLT3-ITD*, *KRAS*, and *NRAS)*. Mutations most frequently gained at relapse included *NRAS*, *PTPN11*, *WT1*, and *NF1*, suggesting selection for clones harboring these alterations under therapeutic pressure and consistent with reported literature^26–31^.

Understanding clonal evolution under therapeutic pressure in AML has been hampered by its genomic heterogeneity and the sparsity of mutations detectable using fixed canonical NGS panels (**Figure 2B-2C**)^32,33^. We hypothesized that addition of noncanonical variants from the personalized selector, together with time-series analysis, can better inform and resolve clonal architecture. Herein we present and discuss three representative AML cases in which we were able to accurately recapitulate clonal dynamics by integrating personalized noncanonical variants discovered with time-series longitudinal monitoring (**Figures 6A–6C**).

**Figure 6.**
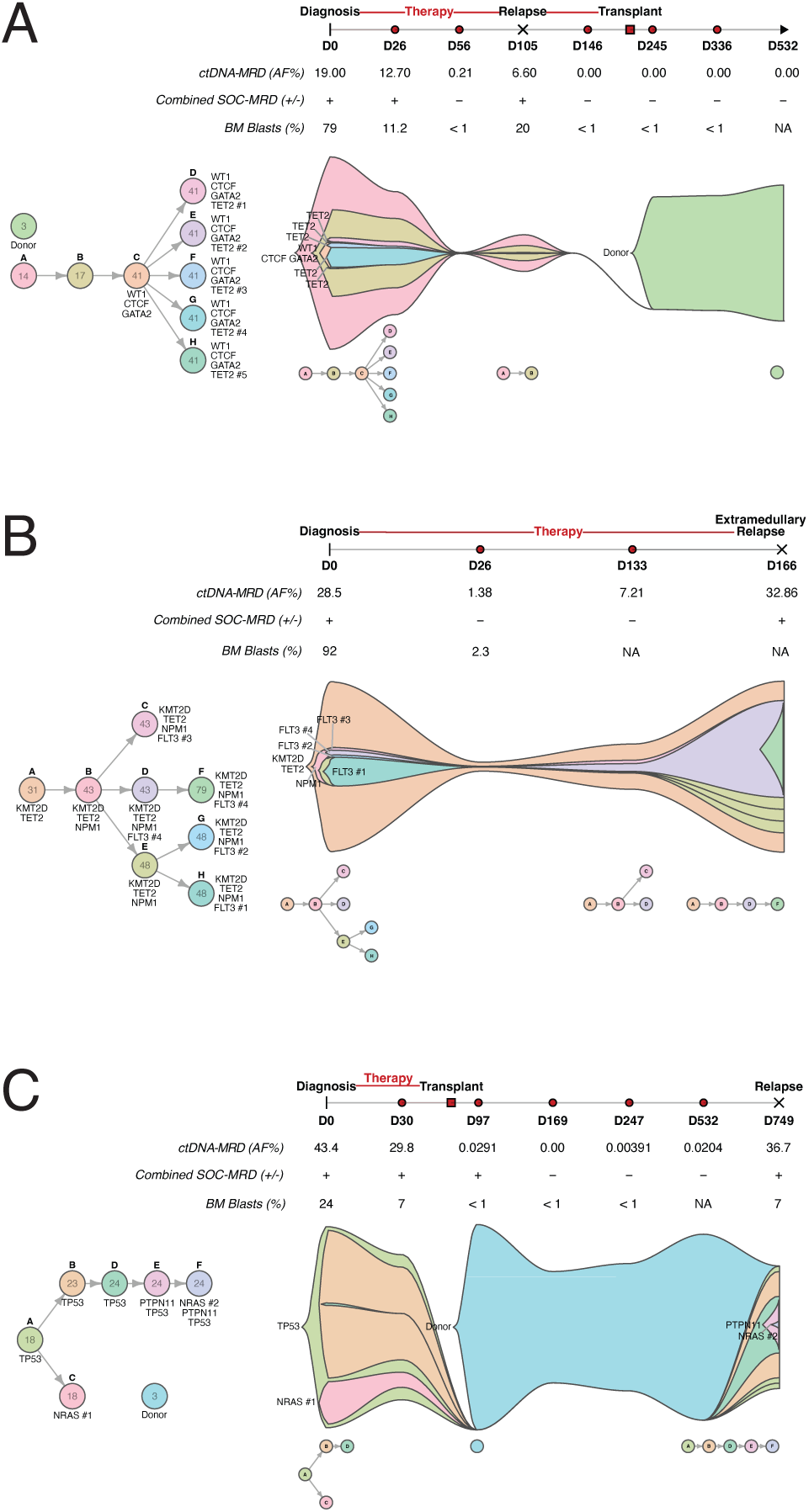
AML-CAPP-Seq enables reconstruction of clonal evolution and resolution of relapse dynamics. (A–C) Clonal reconstructions for three representative patients (AML145, AML162, AML019) with serial ctDNA samples, showing patient-specific clonal architecture, subclonal dynamics, and integration with MRD and clinical course. Colored fish plots (right) depict inferred clonal hierarchy (left) and evolution over time based on SNV cluster assignments and variant allele frequencies. Canonical variants within each clone are listed, and the number of accompanying noncanonical variants is indicated numerically.

#### Case 1 – AML145 (Figure 6A)

*A middle-aged female with intermediate-risk AML characterized by WT1, GATA2, and five distinct TET2 mutations along with normal karyotype at diagnosis. Despite achieving an MRD^−^ CR1 by MFC after induction treatment, ctDNA-MRD remained detectable, and she experienced early relapse (Day 105). She received salvage therapy and attained CR2 followed by durable MRD^−^ remission by both MFC and ctDNA post-HSCT* (**Figure 4A**). AML-CAPP-Seq identified multiple evolving clones, including the primary clone present at relapse (clone A), which was defined exclusively by noncanonical SNVs without any known recurrent driver mutation. This case underscores the ability of personalized variant detection to identify clinically important clones which would not otherwise be detectable.

#### Case 2 – AML162 (Figure 6B)

*An elderly male with favorable-risk AML harboring NPM1c*, *TET2*, *and four distinc*t *FLT3*-TKD *mutations. He achieved an MRD^−^ CR by SOC single-variant NPM1c testing after initial induction but experienced relapse 4.7 months later with multiple sites of extramedullary AML without detectable marrow relapse* (**Figure 4A**). At diagnosis, AML-CAPP-Seq identified a founding *TET2/KMT2D* defined clone (clone A) and an *NPM1* containing subclone (clone B) that then gave rise to four distinct *FLT3-TKD* subclones at diagnosis (of which clone C was most abundant initially). He remained ctDNA-MRD**^+^** during remission with expansion of clone F as the dominant and only surviving FLT3-TKD subclone at relapse. This case highlights the unique ability of ctDNA to capture residual disease in extramedullary sites inaccessible to SOC methods and inform pre-emptive targeted (ex. FLT3-directed) therapeutic strategies.

#### Case 3 – AML019 (Figure 6C)

*A middle-aged male with TP53-mutant complex karyotype adverse-risk therapy-related AML initially treated with decitabine/venetoclax who achieved an MRD*^−^ *CR1 by MFC post-alloHSCT but ultimately relapsed ∼21 months later* (**Figure 4A**). AML-CAPP-Seq revealed a founding *TP53*-mutated clone (clone A) with multiple other subclones at diagnosis defined by both canonical (*NRAS*, clone C) and noncanonical variants (clones B, D). Aided by inclusion of noncanonical SNVs, clonal analysis demonstrated loss of the initial *NRAS*-containing subclone (clone C) at relapse and emergence of new subclones from within clone D harboring a *PTPN11* (clone E) and a second *NRAS* mutation (clone F), while resolving donor-recipient chimerism dynamics. Notably, rising ctDNA levels post-HSCT anticipated clinical relapse by approximately six months despite sustained MFC MRD negativity, highlighting a potential window for earlier post-transplant intervention prior to overt relapse.

## Discussion

In this study, we overcome limitations posed by the mutational sparsity and genomic heterogeneity of AML by developing AML-CAPP-Seq for peripheral blood plasma enabled ctDNA-MRD monitoring. We demonstrate that individualized ctDNA-based disease quantification permits highly sensitive, longitudinal MRD assessment that is broadly applicable across all AML genotypes. Using a cohort of 56 AML patients, we show that (1) ctDNA consistently outperformed both BM and PB cellular compartments for detecting MRD and predicting clinical outcomes and that (2) WES-informed personalization enables substantially enhanced MRD sensitivity and prognostic power beyond SOC MRD assays.

By robustly comparing AML variant detection between PB and BM cell-free and cellular compartments in > 1100 biological samples, we found that ctDNA captured all canonical driver mutations identified by clinical NGS and outperformed both BMMCs and PBMCs for detecting low level variants (VAF <1%), resulting in superior prediction of RFS and OS with > 2 years of follow up. These results support our hypothesis that plasma ctDNA more accurately reflects total-body AML burden, overcoming known MRD detection difficulties during states of marrow hypocellularity (e.g., morphologic leukemia-free states and pancytopenia after chemotherapy) or extramedullary disease without marrow involvement. Notably, our data clearly demonstrate the superiority of ctDNA and BMMCs over PBMCs for MRD detection—an important finding in AML, where PBMC-based assays are increasingly used for convenience despite limited validation, and where the advantage of ctDNA as an analyte over PBMCs in blood-based malignancies has not been well established.

A central conceptual advance of AML-CAPP-Seq is the use of WES-informed personalization in AML to expand the number of trackable tumor-specific variants per patient. By identifying a median of 30.5 somatic SNVs per individual, this approach markedly increased MRD detection sensitivity compared with fixed canonical panels and enabled MRD assessment in 100% of patients in our cohort. In contrast, currently approved single-variant molecular MRD assays, while capable of achieving higher analytical sensitivity than MFC, are amenable only to 30-40% of AML patients with *FLT3-ITD*, *NPM1c*, or core-binding factor mutations^6^. Under standard clinical sequencing depths (∼2500x), aggregating MRD signal across dozens of patient-specific variants yielded an order-of-magnitude improvement in theoretical per-patient limit of detection (median 3.9 × 10 ) relative to canonical-only approaches. Importantly, our individualized ctDNA approach using noncanonical ‘passenger’ variants allowed MRD monitoring in AML patients without known driver mutations at diagnosis which represents approximately 10% of all AML patients^3^.

These conceptual gains translated directly into superior clinical performance. While ctDNA assessment restricted to canonical AML variants alone yielded modest improvements in RFS and OS prediction over SOC marrow-based MRD approaches (HR 5.7 vs 4.4 for RFS; HR 12.1 vs 6.5 for OS), incorporating noncanonical variants substantially amplified both MRD detection sensitivity and prognostic power (HR 17.8 for RFS; HR 18 for OS). Importantly, across all analyses, personalized ctDNA-MRD demonstrated statistically significant incremental predictive value beyond SOC MRD and canonical-only ctDNA approaches, as evidenced by consistent improvements in model discrimination (higher C-indices), superior model fit (lower AIC), and highly significant likelihood ratio testing (**Table S7**). Furthermore, among the majority of patients who lack approved single-variant molecular MRD markers —many of whom fall into adverse-risk categories and for whom MFC remains the only available MRD modality —personalized ctDNA-MRD demonstrated markedly stronger associations with RFS and OS compared to SOC assessment, highlighting its immediate potential to address a substantial unmet clinical need in AML. Although our sample size limited direct head-to-head comparisons with single-variant MRD assays, ctDNA-MRD remained prognostic even in the minority of patients with approved molecular markers, supporting a complementary role particularly in settings of clonal evolution or loss of the original molecular marker.

Beyond binary MRD status, our data highlight the importance of ctDNA clearance dynamics as a clinically meaningful endpoint. ctDNA-MRD clearance occurred later than clearance by SOC assays, consistent with its greater analytical sensitivity; however, early versus late ctDNA clearance did not stratify outcomes. Instead, the duration of ctDNA negativity emerged as the strongest predictor of relapse, with all patients who maintained ctDNA-negative status for more than two years remaining relapse-free to date. These findings suggest that sustained ctDNA clearance may represent a surrogate biomarker of long-term disease control—and potentially cure—warranting prospective validation.

The prognostic value of ctDNA-MRD was particularly striking in the peri-transplant setting. While SOC MRD positivity pre- and post-transplant is associated with high relapse rates and poor OS, effective MRD-directed interventions remain limited —exemplified only by the use of post-transplant gilteritinib to improve RFS in FLT3-ITD**^+^** AML patients with detectable pre-transplant FLT3-ITD molecular MRD in the recent Phase 3 MORPHO trial^34,35^. In contrast, our strategy of personalized longitudinal MRD monitoring allowed robust relapse risk stratification across all transplanted patients regardless of mutational genotype (HR 36.0 for ctDNA-MRD^-^ vs ctDNA-MRD**^+^**), markedly outperforming SOC approaches. Notably, all 6 patients who achieved ctDNA negativity prior to transplant remain relapse-free, whereas persistent ctDNA positivity following alloHSCT uniformly predicted relapse—including cases in which rising ctDNA levels were detected more than one year prior to fulminant relapse (e.g., AML019 and AML054). Importantly, the vast majority of patients who cleared ctDNA after transplant achieved durable remission, including 3 patients who demonstrated delayed ctDNA-MRD clearance beyond day 90 consistent with ongoing graft-versus-leukemia effects. The only 3 relapses observed in this group—and in the entire cohort following ctDNA negativity—occurred in individuals without plasma sampling in the preceding >5.5 months, likely reflecting real-world sampling limitations rather than true assay failure. cfDNA also enabled donor–recipient chimerism assessment via SNP tracking, allowing more prominent detection of donor chimerism loss compared to clinical assays. Together, these findings highlight serial ctDNA-MRD surveillance as a practical framework for implementing post-alloHSCT interventions such as maintenance therapy, immunosuppression withdrawal, or donor lymphocyte infusion strategies to optimize graft-versus-leukemia effects, while recognizing that these observations warrant further validation in larger cohorts.

Biologically, we view AML-CAPP-Seq by design as a measure of aggregate leukemia burden that integrates signal across multiple leukemia-associated variants rather than a test for any single leukemic clone. We do not *a priori* exclude mutations associated with clonal hematopoiesis unless they are detected in paired germline T-cell controls for selection of the best tumor-associated variants, nor do we attempt to definitively classify individual variants as leukemic versus pre-leukemic. Importantly, exclusion of DTA mutations had no impact on MRD performance of AML-CAPP-Seq, indicating that the composite personalized MRD signal—irrespective of the biological origin of individual variants—remains highly clinically informative and prognostic compared to SOC MRD metrics in AML. Larger prospective studies, potentially incorporating single-cell and lineage-tracking approaches, will be required to refine these biological distinctions. Notwithstanding these complexities, our findings demonstrate that broad mutational profiling with inclusion of noncanonical variants enables higher-resolution tracking of clonal dynamics and relapse-associated molecular changes, including identification of potentially novel drivers of disease.

Certain limitations warrant consideration. This was a retrospective, single-institution study comprising AML patients who received heterogeneous treatments and underwent sampling at non-uniform intervals, reflecting real-world practice but limiting definitive conclusions regarding optimal clinical implementation of ctDNA-MRD. Nevertheless, the strong and consistent effect sizes observed across multiple analyses—including in clinically relevant subcohorts such as transplant recipients and patients lacking approved molecular MRD markers—support the robustness of our conclusions and establish this work as a compelling proof-of-concept for the utility of personalized ctDNA-MRD in AML. These findings emphasize the need for larger, prospective, multi-center studies using harmonized protocols and well-defined AML subpopulations to validate and extend our observations, including defining optimal monitoring intervals, response thresholds, and MRD-guided intervention strategies. Although our study was not powered to assess RFS or OS prognostication among patients receiving lower intensity therapy, we also demonstrate that ctDNA-MRD enables longitudinal disease monitoring for this vulnerable population, who often remain on indefinite treatment without adequate surrogate biomarkers to guide treatment duration or therapy de-escalation^36^.

From a practical standpoint, while the current AML-CAPP-Seq workflow requires upfront WES and germline DNA from sorted T cells to define personalized variant panels and achieve high analytical sensitivity, future work will focus on implementing germline-free informatic methods for high-fidelity somatic variant calling. Importantly, personalized WES-based ctDNA assays using typical cfDNA yields from standard blood draw volumes are increasingly feasible in clinical workflows. Most notably, Signatera (Natera) is now commercially available for MRD monitoring across multiple solid tumor indications^37^, with assay design turnaround times of ∼3–4 weeks and subsequent MRD results available within 7–10 days, parameters compatible with routine AML management and longitudinal surveillance.

In conclusion, personalized ctDNA monitoring offers a powerful, minimally invasive approach for high-sensitivity, genotype-agnostic MRD detection and disease tracking in AML, surpassing the prognostic performance of clinical SOC assays. By linking durable ctDNA-MRD negativity with long-term relapse-free survival, our study suggests that ctDNA clearance could serve as a meaningful surrogate endpoint for cure in AML—an insight with major implications for trial design and individualized patient counseling. Our immediate future work is aimed at translating these capabilities into ctDNA-MRD–guided clinical trials for AML to refine risk-adapted strategies and ultimately improve survival outcomes for this challenging disease.

## Materials and Methods

### Patients

A total of 56 patients with acute myeloid leukemia (AML) who were treated at Stanford Cancer Center, CA, USA, were included in this study. Patient selection was prioritized based on the availability of tumor samples collected at the time of AML diagnosis for whole-exome sequencing (WES) genotyping and the number of follow-up samples available. Among these patients, 50 had newly diagnosed treatment-naïve AML (4 with *de novo* AML, 4 with secondary AML from prior myelodysplastic syndrome or myeloproliferative neoplasm, 5 with therapy-related AML, and 1 with isolated myeloid sarcoma), while 6 patients presented with relapsed and/or refractory (R/R) AML at time of study enrollment. The cohort included both male and female adult patients spanning a broad age range reflective of the clinical AML population treated at our institution. Patient demographic characteristics, including age at diagnosis and sex, are summarized in **Supplementary Tables S2 and S3**. Patient IDs (“AMLxxx”) were not known to anyone outside the research group.

AML risk stratification was assigned uniformly according to European LeukemiaNet (ELN) 2022 criteria for all patients, regardless of treatment intensity, based on diagnostic cytogenetic and molecular features. Treatment intensity was classified as high-intensity therapy, defined as conventional intensive induction regimens (e.g., 7+3–based or FLAG-Ida–based approaches, including cladribine-containing regimens), or lower-intensity therapy, defined as hypomethylating agent (HMA)–based regimens administered with or without venetoclax, as well as oral targeted therapies directed at FLT3 or IDH1/2 mutations. Detailed patient-level information regarding frontline and subsequent treatment regimens, treatment intensity, genomic features, and clinical outcomes is provided in **Supplementary Tables S2–S5**.

Composite clinical remission (cCR) was defined as either complete remission (CR) with full hematologic recovery, CR with incomplete/partial hematologic recovery (CRi/CRp), or morphologic leukemic free state (MLFS) without hematologic recovery. Patients with R/R AML at study enrollment were included in relapse-free survival (RFS) analyses if they subsequently achieved a cCR during the study period, consistent with standard definitions of RFS.

The study was approved by the Stanford Institutional Review Board (protocol #18329) in accordance with the Declaration of Helsinki, and all patients provided written informed consent, including consent for genomic sequencing analyses. Longitudinal samples—including plasma (both peripheral blood and bone marrow derived), peripheral blood mononuclear cells (PBMCs), and bone marrow mononuclear cells (BMMCs)—were collected at clinically relevant timepoints throughout the treatment course, including diagnosis, post-induction, post-consolidation, pre/post-allogenic stem cell transplant (∼Day 90, ∼Day 180, ∼1 year), relapse, and approximately every 3 months thereafter in surveillance or maintenance therapy when available. Not all patients had evaluable samples available at every clinical timepoint due to real-world variation in sample availability, treatment course, and biospecimen banking logistics; all available samples meeting quality and coverage thresholds were included, and no samples were excluded based on clinical outcome. All samples were fully de-identified, and the patient identifiers used in the manuscript cannot be linked to individual patients by anyone outside the research team.

### Sample Collection and Processing

Peripheral blood and bone marrow aspirate samples were collected in EDTA tubes (BD, Franklin Lakes, NJ) and processed within 24 hours. Plasma was isolated via sequential centrifugation at 1,400 RPM for 10 minutes and then stored at –80°C. Peripheral blood mononuclear cells (PBMCs) and bone marrow mononuclear cells (BMMCs) were isolated using the Ficoll-Paque (Cytiva, Marlborough, MA, RRID:SCR_013163) density gradient centrifugation method, following the manufacturer’s protocol. PBMCs and BMMCs were cryopreserved in freezing media (90% FBS, 10% DMSO) and stored in liquid nitrogen.

Cell-free DNA (cfDNA) was extracted from 1.5-8 mL of plasma reflecting real-world clinical sample availability using the QIAamp Circulating Nucleic Acid Kit (Qiagen, Hilden, Germany, RRID:SCR_017340), according to the manufacturer’s protocol. Cellular DNA was isolated from PBMCs, BMMCs, or purified blasts and T cells using the QIAamp DNA Micro Kit (Qiagen, RRID:SCR_012948) following the manufacturer’s instructions. Following isolation, cellular DNA was mechanically fragmented to a target size of 170 bp using a Covaris S2 sonicator (Covaris; RRID:SCR_018210) or enzymatically fragmented using the KAPA HyperPlus Kit (Roche, Pleasanton, CA, RRID:SCR_000593), according to the manufacturer’s protocol. DNA concentration was quantified using the Qubit dsDNA High Sensitivity Kit (Thermo Fisher Scientific, Waltham, MA, RRID:SCR_018095), and fragment length was assessed using an Agilent Bioanalyzer or Fragment Analyzer (Agilent, Santa Clara, CA, RRID:SCR_018043). For downstream library preparation, 5–50 ng of cfDNA or 50-75 ng of cellular DNA was used per sample, depending on yield.

### Blast and T cell Isolation for WES

For WES genotyping, AML blasts were purified from diagnostic timepoint PBMC or BMMC samples by flow cytometry when available; otherwise, WES was performed on bulk cellular DNA from whole blood (bone marrow or peripheral). T cells were isolated from either flow-sorted CD3+ T cells (see below) or CD3+ magnetic bead-purified T cells (EasySep Human CD3 Positive Selection Kit II, Stemcell, Vancouver, RRID:SCR_014955) per manufacturer’s instructions.

For flow cytometry sorting, cryopreserved samples were thawed at 37°C and diluted with pre-warmed thawing media (IMDM with 20% FBS and 1x penicillin/streptomycin). After centrifugation at 300g for 5 minutes, cells were initially resuspended in thawing media containing DNase I (100 U/mL), washed and then subsequently resuspended in FACS buffer. Cells (5×10^6^ per FACS tube) were stained with a master mix of CD45-V450 (BD Horizon, Cat. #560368), CD3-PeCy5 (BD Pharmingen, Cat. #555334), CD11b-APC (Biolegend, Cat. #301309), and CD14-FITC (Biolegend, Cat. #367115) in FACS buffer for 25 minutes at 4°C (dark). After washing, cells were resuspended in 500 μL FACS buffer containing propidium iodide (PI) for viability staining. Sorting was conducted on the FACS Aria II flow cytometry (BD Biosciences, RRID:SCR_01809), excluding dead cells and doublets based on forward-scatter, side-scatter, and PI staining. Myeloid cells were separated from lymphocytes using CD45 staining and side-scatter profile, and blasts were then identified by CD11b and CD14 expression patterns, guided by match clinical flow cytometry reports (**Figure S1**). T cells were gated based on CD3 expression, with purity confirmed to be greater than 95% for each sorted population (**Figure S1**).

### Hybrid Capture Panel Generation

#### Canonical Selector Design

To detect recurrent genomic alterations in AML, we designed a custom AML-specific CAPP-Seq panel using publicly available whole-genome sequencing (WGS) and whole-exome sequencing (WES) data from the BeatAML^13^ and The Cancer Genome Atlas (TCGA)^2^ databases to identify candidate genes. This 225kB panel encompasses the coding regions of 58 genes known to be recurrently mutated or of biological significance in AML (**Table S1**). A set of 120 bp biotinylated custom oligonucleotides (IDT, Coralville, IA, RRID:SCR_011151) was designed to tile across the coordinates of these genes for use in hybrid capture.

#### Personalized Selector Design

To generate each patient’s personalized panel for targeted MRD detection, we performed WES on diagnostic timepoint tumor samples, which included cellular DNA from either flow-purified AML blasts (when available) or cellular DNA derived from peripheral blood or bone marrow aspirate obtained from the Stanford Clinical Lab, along with matched T cells to exclude germline variants. Whole-exome sequencing was performed with a target mean coverage of approximately 300× for tumor samples and matched germline controls to enable sensitive detection of somatic variants for personalized MRD tracking. Single nucleotide variants (SNVs) were identified using three independent variant callers (Strelka RRID:SCR_005109, MuTect RRID:SCR_000559, Varscan RRID:SCR_006849) and prioritized for inclusion if detected by ≥2 callers with a tumor allele frequency (AF) ≥5-fold that of the matched germline sample. The exome variant lists for the 56 patients were then split across six total personalized panels, such that each patient’s study samples were captured using a shared panel that included both their personalized variant list alongside those of other unrelated patients, allowing for cross-monitoring of patient-specific variants for downstream filtering. Each personalized SNV was tiled with a 120 bp biotinylated custom oligonucleotide (IDT) designed to cover the coordinates of the identified mutation.

### DNA library preparation and sequencing

Barcoded DNA sequencing libraries were prepared with unique molecular identifiers as previously described using KAPA HyperPlus Kits (Roche, RRID:SCR_000593)^38^. All patient samples underwent hybrid capture using biotinylated oligonucleotides targeting selected regions of the hg19 reference genome (IDT), including the canonical AML panel along with the personalized panel specific to each patient derived from WES. Hybrid capture was performed as previously described^38^ using the xGen Hybridization and Wash v2 Kit (IDT, RRID:SCR_011151). Libraries were sequenced on Illumina HiSeq4000 or NovaSeqXPlus instruments (Illumina; RRID:SCR_016387) with 2 x 150-bp paired-end reads.

### Sequencing data analysis and variant calling

Raw FASTQ files were demultiplexed using 8-bp dual sample barcodes and reads were mapped to the human reference genome (hg19) using Burrows-Wheeler Alignment (BWA) ALN (RRID:SCR_010910). Molecules were deduplicated with error suppression using a custom pipeline as previously described^11,38^, with samples excluded if their deduplicated depth was below 50x. Variant calling was performed using an adaptive variant calling strategy that considers the specific base change observed, the local depth, and both global and local error rates, as previously described^11^. Somatic variant calls were further filtered using the following heuristics to increase specificity: tumor VAF >5x matched germline (T cell) AF, and present in <10% of a panel of controls consisting of N=21 unrelated healthy controls.

### AML-CAPP-Seq MRD criteria

For each longitudinal sample, MRD detection was performed using the complete set of tumor-specific variants identified for that patient at diagnosis, including variants derived from both the fixed canonical AML panel and personalized WES-informed selector. All variants were analyzed jointly using a previously established Monte Carlo-based MRD detection framework adapted from prior CAPP-Seq studies^11,12,22^.

#### Variant selection and background modeling

The tumor-specific variants identified at diagnosis by WES of bulk or flow-purified AML cells were required to be enriched in tumor (>5-fold) relative to matched CD3 T-cell DNA. Variants detected in paired T cells were considered germline or non–tumor/AML-specific and were excluded from MRD tracking. Mutations associated with clonal hematopoiesis (e.g., DNMT3A, TET2, ASXL1) were not excluded *a priori* unless present in the matched T-cell compartment. Background error rates were empirically estimated for each base substitution using sequencing data from a panel of 21 unrelated healthy controls processed with identical laboratory and bioinformatic pipelines.

#### Monte Carlo–based MRD detection index

For each sample, observed mutant reads across all tracked variants were integrated into a single sample-level detection statistic^11,12,39^. Briefly, Monte Carlo simulations were used to model the expected distribution of mutant read counts under a tumor-free null hypothesis, accounting for locus-specific sequencing depth and empirically estimated base-specific error rates. The probability of observing the measured aggregate mutant signal under the null distribution was calculated to generate a patient-and sample-specific MRD detection p-value.

#### Calibration of MRD positivity and false-positive rate

To control false-positive detection, the same variant sets were evaluated in the healthy control cohort, and the false-positive rate (FPR) was defined as:

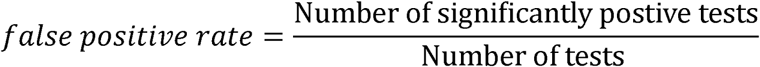

The MRD positivity threshold was then calibrated such that the overall FPR was maintained below 5%. Samples exceeding this calibrated threshold were classified as MRD-positive, whereas those below the threshold were classified as MRD-negative. As a consequence of this framework, MRD positivity thresholds and effective limits of detection are inherently patient-specific, depending on the number, identity, and sequencing depth of tracked variants.

#### Continuous ctDNA burden metrics

In addition to binary MRD classification, ctDNA burden was quantified using continuous measures, including mean tumor allele fraction (mean AF) across all tracked variants and longitudinal variant allele frequency trajectories. These continuous metrics were used to assess MRD depth, clonal dynamics, and temporal changes in disease burden.

#### Theoretical limits of detection (LOD)

Patient-specific theoretical limits of detection (LOD) for AML-CAPP-Seq-based MRD were estimated based on sequencing depth, number of tracked variants, and desired detection probability^39^. Assuming a mean deduplicated sequencing depth of approximately 2,500× per locus, a detection probability of 95%, and independent sampling across variants, the probability of observing at least one mutant molecule was modeled as a binomial process across all tracked tumor-specific SNVs. Under this framework, the per-patient LOD scales inversely with the number of independent tumor-specific variants available for tracking, resulting in a distribution of achievable sensitivities across patients rather than a single fixed threshold.

### Standard-of-Care Molecular and MRD Assessment

As part of routine clinical care, clinical NGS for diagnostic genotyping and subsequent molecular assessment was performed using the CLIA-certified HemeSTAMP assay (Stanford Clinical Labs). Conventional cytogenetic analyses, including karyotyping and fluorescence in situ hybridization (FISH) when indicated, were also performed using CLIA-validated assays (Stanford Clinical Labs). SOC MRD testing was performed using 10-color multiparameter flow cytometry (Stanford Clinical Labs; sensitivity 0.05–0.1%) and molecular assays (PCR or NGS) for NPM1c (Invivoscribe, San Diego, RRID:SCR_018209), FLT3-ITD (Invivoscribe), CBFB-MYH11 (Arup, Salt Lake City), or RUNX1-RUNX1T1 (Arup) when applicable. Based on manufacturer specifications and established clinical validation, the reported analytical sensitivity of these molecular assays is approximately 10 under optimized conditions.

Because approved molecular MRD assays are available for only a subset of AML genotypes, molecular MRD testing was not uniformly applicable across the cohort, whereas MFC was applied longitudinally to all patients. Accordingly, for comparative analyses, we defined a pragmatic “SOC combined MRD” comparator reflecting routine AML care. This composite SOC MRD assessment consisted of MFC for all patients, supplemented by single-variant molecular MRD results when such assays were clinically available and performed. In patients with approved molecular MRD markers, molecular results were incorporated alongside contemporaneous MFC findings; in patients without such markers, MFC alone constituted the SOC MRD assessment. This approach was used to enable a clinically relevant, real-world comparison between personalized ctDNA-MRD and standard MRD practices across the full AML cohort.

### Chimerism Assessment

AML-CAPP-Seq capture panels (canonical and personalized) were designed to target tumor-specific somatic mutations while also capturing germline single nucleotide polymorphisms (SNPs) within the covered regions. For post-transplant samples, donor-derived germline SNPs were identified by comparing variant allele frequencies (VAFs) across serial timepoints, particularly during clinical remission, to distinguish donor versus recipient alleles. Fractional estimates of donor-recipient chimerism were determined by calculating the quantitative proportion of donor-matching SNPs within cfDNA or leukocyte-derived DNA (BMMC/PBMC) to the recipient-derived tumor fraction in each sample. For comparison, standard-of-care short tandem repeat (STR) chimerism testing was performed at Stanford Clinical Laboratories on peripheral whole blood and leukocyte subsets, typically at Day +30, Day +90, and 1 year post-transplant, as part of routine clinical follow-up.

### Pathogenicity Prediction Analysis

To assess the pathogenicity of tumor SNVs (comparing variants located within canonical AML genes to those in noncanonical genes), we retrieved CADD scaled C-scores^19,20^, PHAST phastCons scores^15^, Zoonomia phyloP scores^16^, SIFT scores^17^, and ESM-1v log-likelihood probabilities^18^ for each variant from the Combined Annotation Dependent Depletion (CADD) database (RRID:SCR_018393). Zoonomia phyloP scores were subjected to negative log transformation, while ESM-1v log-likelihood probabilities were unsigned and then log transformed.

### Clonal Dynamics and Phylogenetic Inference

For selected cases with serial sampling, clonal architecture was reconstructed using VAF trajectories and variant co-occurrence patterns. Putative subclones were inferred and annotated based on diagnostic and relapse genotypes, and phylogenies were visualized to track clonal shifts over time, emergence of resistance mutations, and persistence of founder clones.

For selected patients with serial sampling, clonal architecture and phylogenetic relationships were inferred from variant allele frequency (VAF) trajectories and co-occurrence patterns. In most cases, PB-ctDNA samples were used for clonal analysis; alternate compartments were used when necessary based on clinical context and variant availability. For most samples, only variants with VAF ≥1% were included, unless clinically reported mutations were present below this threshold.

Filtered variant sets were clustered within each sample using the *SciClone* R package^33^ (RRID:SCR_013999), assuming diploid copy number across all loci. SciClone was run with a minimum read depth of 10 and a maximum number of clusters set to the lesser of 10 or the total number of variants per sample. This generated timepoint-specific cluster assignments for each variant. To derive patient-level clonal structures, SciClone cluster assignments were joined across timepoints to form a variant-by-sample matrix. Latent class analysis was then performed on this matrix using the *diceR* R package^40^ to generate consensus clone assignments. Clonal phylogenies were inferred using *ClonEvol*^41^ (RRID:SCR_018512) assuming a polyclonal initiation model due to the high frequency of post-transplant cases. Cluster centers were calculated as the mean VAF of constituent variants, and the founding clone was defined as the cluster with the highest average VAF across all samples.

Following clonal inference, biological constraints were applied. Clones containing mutations considered to be mutually exclusive—such as multiple variants within the same gene—were subdivided into separate subclones. For each subdivided clone, VAFs were recalculated by scaling individual variant VAFs according to their contribution relative to the original cluster mean. Clonal evolution was visualized using the *fishplot* R package^42^.. For timepoints flanked by detectable disease but with mean VAF <1%, minimal VAF values were imputed to preserve clonal continuity in the visualization. Final fish plots were manually annotated and refined using vector-based image editing software (Inkscape, Adobe Illustrator).

### Statistical analysis

Analyses were performed using R v4.5.0 (RRID:SCR_001905) with RStudio v2025.05.0 (RRID:SCR_000432), MATLAB R2020b (RRID:SCR_001622). Unless otherwise specified, all statistical tests were two-sided with a significance threshold of *p* < 0.05.

#### Comparisons across compartments and timepoints

Paired comparisons of VAFs or sample-level mean tumor allele fractions between matched compartments (PB-ctDNA, BM-ctDNA, BMMCs, PBMCs) were evaluated using the paired Wilcoxon signed-rank test. For analyses requiring correlation between continuous measures across paired samples, association was assessed using Spearman’s rank correlation (ρ). Correlation significance was computed using standard *t*-based tests.

#### Time-to-event analyses

Kaplan–Meier estimates were generated and compared between groups using the log-rank test. Effect sizes were summarized using hazard ratios (HRs) with 95% confidence intervals (CIs) derived from Cox proportional hazards regression models. In settings with small sample sizes, low event counts, or complete or near-complete separation between risk groups, Firth penalized Cox regression was used to reduce small-sample bias and ensure stable estimation of effect sizes. For relapse-free survival (RFS) analyses, patients were stratified by MRD status at the most recent evaluable pre-relapse timepoint (for relapsed patients) or at last available follow-up (for non-relapsed patients), as described in the Results. Overall survival (OS) analyses stratified patients by achievement of MRD negativity at any timepoint unless otherwise specified. Prespecified subgroup analyses included stratification by ELN 2022 risk category, treatment intensity, and availability of approved single-variant molecular MRD markers.

#### Comparative prognostic performance

To compare MRD strategies (SOC MFC, SOC combined, ctDNA using canonical variants only, and ctDNA using all personalized variants), we evaluated model discrimination using Harrell’s concordance index (C-index) and model fit using likelihood ratio statistics and Akaike Information Criterion (AIC), reporting ΔAIC relative to the best-performing model within each endpoint.

#### Chimerism analyses

Quantitative concordance between cfDNA-derived and STR-derived donor chimerism estimates was assessed using Spearman correlation. Categorical concordance at clinically relevant thresholds was evaluated using Fisher’s exact test.

#### Reproducibility and code availability

All statistical analyses and figure-generation code (including preprocessing, modeling, and plotting scripts) will be made publicly available in a Code Ocean capsule prior to publication; this capsule will enable full computational reproducibility of all main and supplementary results.

## Data availability

Genotypic data generated in this study will be deposited in Figshare and made publicly available upon publication. Due to patient confidentiality and restrictions in the informed consent approved by the Stanford Institutional Review Board, raw sequencing data are considered protected health information and are not publicly available. Additional processed data and analysis outputs are available from the corresponding author upon reasonable request. For inquiries, please contact the corresponding author at.

## Acknowledgments

This work was supported by the National Institutes of Health (K08CA248940 to T.Y.Z.; K08CA241076 to D.M.K.) and the Stanford Cancer Institute (T.Y.Z., D.M.K.). R.G. has received support from the Stanford Division of Hematology (T32 HL120824, Dr. Steven Coutre Fellowship), the American Society of Hematology (Research Training Award for Fellows), the Stanford Cancer Institute Fellowship, and the Association for Northern California Oncologists. We thank Hitomi Hosoya, Asiri Ediriwickrema, Brooks Benard, and Jason Gotlib for technical and/or manuscript advice.

## Authors’ Contributions

R.G., D.M.K., and T.Y.Z. designed and conceptualized the study. K.T., C.T., G.N.M., and T.Y.Z. enrolled patients. K.T., C.T., C.Z., R.G., and E.Y. processed samples. R.G., K.T., C.T., C.Z., and T.Y.Z. curated clinical data. R.G., C.Z., G.G., R.Y., and M.C. performed experimental investigations. R.G., S.R., M.S.A., E.Y., B.J.S., and D.M.K. performed data analysis. M.S. and D.M.K. designed the canonical selector. J.W.T. and S.K. performed flow cytometry. M.S.K. and D.M.K. provided software support. R.M. provided technical and conceptual advice. R.G. and T.Y.Z. wrote the manuscript. S.R., E.Y., M.S.A., M.S., M.S.K., R.M., D.M.K., and T.Y.Z. critically reviewed and revised the manuscript. C.Z. and S.R. contributed equally to this work.

