## Supplemental Information File for "Personalized Circulating Tumor DNA (ctDNA) Profiling Enables Superior and Universal Measurable Residual Disease (MRD) Detection in Acute Myeloid Leukemia (AML)"

**Supplementary Tables**

**Table S1:** Genes and fusion regions covered by canonical AML-CAPP-Seq panel

| Genes | | | | | | | Fusions |
|---|---|---|---|---|---|---|---|
| *ARID1A* | *CDKN2A* | *FLT3* | *KIT* | *NRAS* | *SETD2* | *U2AF1* | *MYH11-INTRON-32* |
| *ARID2* | *CEBPA* | *GATA2* | *KMT2A* | *PHF6* | *SF3B1* | *U2AF2* | *KMT2A-INTRON-7* |
| *ASXL1* | *CREBBP* | *GNAS* | *KMT2D* | *PPM1D* | *SMC1A* | *WT1* | *KMT2A-INTRON-8* |
| *BCOR* | *CTCF* | *HNRNPK* | *KRAS* | *PTEN* | *SMC3* | *ZRSR2* | *KMT2A-INTRON-9* |
| *BCORL1* | *DDX41* | *IDH1* | *MED12* | *PTPN11* | *SRSF2* | | *KMT2A-INTRON-10* |
| *BRAF* | *DNMT3A* | *IDH2* | *MPL* | *RAD21* | *STAG2* | | |
| *BRINP3* | *EED* | *IKZF1* | *MYC* | *RET* | *SUZ12* | | |
| *CALR* | *ETV6* | *JAK2* | *NF1* | *RUNX1* | *TET2* | | |
| *CBL* | *EZH2* | *JAK3* | *NPM1* | *SETBP1* | *TP53* | | |

### Table S2. Clinical characteristics of patients with newly diagnosed AML at study enrollment.

| Patient ID | Gender | AML Type | ELN Risk | Induction Intensity | Induction Treatment | Consolidation Treatment | Achieved cCR1 | HSCT in cCR1 | Relapse in cCR1 | Achieved cCR2 | HSCT in cCR2 | Relapse in cCR2 | Vital status |
|---|---|---|---|---|---|---|---|---|---|---|---|---|---|
| AML002 | M | De novo | Adverse | Low | Dec+Ven | Dec+Ven | Yes | No | Yes | No | NA | NA | Dead |
| AML003 | F | De novo | Adverse | High | 7+3 | IDAC | Yes | Yes | Yes* | No | NA | NA | Dead |
| AML004 | M | De novo | Adverse | Low | Dec+Ven | Dec+Ven | Yes | No | Yes | No | NA | NA | Dead |
| AML005 | M | Secondary (JAK2+ MPN) | Adverse | High | FLAG+IDA | IDAC+GO | Yes | No | Yes | Yes | Yes | Yes* | Dead |
| AML007 | M | De novo | Favorable | High | 7+3 | IDAC | Yes | No | No | NA | NA | NA | Alive |
| AML008 | M | De novo | Adverse | High | 7+3 | Dec+Ven | Yes | Yes | No | NA | NA | NA | Alive |
| AML009 | F | De novo | Intermediate | Low | Dec+Ven | Dec+Ven | Yes | No | Yes | No | NA | NA | Dead |
| AML012 | F | De novo | Intermediate | High | CPX-351+mido | Dec+Gilt | Yes | Yes | No | NA | NA | NA | Alive |
| AML019 | M | Therapy-related (Hodgkin lymphoma) | Adverse | Low | Dec+Ven | Dec+Ven | Yes | Yes | Yes* | No | NA | NA | Dead |
| AML020 | M | De novo | Adverse | High | FLAG+Ida+Ven | FLAG+Ida+Ven | Yes | No | Yes | No | NA | NA | Dead |
| AML023 | M | Secondary (MDS) | Adverse | Low | ASTX-727+Ven | ASTX-727+Ven | Yes | Yes | No | NA | NA | NA | Alive |
| AML028 | M | De novo | Adverse | High | FLAG+Ida+Ven | FLAG+Ven | Yes | No | Yes | No | NA | NA | Dead |
| AML029 | M | De novo | Adverse | High | 7+3 | Dec+Ven | Yes | Yes | Yes* | Yes | No | NA | Alive |
| AML033 | F | De novo | Intermediate | High | FLAG+Ida+Ven | FLAG+Ida+Ven | Yes | Yes | No | NA | NA | NA | Alive |
| AML041 | M | De novo | Favorable | High | 7+3 | IDAC | Yes | Yes | Yes | Yes | Yes | No | Alive |
| AML047 | M | De novo | Adverse | Low | Dec+Ven | Dec+Ven | Yes | No | Yes | No | NA | NA | Dead |
| AML054 | M | De novo | Adverse | High | FLAG+Ida+Ven | FLAG+Ida+Ven | Yes | Yes | Yes* | No | NA | NA | Dead |
| AML058 | F | Therapy-related (colorectal cancer) | Adverse | High | FLAG+Ida+Ven | NA | Yes | No | Yes | No | NA | NA | Dead |
| AML059 | M | De novo | Intermediate | High | 7+3+Mido | IDAC | Yes | No | Yes | Yes | Yes | No | Alive |
| AML067 | M | De novo | Adverse | Low | Dec+Ven | Dec+Ven | Yes | NA | Yes | No | NA | NA | Dead |
| AML068 | F | De novo | Adverse | High | 7+3+Mido | Gilt (later Dec+Ven) | Yes | Yes | No | NA | NA | NA | Alive |
| AML071 | F | De novo | Favorable | High | 7+3+Mido | IDAC | Yes | No | Yes | Yes | Yes | No | Alive |
| AML077 | F | Therapy-related (DLBCL) | Intermediate | High | 7+3+mido | Gilt | Yes | Yes | Yes* | Yes | No | No | Alive |
| AML078 | M | De novo | Favorable | High | 7+3+Mido | IDAC | Yes | No | Yes | No | No | Yes | Dead |
| AML080 | M | Secondary (ET) | Adverse | Low | Dec+Ven | Dec+Ven | No | NA | NA | NA | NA | NA | Dead |
| AML086 | M | De novo | Intermediate | High | 7+3 | IDAC | Yes | No | Yes | Yes | Yes | No | Alive |
| AML093 | F | De novo | Favorable | High | 7+3+GO | IDAC | Yes | No | No | NA | NA | NA | Alive |
| AML103 | F | Therapy-related (peritoneal cancer) | Adverse | Low | Aza+Ven | Aza+Ven | No | NA | NA | NA | NA | NA | Dead |
| AML109 | M | De novo | Adverse | Low | Dec+Ven | Dec+Ven | Yes | Yes | Yes* | No | NA | NA | Dead |
| AML113 | F | De novo | Adverse | High | Clad+LDAC+Ven | Clad+LDAC+Ven | Yes | Yes | No | NA | NA | NA | Alive |
| AML117 | M | De novo | Intermediate | High | 7+3+Mido | IDAC+Mido | Yes | Yes | No | NA | NA | NA | Alive |
| AML118 | F | Therapy-related (breast cancer) | Adverse | High | FLAG+Ida+Ven | FLAG+Ida+Ven | Yes | Yes | No | NA | NA | NA | Alive |
| AML122 | M | De novo | Intermediate | High | Clad+LDAC+Ven | Clad+LDAC+Gilt | Yes | Yes | No | NA | NA | NA | Alive |
| AML131 | M | Secondary (PV) | Adverse | High | Clad+LDAC+Ven | Clad+LDAC+Ven | Yes | No | Yes | No | NA | NA | Dead |
| AML138 | M | Therapy-related (breast cancer) | Intermediate | High | 7+3+Mido | IDAC+Mido | Yes | Yes | Yes* | Yes | No | No | Alive |
| AML142 | M | De novo | Adverse | High | 7+3 | Dec+Ven | Yes | Yes | No | NA | NA | NA | Alive |
| AML145 | F | De novo | Intermediate | High | 7+3 | Dec+Ven | Yes | No | Yes | Yes | Yes | No | Alive |
| AML146 | F | De novo | Favorable | Low | Dec+Ven | Dec+Ven | No | NA | NA | NA | NA | NA | Dead |
| AML147 | F | De novo | Intermediate | High | FLAG+Ida+Ven | FLAG+Ida+Ven | Yes | Yes | No | NA | NA | NA | Alive |
| AML149 | M | De novo | Favorable | High | 7+3+GO | IDAC+GO | Yes | No | Yes | No | NA | NA | Dead |
| AML151 | M | De novo | Adverse | Low | Dec+Ven | Dec+Ven | Yes | No | Yes | No | NA | NA | Dead |
| AML152 | M | De novo | Intermediate | High | Clad+LDAC+Ven | Clad+LDAC+Ven | Yes | Yes | No | NA | NA | NA | Alive |
| AML155 | F | De novo | Favorable | High | 7+3 | IDAC | Yes | No | Yes* | Yes | Yes | Yes* | Dead |
| AML156 | F | De novo | Favorable | High | Clad+LDAC+Ven | IDAC | Yes | No | No | NA | NA | NA | Alive |
| AML162 | M | De novo | Favorable | Low | Aza+Ven | Aza+Ven | Yes | No | Yes | No | NA | NA | Dead |
| AML163 | F | De novo | Adverse | High | Clad+LDAC+Ven | Dec+Ven | Yes | No | Yes | No | No | No | Dead |
| AML165 | F | De novo | Adverse | High | 7+3+GO | Dec+Ven | Yes | Yes | Yes* | No | NA | NA | Dead |
| AML167 | M | De novo | Favorable | High | 7+3 | IDAC | Yes | No | Yes | No | NA | NA | Dead |
| AML169 | M | De novo | Favorable | High | 7+3+GO | HIDAC | Yes | No | No | NA | NA | NA | Alive |
| AML171 | M | De novo | Adverse | Low | Dec | Dec | Yes | No | Yes | No | NA | NA | Dead |

This table summarizes baseline demographic, diagnostic, treatment, and outcome characteristics for patients with newly diagnosed AML (n = 50) included in the study. Variables shown include patient identifier, sex, leukemia type (de novo, secondary, or therapy-related), European LeukemiaNet (ELN) risk category, induction treatment regimen and intensity, consolidation therapy, achievement of first composite clinical remission (cCR1), receipt of allogeneic HSCT in cCR1, and relapse status during cCR1. Among those who relapsed in cCR1, additional outcome variables include achievement of second composite clinical remission (cCR2), receipt of allogeneic HSCT in cCR2, and relapse status during cCR2. ELN risk groups were assigned according to 2022 criteria. Current vital status is also reported. * denotes relapse after allogeneic HSCT.

**Abbreviations**: 7+3, standard induction consisting of 7 days of cytarabine plus 3 days of an anthracycline; Dec, decitabine; Aza, azacitidine; Ven, venetoclax; Mido, midostaurin; Gilt, gilteritinib; Ida, idarubicin; Clad, cladribine; LDAC, low-dose cytarabine; IDAC, intermediate-dose cytarabine; HIDAC, high-dose cytarabine; GO, gemtuzumab ozogamicin; FLAG, fludarabine, cytarabine, and granulocyte colony-stimulating factor; Magro, magrolimab; MEC, mitoxantrone, etoposide, and cytarabine.

**Table S3. Clinical characteristics of patients with relapsed or refractory (R/R) AML at study enrollment.** This table summarizes demographic, disease, treatment, and outcome characteristics for patients with R/R AML (n = 6) at the time of study enrollment. Variables shown include patient identifier, sex, leukemia type (de novo, secondary, or therapy-related), European LeukemiaNet 2022 risk category, prior therapies, re-induction treatment regimen and intensity, achievement of first composite clinical remission (cCR1) after study enrollment, receipt of allogeneic HSCT in cCR1, and relapse status during cCR1. Among patients who achieved cCR1 and subsequently relapsed, additional outcome variables include achievement of second composite clinical remission (cCR2), receipt of allogeneic HSCT in cCR2, and relapse status during cCR2. Current vital status is also reported.

**Abbreviations**: 7+3, standard induction consisting of 7 days of cytarabine plus 3 days of an anthracycline; Dec, decitabine; Aza, azacitidine; Ven, venetoclax; Mido, midostaurin; Gilt, gilteritinib; Ida, idarubicin; Clad, cladribine; LDAC, low-dose cytarabine; IDAC, intermediate-dose cytarabine; HIDAC, high-dose cytarabine; GO, gemtuzumab ozogamicin; FLAG, fludarabine, cytarabine, and granulocyte colony-stimulating factor; Magro, magrolimab; MEC, mitoxantrone, etoposide, and cytarabine.

| Patient ID | Gender | AML Type | ELN Risk | Prior Treatments | Re-induction Treatment | Re-induction Intensity | Achieved cCR1 | HSCT in cCR1 | Relapse in cCR1 | Achieved cCR2 | HSCT in cCR2 | Relapse in cCR2 | Vital status |
|---|---|---|---|---|---|---|---|---|---|---|---|---|---|
| AML044 | M | De novo | Adverse | CPX-351 | Clad+LDAC+Ven | High | No | No | NA | NA | NA | NA | Dead |
| AML061 | M | De novo | Adverse | 7+3, Clad+LDAC+Ven | SNDX-5613 | Low | Yes | No | Yes | Yes | No | Yes | Dead |
| AML070 | M | De novo | Adverse | 7+3 | Clad+LDAC+Ven | High | No | NA | NA | NA | NA | NA | Dead |
| AML087 | M | De novo | Favorable | 7+3+GO | MEC+Magro | High | Yes | Yes | No | NA | NA | NA | Alive |
| AML099 | F | De novo | Adverse | 7+3, Dec+Ven | MEC+Magro | High | No | NA | NA | NA | NA | NA | Dead |
| AML141 | M | De novo | Favorable | 7+3+GO | FLAG+Ida+Ven | High | Yes | Yes | No | NA | NA | NA | Dead |

**Table S4. Diagnostic genomic characteristics of patients with newly diagnosed AML at study enrollment.** This table summarizes genomic and molecular features identified at time of diagnosis for patients with newly diagnosed AML (n = 50) included in the study. Variables shown include patient identifier, European LeukemiaNet 2022 risk category, availability and type of standard-of-care (SOC) molecular MRD assay, pathogenic somatic mutations detected by clinical NGS at diagnosis (with gene name, amino acid change, and variant allele fraction if available), and cytogenetic abnormalities identified by conventional karyotyping and/or fluorescence in situ hybridization. Cytogenetic findings are reported using standard International System for Human Cytogenomic Nomenclature conventions. * denotes nonsense mutation.

| Patient ID | ELN Risk | SOC Molecular MRD Assay | Clinical NGS at Dx (Pathogenic Variants) | Cytogenetics at Dx |
|---|---|---|---|---|
| AML002 | Adverse | NA | DNMT3A D472fs 52%, TET2 Q108fs 48%, U2AF1 S34F 43%, PTPN11 E76G 27%, NRAS Q61H 15%, FLT3 D835Y 2% | 47,XY, del(9)(q12q22),+11 |
| AML003 | Adverse | NA | KMT2A-MLLT10 fusion, SIK3-KMT2A fusion | 46,XX[20] |
| AML004 | Adverse | NA | DNMT3A R882H 46%, EZH2 L240fs 28%, KRAS S65delinsTPR 4%, SF3B1 K666N 44%, TET2 E576X 7% | 46, XY, add(11)(p11.2)[10]/46,XY[10] |
| AML005 | Adverse | NA | JAK2 V617F 4%, KRAS K117N 21%, NRAS G13C 47%, TET2 R1216X 46%, U2AF1 S34F 42% | 46, XY[20] |
| AML007 | Favorable | NA | CEBPA G38fs 49%, GATA2 A318T 49%, NRAS G12D 7%, CEBPA Q311_Q312insPRQRNVETQ 11%, KRAS G12D 3%, NRAS Q61H 2% | 46,XY[20] |
| AML008 | Adverse | NA | SRSF2 P95_R102del (27%), TP53 L252_I254del (75%), STAG2 E265X (44%) | 47,XY,add(3) (p13),-5,+6,+8,add(15) (p11.2) [cp14]/47,idem,del (12)(p11.20[2]/49,idem,+1.+2[cp3] / 46,XY[1] |
| AML009 | Intermediate | FLT3-ITD, NPM1 | NPM1 W288Fs (43%), FLT3-ITD , WT1 A382fs | 46,XX +8,t(15;17) (q15;q11) [16]/46,idem,+8[1] / 46,XX[3] |
| AML012 | Intermediate | FLT3-ITD | FLT3 N676K (43%), FLT3-ITD | 46, XX[10] |
| AML019 | Adverse | NA | TP53 M237I 86% | 43~44,XY,add(1)(p?13~p22),del(5)(q13q33),+6,add(6)(q12),-7,del(12)(p11.2),add(20)(q?11.2)[cp2]/45,XY,add(1)(p13),del(5)(q13q33),+6,add(6)(q12)x2,-7,-8,add(11)(q25),del(12)(p11.2), add(20)(q?11.2)[18]/46,XY[1].ish add(11)(q25)(RUNX1T1+++,KMT2A+++) |
| AML020 | Adverse | NA | ASXL1 G645fs (x2) (27%) (10%), NRAS G12D (48%), RUNX1 R107C (19%), RUNX1 A142fs (26%) | 46,XY,inv(3) (q21q26.2) [21] |
| AML023 | Adverse | NPM1 | CBL C384R (44%), IDH1 R132C (51%), NPM1 W288fs (41%), SRSF2 P95L (4%) | 46,XY[21] |
| AML028 | Adverse | NA | TP53 L252del (9%) | 43,XY,del(3)(p21),-5,add(5)(p15),-7,-9,add(9)(p24),add(11)(p15),-14,-18,+2mar[1]/46,XY[2] |
| AML029 | Adverse | NA | CEBPA G53fs 52%, CEBPA Q312dup 44% | 46,XY,i(17),(q10)[17]/46,XY[3] |
| AML033 | Intermediate | NA | FLT3 I836del 27%, PTPN11 A72V 4%, KMT2A:MLLT3 fusion | 46,XX,t(9;11)(p22;q23)[20] |
| AML041 | Favorable | CBFB:MYH11 | NRAS G12D (3%), CBFB:MYH11 fusion | 46, XY[20] |
| AML047 | Adverse | NA | TERT c. -146C>T Promoter mutation 47% | 46, XY, t(8;16)(p11.2;p13.3)[17]/46,XY[3] |
| AML054 | Adverse | NA | NF1 I679fs 17%, TP53 L257Q 78% | 42~43,X,-Y,add(3)(q12),del(4)(q13),add(5)(q12),-6,der(7;14)(p10;q10),+11,-12,-17,+mar[cp20]/46,XY[1] |
| AML058 | Adverse | FLT3-ITD | DNMT3A R882H (1%), FLT3-TKD E598_Y599insRT (4%), TP53 D184fs (45%), FLT3-ITD | 48~52, XX,-5,del(5)(q22q35),der(11)del(11)(p13p11.2)del(11)(q13q23),+der(11)x2~4,add(13)(q22),-16,+19,+22,+1~2mar[cp20 |
| AML059 | Intermediate | NA | DNMT3A K577* (6%), FLT3-TKD N676K (2%), KDM6A D1069fs (75%) | 46,XY[19] |
| AML067 | Adverse | NA | TP53 Q331fs 46%, TP53 P151T 37%, | 45~49,XY,-5,der(7)add(7)(p12)add(7)(q32),add(8)(q22),-16,add(17)(p12),-18,+idic(22)(q12), +1~6mar[cp20] |
| AML068 | Adverse | NA | NRAS G12S 3%, FLT3-TKD D835V 2%, RUNX1 R201G 48%, ASXL1 G646fs 44%, PHF6 c.834+1G>T 16% | 47,XX,+22[9]/50,idem,+9,+13,+21[7]/46,XX[4] |
| AML071 | Favorable | NPM1 | DNMT3A R882C 41%, NPM1 W288fs 30%, FLT3-TKD D835V 19% | 46,XX[20] |
| AML077 | Intermediate | FLT3-ITD | CSF3R W547* (48%), FLT3 W599_E604dup, IDH2 R140Q (1%), FLT3-ITD | 46,XX,del(9)(q13q22)[22] |
| AML078 | Favorable | CBFB:MYH11 | FLT3 D835H (6%), FLT3 D835V (6%), KRAS G13D (2%), NRS G13D (2%), NRAS Q61R (2%), KRAS G12A (1%), CBFB:MYH11 fusion, ASXL1 T655fs | 46, XY [20] |
| AML080 | Adverse | NA | NF1 L2158* 56%, RUNX1 D198G 40%, ETV6 S131fs 44% | 45, XY, -7[21] |
| AML086 | Intermediate | NA | NA | 46,XY[20] |
| AML093 | Favorable | NPM1 | NPM1 W288fs 28%, STAG2 G313fs 22%, PHF6 Y103* 3%, TET2 S167fs 2% | 46 XX |
| AML103 | Adverse | NA | TP53 Y220C 96%, NRAS Q61L 38% | 42~45,XX,-3,del(4)(q25q35),del(5)(q31q35),-7,-11,add(12)(p11.2),-13,?der(13;17)(q10;q10), -21,add(21)(p11.2),-22,+2~6mar[cp9]/46,XX[1] |
| AML109 | Adverse | NA | ASXL1 V515fs (44%), U2AF1 Q157P (49%), SETBP1 D868N (46%) | 46,XY[15] |
| AML113 | Adverse | NA | ASXL1 G646fs 48%), CEBPA V159fs*11 (58%), KRAS G13D (22%), NRAS G12V (5%), NRAS A146T (5%), SRSF2 P95H (49%), TET2 R1465* (96%) | 46,XX[20] |
| AML117 | Intermediate | NPM1, FLT3-ITD | DNMT3A R882H (46%), NPM1 W288fs (55%), FLT3-ITD, KRAS G13D (22%), PTPN11 A72T (2%) | 46,XY[20] |
| AML118 | Adverse | NA | CCND3 R271fs, KRAS G12V, NRAS G13R, KMT2A:ELL rearrangement | 46,XX,r(6)(p23q25),t(11;19)(q23;p13.?.1)[21] |
| AML122 | Intermediate | FLT3-ITD | TET2 Q847* (49%), RAD21 (45%), FLT3-ITD, RUNX1:intergenic fusion | 46,XY,t(1;21)(p13;q22.1)[2]/46,XY[9] |
| AML131 | Adverse | NA | RUNX1 (59%), JAK2 V617F (93%), PPM1D N512fs (43%) | 46,XY,del(20)(q11.2q13.1)[5]/46,XY[18] |
| AML138 | Intermediate | FLT3-ITD | CEBPA E309_T310insGPALA (35%), CEBPA A91fs (48%) , FBXW7 R505L (48%), FLT3-ITD | 46,XY[20] |
| AML142 | Adverse | NA | ASXL1 G646fs (44%), IDH2 R140Q (48%), RUNX1 P294fs (4%), SRSF2 P95H (46%), STAG2 F468fs (85%) | 46,XY[18] |
| AML145 | Intermediate | NA | TET2 S1525fs (4%), TET2 E1492fs (4%), WT1 Q142 (6%) | 46,XX[19] |
| AML146 | Favorable | NA | ASXL1 G646fs (39%), CEBPA L338P (40%), CEBPA H24fs (35%), TET2 C1271fs (88%) | 46,XX[20] |
| AML147 | Intermediate | NA | FLT3 D835Y (30%), FLT3 D835E (4%), KMT2A:MLLT3 rearrangement | 46,XX,t(9;11)(p22;q23)[cp9]/46,XX[3] |
| AML149 | Favorable | RUNX1-RUNX1T1 | RUNX1:RUNX1T1 fusion, KIT D816V (12%), NRAS Q61K (3%) | 45,X.-Y,t(8;21)(q22;q22)[20];t(t;21)(q22;q22);RUNX1::RUNX1T1 |
| AML151 | Adverse | NA | ATM L2850 (44%), FLT3 D835Y (7%), FLT3-TKD R834_D835del (3%), RUNX1 R320fs (38%), SF3B1 K666N (48%) | 46,XY[19] |
| AML152 | Intermediate | NA | KMT2A:MLLT3 fusion | 46,XX[20] |
| AML155 | Favorable | NA | CEBPA P23fs (42%), CEBPA K313dup (42%), EP300 W1681* (14%) | 46,XX[20] |
| AML156 | Favorable | NA | CEBPA Y108fs (58%), CEBPA Q311P (37%), TET2 R1216 (38%) | 46,XX,del(9)(q?13q?22)[2]/46,XX[20] |
| AML162 | Favorable | NPM1 | NPM1 W288fs (34%), FLT3-TKD D835H (3%), FLT3-TKD D835E (2%), TET2 (36%), FLT3 N831T (27%) | 45,X,-Y[15]/46,XY[5] |
| AML163 | Adverse | NA | DNMT3A W327* (42%), IDH2 R140Q (44%), NRAS G13R (34%), TP53 R248W (95%) | 48,XX,+8,+21[20] |
| AML165 | Adverse | NA | GATA2 G320D (20%), IKZF1 N159S (28%), NF1 c.7322-1G>A (29%), RUNX1 E80fs (43%), SF3B1 K700E (35%) | 46,XX,t(3;3)(q21;q26.2)[19]/46,XX[1 |
| AML167 | Favorable | NPM1 | NPM1 W288fs (35%), KRAS G13D (4%), RAD21 G547fs (39%), DNMT3A V716D (42%), FBXW7 R689W (40%), DNMT3A V716D (42%) | 46,XY |
| AML169 | Favorable | CBFB:MYH11 | KIT D816H (22%), NRAS G13D (11%), NRAS Q61R (4%), CBFB:MYH11 fusion | 46,XY,t(16;16)(p13;q22)[6]/47,idem,+8[15] |
| AML171 | Adverse | NA | TP53 R273H (44%), TP53 V173M (45%) | 54,XY,+1,+5,del(5)(q13q35)x2,+6,+8,+11,+18,+19,+21,add(22)(q13)[19]/46,XY[1] |

**Table S5. Genomic characteristics of patients with relapsed or refractory (R/R) AML at study enrollment.** This table summarizes genomic and molecular features identified at the time of study enrollment for patients with R/R AML (n = 6). Variables shown include patient identifier, European LeukemiaNet 2022 risk category, availability and type of standard-of-care (SOC) molecular MRD assay, pathogenic somatic mutations detected by clinical NGS at study enrollment (with gene name, amino acid change, and variant allele fraction if available), and cytogenetic abnormalities identified by conventional karyotyping and/or fluorescence in situ hybridization at study enrollment. Cytogenetic findings are reported using standard International System for Human Cytogenomic Nomenclature conventions. * denotes nonsense mutation.

| Patient ID | ELN Risk | SOC Molecular MRD Assay | Clinical NGS (Pathogenic Variants) | Cytogenetics |
|---|---|---|---|---|
| AML044 | Adverse | NA | ASXL1 C594* (42%), IDH1 R132L (17%), PHF6 c.730-1G>A (32%), IDH2 R172K (8%), PHF6 R342X (12%), U2AF1 Y158_E159dup (37%) | 45,XY,-3,dic(5;17)(q11.2~13;p11.2~13),del(7)(q31q?35),+8[cp21] |
| AML061 | Adverse | NA | RAD21 A21fs (2%), U2AF1 S34T (40%), KMT3A-MLLT3 fusion | 46,XY,t(2;19)(p21;q13.3),t(9;11)(p22;q23)[11]/46,idem,del(11)(q13q23)[2]/46,XY,t(1;10)(p13;q22),t(9;11)(p22;q23),?add(11)(p11.2)[3]/46,XY[4] |
| AML070 | Adverse | NA | CEBPA A111fs (36%), CSF3R T618I (31%), SETBP1 I871T (30%), PHF6 R116* (4%), SETBP1 D868N (4%), SRSF2 P95R (35%) | 46,XY[20] |
| AML087 | Favorable | NPM1 | NPM1 W288fs (38%), IDH2 R140L( 38%), CEBPA Y108* (32%), CEBPA P128S (44%) | 46,XY[20] |
| AML099 | Adverse | NA | PPM1D F445fs (5%), TP53 R273C (4%), DDX41 R525H (4%) | 46,XX[20] |
| AML141 | Favorable | CBFB:MYH11 | FLT3 D835V (35%), CBFB:MYH11 fusion | 46,XY,inv(16)(p13.1q22)[16]/46,idem,del(7)(q22q34)[cp5]/46,XY[2] |

**Table S6. Clinical and transplant characteristics of AML patients undergoing allogeneic hematopoietic stem cell transplantation (HSCT).** This table summarizes demographic, disease, and transplant-related characteristics for patients (n = 29) who underwent allogeneic HSCT during their treatment course. Variables shown include patient identifier, sex, AML subtype, European LeukemiaNet 2022 risk category, receipt of HSCT in first or second composite clinical remission (cCR), transplant donor type, conditioning regimen intensity (myeloablative or reduced intensity), preparative regimen, post-HSCT graft-versus-host disease prophylaxis, receipt of post-HSCT maintenance therapy, occurrence of graft-versus-host disease, relapse following HSCT, and vital status at last follow-up. Date of ctDNA MRD clearance indicates the timepoint at which personalized ctDNA–MRD first became undetectable following HSCT. Entries are reported as the post-HSCT day of initial ctDNA negativity, "Pre-HSCT" if ctDNA-MRD was already negative prior to transplantation, or "Never cleared" if ctDNA-MRD remained persistently detectable throughout post-HSCT follow-up.

**Abbreviations:** MRD, matched related donor; URD, unrelated donor; mmURD, mismatched unrelated donor; haplo, haploidentical donor; MAC, myeloablative conditioning; RIC, reduced intensity conditioning; Flu, fludarabine; Bu, busulfan; TT, thiotepa; Cy, cyclophosphamide; TBI, total body irradiation; Mel, melphalan; FK, tacrolimus; MTX, methotrexate; Siro, sirolimus.

| Patient ID | Gender | AML Subtype | ELN Risk | HSCT in cCR1 or cCR2 | Donor | HSCT Intensity | Preparative Regimen | Prophylaxis | Post-HSCT Maintenance | GVHD | Date of ctDNA MRD Clearance | Relapse Post-HSCT | Vital status |
|---|---|---|---|---|---|---|---|---|---|---|---|---|---|
| AML003 | F | De novo | Adverse | cCR1 | MRD | MAC | Flu/Bu/TT | FK | NA | No | Day 201 | Yes | Dead |
| AML005 | M | Secondary | Adverse | cCR2 | mmURD | MAC | Bu/Cy | FK/MTX | NA | No | Never cleared | Yes | Dead |
| AML008 | M | De novo | Adverse | cCR1 | MRD | MAC | Flu/Bu/TT | FK | NA | Yes | Day 89 | No | Alive |
| AML012 | F | De novo | Intermediate | cCR1 | MRD | MAC | Bu/Cy | FK/MTX | Gilteritinib | Yes | Day 87 | No | Alive |
| AML019 | M | Secondary | Adverse | cCR1 | MRD | MAC | Bu/Cy | FK/MTX | NA | Yes | Day 83 | Yes | Dead |
| AML023 | M | Secondary | Adverse | cCR1 | MUD | RIC | Flu/TT/TBI | FK | NA | No | Day 84 | No | Alive |
| AML029 | M | De novo | Adverse | cCR1 | MRD | MAC | Bu/Cy | FK/MTX | NA | Yes | Day 99 | Yes | Alive |
| AML033 | F | De novo | Intermediate | cCR1 | MUD | MAC | Bu/Cy | FK/MTX | NA | Yes | Pre-HSCT | No | Alive |
| AML041 | M | De novo | Favorable | cCR1 | MUD | MAC | Bu/Cy | FK/MTX | NA | No | Day 90 | No | Alive |
| AML054 | M | De novo | Adverse | cCR1 | mmURD | MAC | Flu/Bu | FK/MMF/Cy | NA | Yes | Day 87 | Yes | Dead |
| AML059 | M | De novo | Intermediate | cCR2 | MUD | MAC | Flu/Bu | FK/MMF/Cy | NA | Yes | Pre-HSCT | No | Alive |
| AML068 | F | De novo | Adverse | cCR1 | MUD | MAC | Flu/Bu/TT | FK | NA | Yes | Day 370 | No | Alive |
| AML071 | F | De novo | Favorable | cCR2 | MRD | MAC | Flu/Bu/TT | FK | Gilteritinib | No | Day 95 | No | Alive |
| AML077 | F | Therapy-related | Intermediate | cCR1 | MRD | MAC | Flu/Bu/TT | FK | Gilteritinib | No | Day 97 | Yes | Alive |
| AML086 | M | De novo | Intermediate | cCR2 | Haplo | MAC | Flu/Bu | FK/MMF/Cy | NA | No | Day 84 | No | Alive |
| AML087 | M | De novo | Favorable | cCR1 | MUD | MAC | Flu/Bu/TT | FK/MTX | NA | Yes | Pre-HSCT | No | Alive |
| AML109 | M | De novo | Adverse | cCR1 | URD | RIC | Flu/Mel | FK/MMF/Cy | NA | Yes | Never cleared | Yes | Dead |
| AML113 | F | De novo | Adverse | cCR1 | MUD | RIC | Flu/Mel | FK/MMF/Cy | NA | Yes | Day 496 | No | Alive |
| AML117 | M | De novo | Intermediate | cCR1 | MRD | MAC | Flu/Bu/TT | FK/MTX | Gilteritinib | Yes | Day 96 | No | Alive |
| AML118 | F | Therapy-related | Adverse | cCR1 | mmURD | MAC | Flu/Bu | FK/MMF/Cy | NA | No | Day 78 | No | Alive |
| AML122 | M | De novo | Intermediate | cCR1 | MRD | RIC | Flu/TT/TBI | FK | Gilteritinib | No | Day 90 | No | Alive |
| AML138 | M | Therapy-related | Intermediate | cCR1 | MRD | RIC | Flu/Cy/TBI | FK/MMF | NA | No | Day 83 | Yes | Alive |
| AML141 | M | De novo | Favorable | cCR2 | MRD | MAC | Flu/Bu | FK/MMF/Cy | NA | No | Pre-HSCT | No | Dead |
| AML142 | M | De novo | Adverse | cCR1 | mmURD | RIC | Flu/Mel | FK/MMF/Cy | NA | No | Day 287 | No | Alive |
| AML145 | F | De novo | Intermediate | cCR2 | Haplo | MAC | Flu/Bu | FK/MMF/Cy | NA | Yes | Pre-HSCT | No | Alive |
| AML147 | F | De novo | Intermediate | cCR1 | MUD | MAC | Flu/Bu/TT | FK | NA | No | Pre-HSCT | No | Alive |
| AML152 | M | De novo | Intermediate | cCR1 | MUD | RIC | Flu/TT/TBI | FK | NA | No | Day 175 | No | Alive |
| AML155 | F | De novo | Favorable | cCR2 | MUD | RIC | Flu/TBI | MMF/FK/Siro | NA | No | Never cleared | Yes | Dead |
| AML165 | F | De novo | Adverse | cCR1 | MUD | MAC | Bu/Cy | FK/MTX | NA | No | Never cleared | Yes | Dead |

**Table S7. Comparative prognostic performance of MRD strategies for relapse-free survival (RFS) and overall survival (OS).** Hazard ratios (HRs) were estimated using Cox proportional hazards regression models. Model discrimination was assessed using Harrell's concordance index (C-index). Model fit was evaluated using likelihood ratio (LR) statistics and Akaike Information Criterion (AIC), with lower AIC indicating improved model fit. ΔAIC values are shown relative to the ctDNA MRD (all mutations) model within each panel. All analyses were performed using the same evaluable patient cohorts for each endpoint.

**Panel A. Univariate Cox proportional hazards models for RFS** evaluating the association between different MRD strategies and RFS among patients who achieved first composite clinical remission (CR1) (n = 49; 32 relapse events). MRD approaches include standard-of-care (SOC) multiparameter flow cytometry (MFC), SOC combined MRD assessment (incorporating single-variant molecular MRD assessment when available in combination with MFC), ctDNA MRD using canonical mutations only, and personalized ctDNA MRD incorporating all patient-specific mutations.

| MRD strategy | N | Events | Hazard Ratio (95% CI) | p-value | C-index | LR $\chi^2$ | AIC | ΔAIC vs ctDNA |
|---|---|---|---|---|---|---|---|---|
| SOC MFC MRD | 49 | 32 | 3.61 (1.74–7.53) | $6.0 \times 10^{-4}$ | 0.619 | 10.55 | 210.66 | +24.44 |
| SOC Combined MRD | 49 | 32 | 4.43 (2.13–9.22) | $7.1 \times 10^{-5}$ | 0.650 | 14.97 | 206.24 | +20.54 |
| ctDNA MRD (canonical mutations only) | 49 | 32 | 5.71 (2.64–12.36) | $1.0 \times 10^{-5}$ | 0.717 | 21.49 | 199.72 | +14.03 |
| **ctDNA MRD (all mutations)** | **49** | **32** | **17.83 (5.15–61.79)** | **$5.5 \times 10^{-6}$** | **0.738** | **36.70** | **185.70** | **0 (reference)** |

**Panel B. Incremental prognostic value of personalized ctDNA for RFS.** Nested Cox regression analyses assessing the incremental prognostic value of adding personalized ctDNA MRD (all mutations) beyond SOC MRD approaches and ctDNA MRD based on canonical mutations alone for RFS. Improvements in model fit were quantified using likelihood ratio tests (LRTs) and changes in AIC (ΔAIC).

| Base model | Added variable | ΔLR $\chi^2$ | Δdf | LRT p-value | ΔAIC |
|---|---|---|---|---|---|
| SOC MFC MRD | ctDNA MRD (all mutations) | 26.44 | 1 | $2.7 \times 10^{-7}$ | **−24.44** |
| SOC Combined MRD | ctDNA MRD (all mutations) | 22.54 | 1 | $2.1 \times 10^{-6}$ | **−20.54** |
| ctDNA MRD (canonical only) | ctDNA MRD (all mutations) | 17.41 | 1 | $3.0 \times 10^{-5}$ | **−14.03** |

**Panel C. Univariate Cox proportional hazards models for OS** evaluating the association between MRD strategies and overall survival (OS) in the full cohort (n = 56; 31 death events), using the same MRD definitions as in Panel A.

| MRD strategy | N | Events | Hazard Ratio (95% CI) | p-value | C-index | LR $\chi^2$ | AIC | ΔAIC vs ctDNA |
|---|---|---|---|---|---|---|---|---|
| SOC MFC MRD | 56 | 31 | 5.36 (2.60–11.03) | $5.2 \times 10^{-6}$ | 0.681 | 19.26 | 203.98 | +25.22 |
| SOC Combined MRD | 56 | 31 | 6.21 (2.97–12.98) | $1.2 \times 10^{-6}$ | 0.702 | 23.09 | 200.16 | +21.40 |
| ctDNA MRD (canonical mutations only) | 56 | 31 | 11.08 (4.76–25.78) | $2.4 \times 10^{-8}$ | 0.765 | 37.07 | 186.17 | +7.41 |
| **ctDNA MRD (all mutations)** | **56** | **31** | **19.14 (6.51–56.22)** | **$8.0 \times 10^{-8}$** | **0.793** | **46.45** | **178.76** | **0 (reference)** |

**Panel D. Incremental prognostic value of personalized ctDNA-MRD for OS.** Nested Cox regression analyses evaluating the incremental prognostic value of personalized ctDNA MRD (all mutations) beyond SOC MRD approaches and ctDNA MRD based on canonical mutations alone for OS, assessed using LRTs and ΔAIC.

| Base model | Added variable | ΔLR $\chi^2$ | Δdf | LRT p-value | ΔAIC |
|---|---|---|---|---|---|
| SOC MFC MRD | ctDNA MRD (all mutations) | 27.19 | 1 | $1.8 \times 10^{-7}$ | **−25.19** |

| | | | | | |
|---|---|---|---|---|---|
| SOC Combined MRD | ctDNA MRD (all mutations) | 23.40 | 1 | $1.3 \times 10^{-6}$ | **−21.40** |
| ctDNA MRD (canonical only) | ctDNA MRD (all mutations) | 10.34 | 1 | 0.0013 | **−8.34** |

Across both RFS and OS analyses, personalized ctDNA MRD incorporating all patient-specific mutations demonstrated the highest discriminatory performance (C-index), strongest effect sizes, and the greatest improvement in model fit compared with SOC MRD strategies and ctDNA MRD limited to canonical mutations, supporting its added prognostic value in this cohort.

### Supplementary Figures

**Figure S1.** Flow cytometric purification of AML blasts and T cells for WES genotyping. (A) Representative gating strategy used for flow sorting. Post-sort purity assessments for (B) blasts and (C) T cells.

A Representative flow cytometry sorting strategy for blasts and T cells

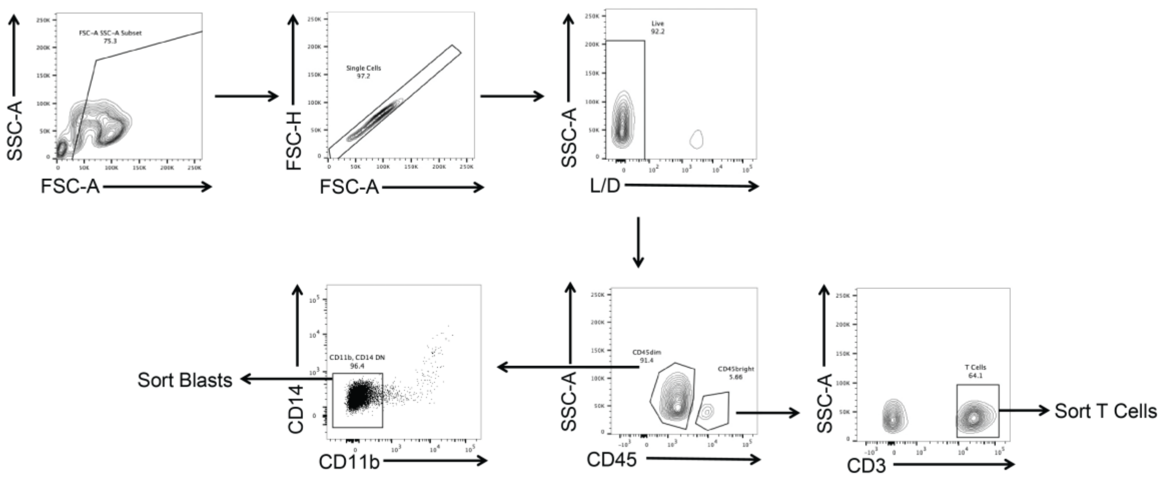


B Representative post-sort purity of blasts

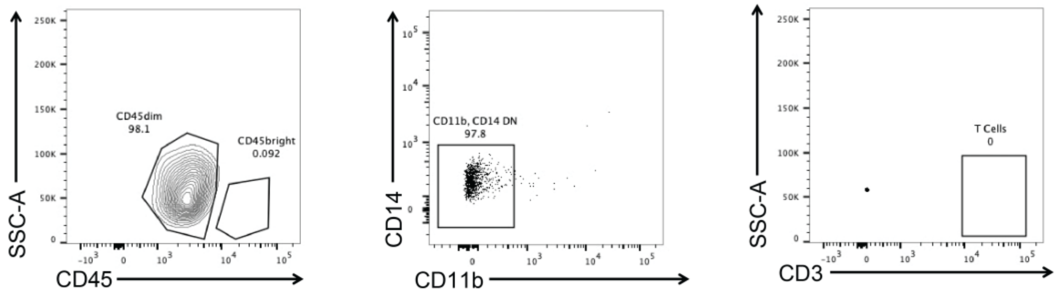


C Representative post-sort purity of T cells

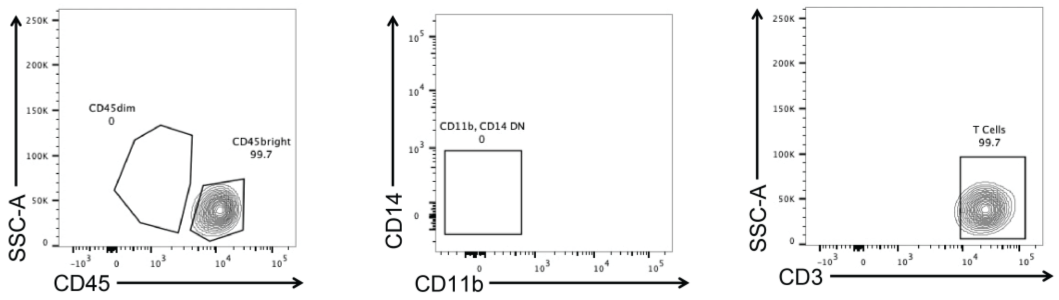

**Figure S2. Characterization of canonical vs noncanonical mutations.** (A–E) Comparative analysis of evolutionary conservation (PHAST, A; Zoonomia; B), functional impact (SIFT; C, ESM; D), and composite CADD scores (E) between canonical and noncanonical variants.

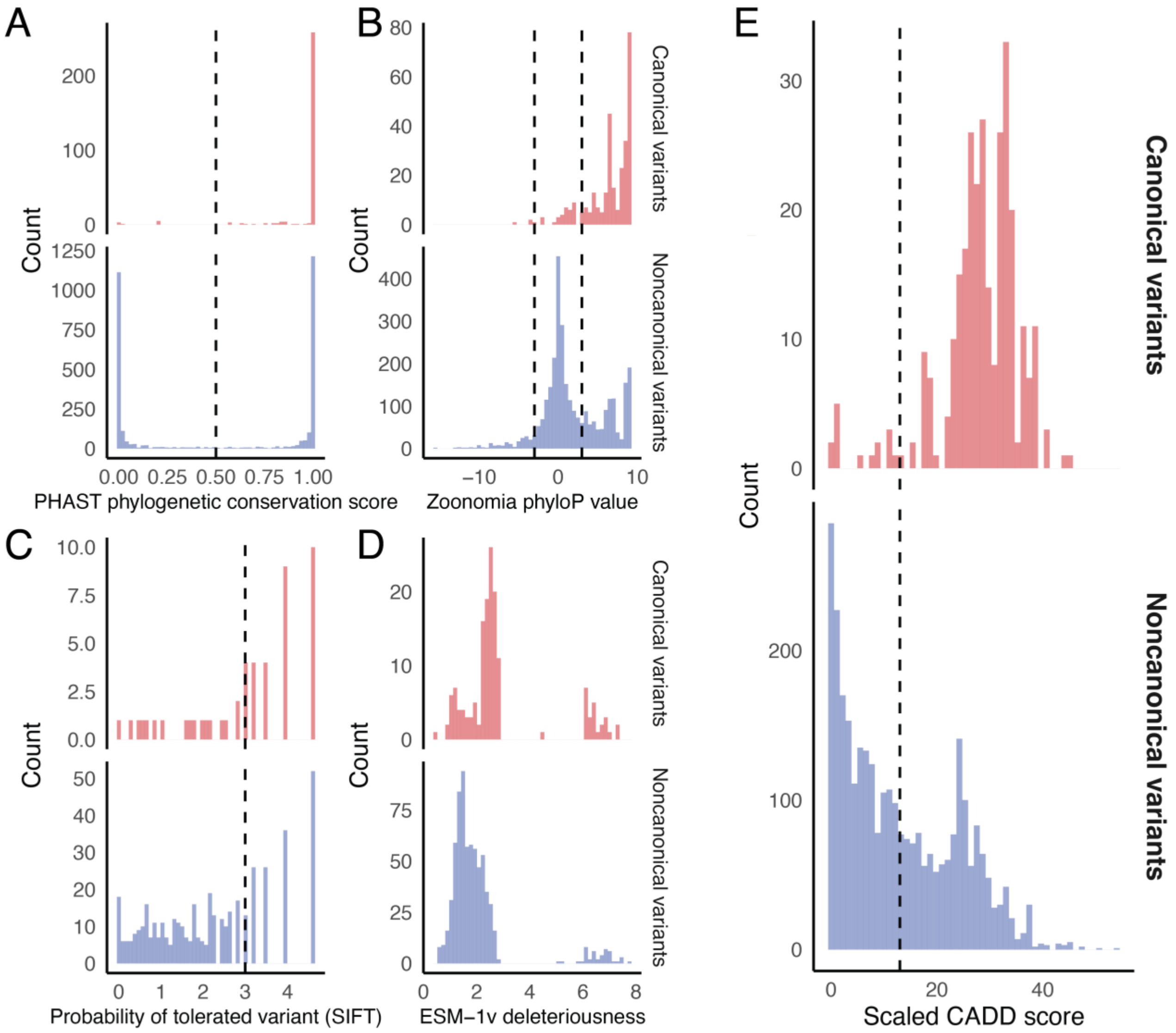

**Figure S3. Baseline VAF distributions of canonical and noncanonical somatic SNVs identified by WES.** Violin plots depicting the distribution of variant allele fractions (VAFs) for canonical AML-associated SNVs and noncanonical (patient-specific) SNVs identified by WES across all patients. Each point represents the VAF of an individual variant, pooled across patients. Median VAFs are indicated by horizontal lines. P-value based on Wilcoxon rank-sum test.

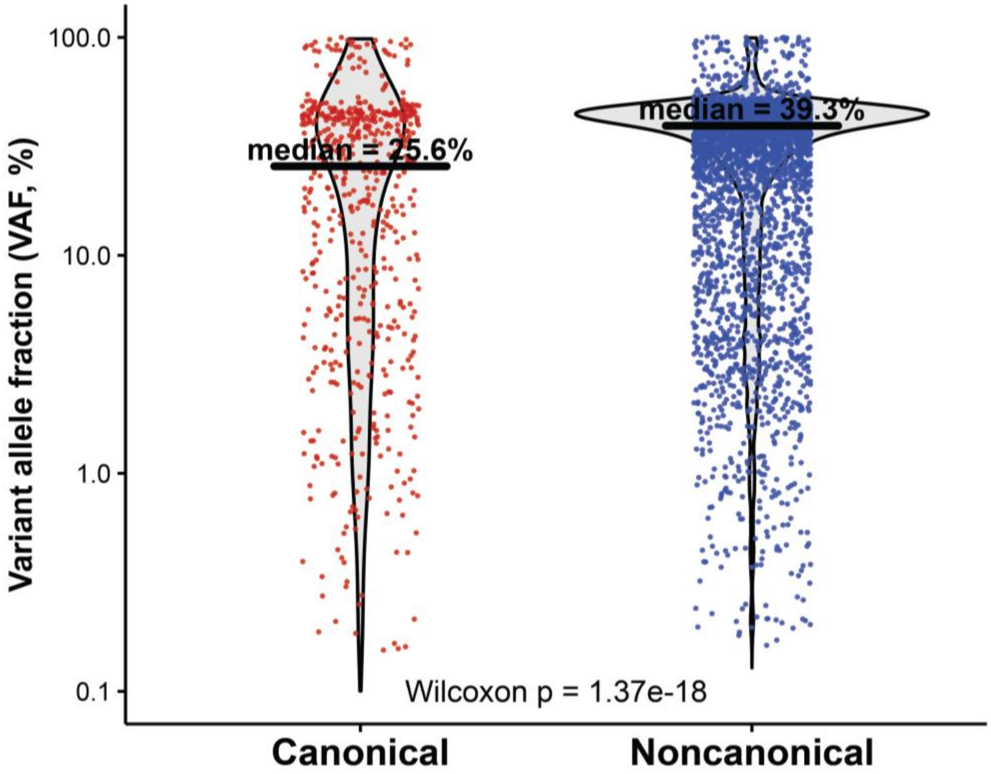

**Figure S4. Concordance of clinically reported mutations across sample compartments.** (A) Qualitative detection rates for canonical SNVs with VAF >1% at diagnosis across paired PB-ctDNA, BMMC, and PBMC compartments. (B–C) Qualitative concordance between each pair of compartments (B: BMMC vs PBMC; C: BMMC vs PB-ctDNA; D: PBMC vs PB-ctDNA)

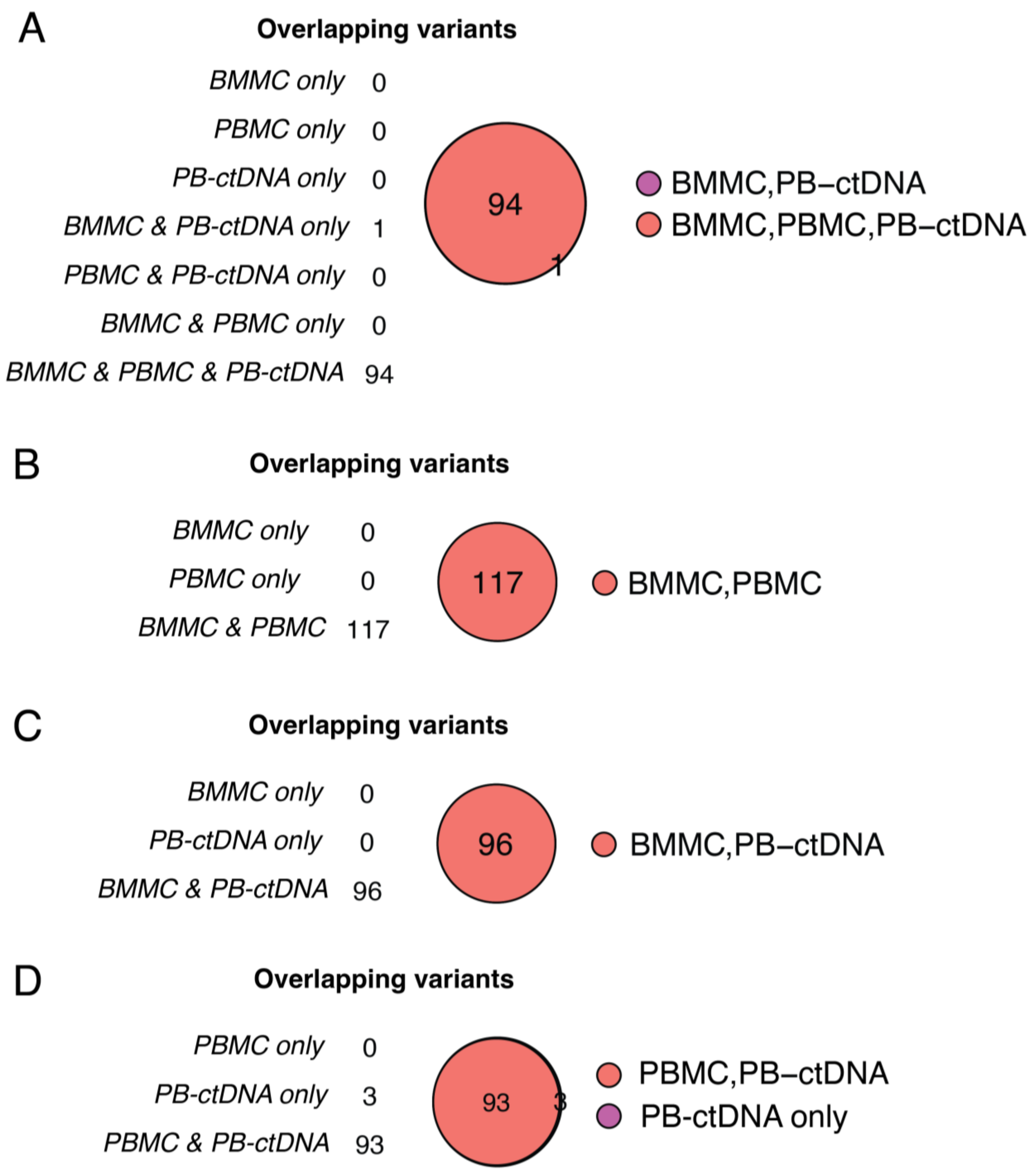

**Figure S5. Sensitivity of ctDNA in detecting low-frequency mutations.**
(A–B) Paired SNV VAF comparisons between PB-ctDNA and BMMCs (A) or PBMCs (B), limited to variants with cellular VAF <1%. (C) Paired SNV VAFs in BMMCs for variants with corresponding PBMC VAF <1%.

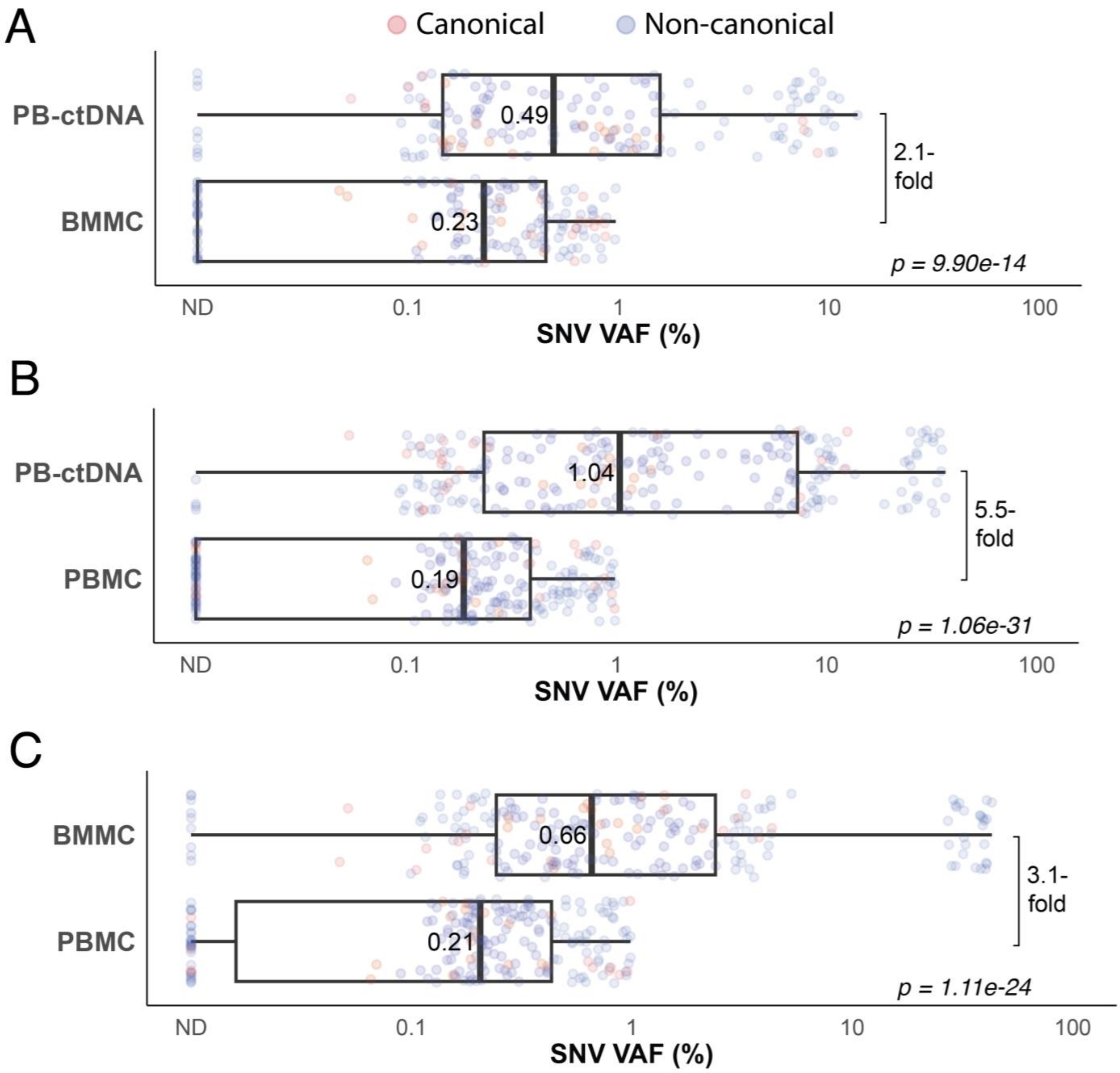

**Figure S6. Sensitivity of ctDNA in detecting low-burden disease.**
(A–B) Paired mean tumor allele fraction (AF) comparisons between PB-ctDNA and BMMCs (A) or PBMCs (B), restricted to samples with cellular tumor AF <1%. (C) Paired mean tumor AFs in BMMCs for samples with corresponding PBMC tumor AF <1%.

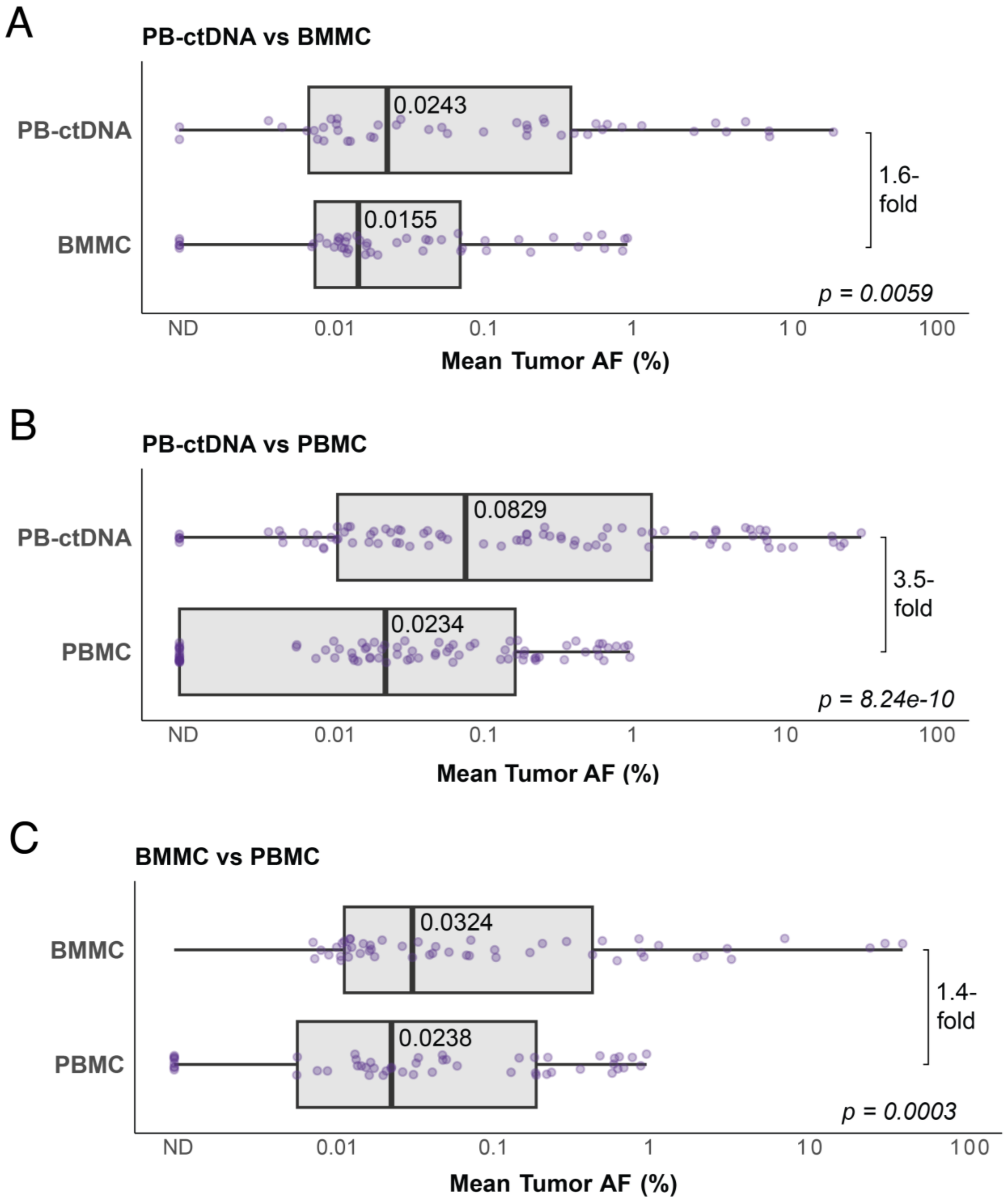

**Figure S7. Comparison of PB-ctDNA and BM-ctDNA compartments.** (A) Variant-level VAF correlation between paired PB and BM ctDNA samples. (B) Sample-level mean tumor AF correlation between PB and BM ctDNA, across active and remission states.

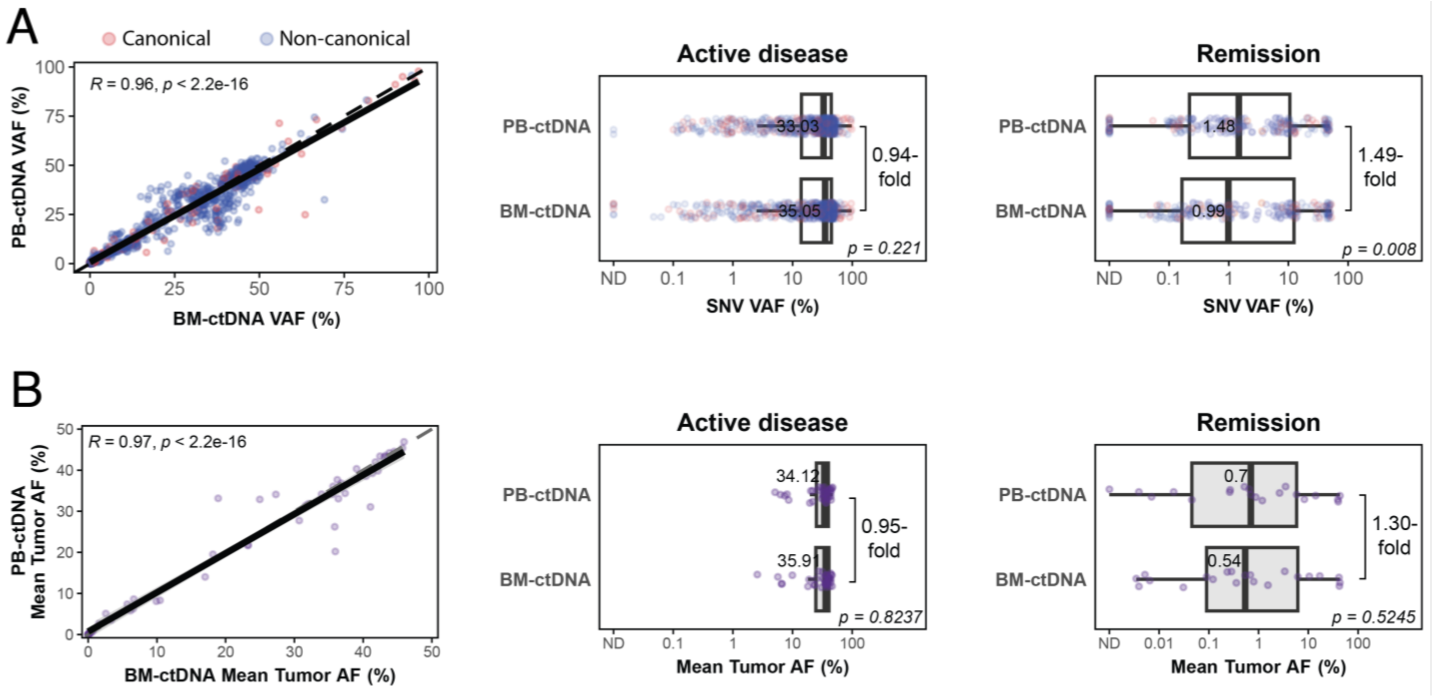

**Figure S8. Comparison of ctDNA-MRD and standard-of-care (SOC) MRD.** Alluvial plot of 204 paired AML samples taken during timepoints of composite complete remission across the cohort (n = 50 patients), illustrating the concordance of SOC-MRD (MFC ± single-variant molecular assays) and ctDNA-MRD results (derived from AML-CAPP-Seq).

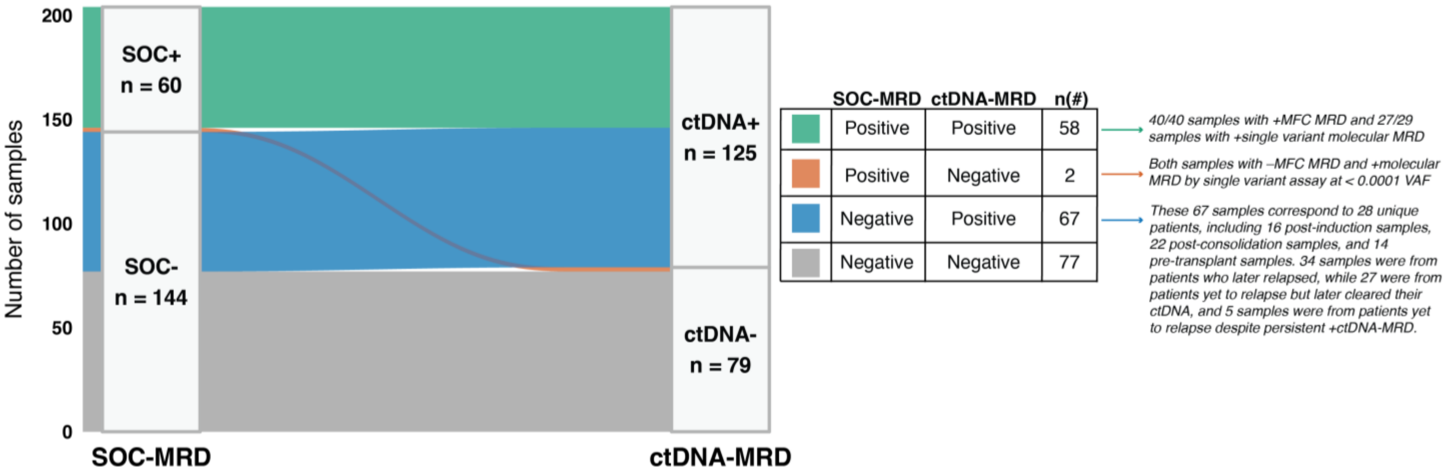

**Figure S9. Comparison of ctDNA-MRD and SOC-MRD for clinical outcome prediction in newly diagnosed AML.** (A–D) Kaplan–Meier curves depicting relapse-free survival (RFS) among patients with newly diagnosed AML at the time of study enrollment who achieved an initial composite complete remission (n = 47), stratified by MRD status using SOC multiparameter flow cytometry (MFC) alone (A), SOC combined MRD assessment (MFC + molecular assays, B), ctDNA-MRD using canonical AML gene mutations only (C), or ctDNA-MRD using all whole-exome sequencing (WES)–derived personalized tumor variants (D). (E–H) Kaplan–Meier curves depicting overall survival (OS) among patients with newly diagnosed AML at study enrollment (n = 50), stratified by achievement of MRD negativity at any timepoint using SOC MFC MRD alone (E), SOC combined MRD (F), ctDNA-MRD using canonical variants only (G), or ctDNA-MRD using all personalized tumor variants (H). Hazard ratios (HR), 95% confidence intervals (CI), and p-values from log-rank tests are shown for each comparison.

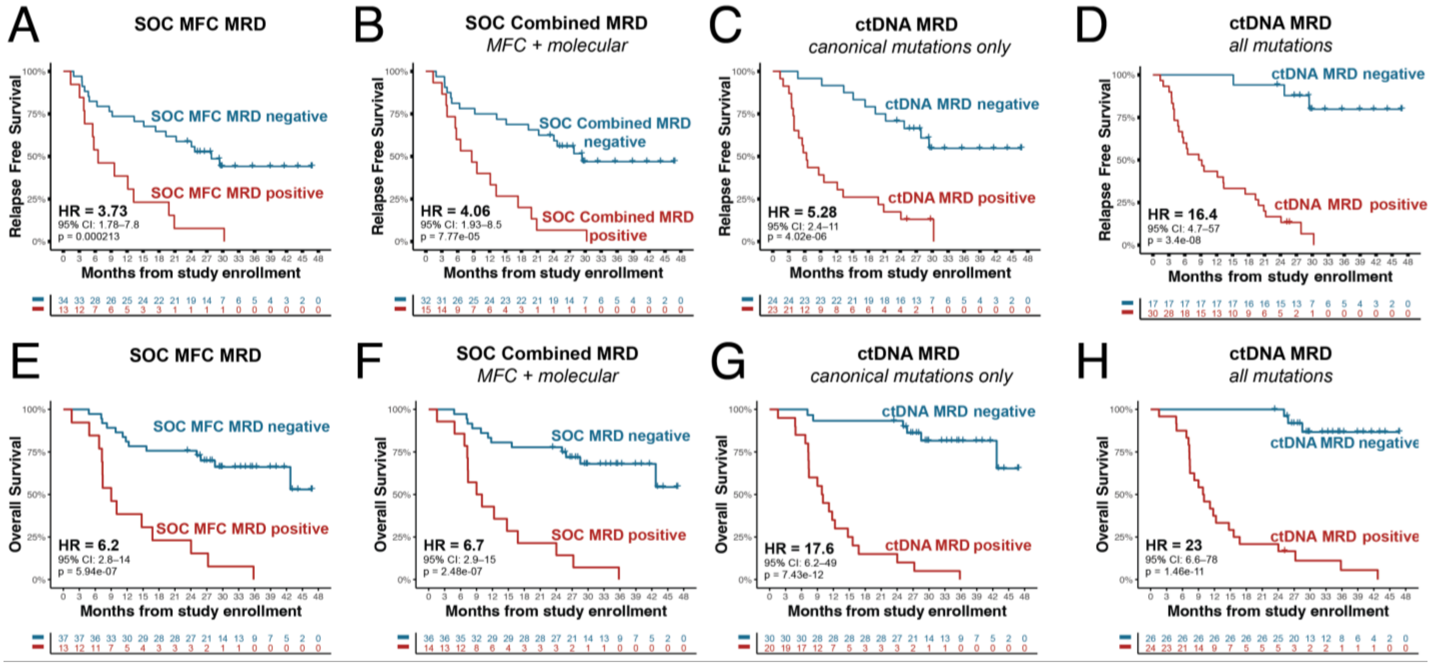


**Panel I. Univariate Cox proportional hazards models for RFS in newly diagnosed AML.** Univariate Cox models evaluating the association between different MRD strategies and relapse-free survival among patients with newly diagnosed AML who achieved CR1 (n = 47; 31 relapse events). Hazard ratios (HRs) were estimated using Cox proportional hazards regression models. Model discrimination was assessed using Harrell's concordance index (C-index). Model fit was evaluated using likelihood ratio (LR) statistics and Akaike Information Criterion (AIC), with lower AIC indicating improved model fit. ΔAIC values are shown relative to the personalized ctDNA-MRD (all mutations) model within each panel.

| MRD strategy | N | Events | Hazard Ratio (95% CI) | p-value | C-index | LR $\chi^2$ | AIC | ΔAIC vs ctDNA |
|---|---|---|---|---|---|---|---|---|
| SOC MFC MRD | 47 | 31 | 3.73 (1.78–7.83) | $5.1 \times 10^{-4}$ | 0.627 | 10.88 | 200.26 | +22.04 |
| SOC Combined MRD | 47 | 31 | 4.06 (1.93–8.54) | $2.2 \times 10^{-4}$ | 0.639 | 12.84 | 198.29 | +20.07 |
| ctDNA MRD (canonical mutations only) | 47 | 31 | 5.28 (2.43–11.48) | $2.7 \times 10^{-5}$ | 0.711 | 19.23 | 191.90 | +13.68 |
| **ctDNA MRD (all mutations)** | **47** | **31** | **16.42 (4.74–56.88)** | $\mathbf{1.0 \times 10^{-5}}$ | **0.734** | **33.99** | **178.22** | **0 (reference)** |

**Panel J. Incremental prognostic value of personalized ctDNA-MRD for RFS in newly diagnosed AML.** Nested Cox regression analyses assessing the incremental prognostic value of adding personalized ctDNA-MRD (all mutations) beyond SOC MRD approaches and ctDNA-MRD based on canonical mutations alone.

| Base model | Added variable | ΔLR $\chi^2$ | Δdf | LRT p-value | ΔAIC |
|---|---|---|---|---|---|
| SOC MFC MRD | ctDNA MRD (all mutations) | 23.58 | 1 | $1.2 \times 10^{-6}$ | −21.58 |
| SOC Combined MRD | ctDNA MRD (all mutations) | 21.69 | 1 | $3.2 \times 10^{-6}$ | −19.69 |

| Base model | Added variable | $\Delta$LR $\chi^2$ | $\Delta$df | LRT p-value | $\Delta$AIC |
|---|---|---|---|---|---|
| ctDNA MRD (canonical only) | ctDNA MRD (all mutations) | 16.68 | 1 | $4.4 \times 10^{-5}$ | −14.68 |

**Panel K. Univariate Cox proportional hazards models for OS in newly diagnosed AML.** Univariate Cox models evaluating the association between different MRD strategies and overall survival among patients with newly diagnosed AML (n = 50; 26 death events).

| MRD strategy | N | Events | Hazard Ratio (95% CI) | p-value | C-index | LR $\chi^2$ | AIC | $\Delta$AIC vs ctDNA |
|---|---|---|---|---|---|---|---|---|
| SOC MFC MRD | 50 | 26 | 6.18 (2.76–13.87) | $9.8 \times 10^{-6}$ | 0.680 | 17.99 | 164.92 | +22.73 |
| SOC Combined MRD | 50 | 26 | 6.65 (2.94–15.08) | $5.6 \times 10^{-6}$ | 0.691 | 19.72 | 163.19 | +21.02 |
| ctDNA MRD (canonical mutations only) | 50 | 26 | 17.56 (6.25–49.38) | $5.5 \times 10^{-8}$ | 0.786 | 39.19 | 143.73 | +1.55 |
| **ctDNA MRD (all mutations)** | **50** | **26** | **22.75 (6.64–77.89)** | $\mathbf{6.5 \times 10^{-7}}$ | **0.802** | **42.48** | **142.17** | **0 (reference)** |

**Panel L. Incremental prognostic value of personalized ctDNA-MRD for OS in newly diagnosed AML.** Nested Cox regression analyses evaluating the incremental prognostic value of personalized ctDNA-MRD (all mutations) beyond SOC MRD approaches and ctDNA-MRD based on canonical mutations alone.

| Base model | Added variable | $\Delta$LR $\chi^2$ | $\Delta$df | LRT p-value | $\Delta$AIC |
|---|---|---|---|---|---|
| SOC MFC MRD | ctDNA MRD (all mutations) | 24.73 | 1 | $6.6 \times 10^{-7}$ | −22.73 |
| SOC Combined MRD | ctDNA MRD (all mutations) | 23.02 | 1 | $1.6 \times 10^{-6}$ | −21.02 |
| ctDNA MRD (canonical only) | ctDNA MRD (all mutations) | 6.51 | 1 | 0.0108 | −4.51 |

**Figure S10. Prognostic performance of ctDNA-MRD stratified by availability of approved molecular MRD markers. (A)** Kaplan–Meier curves depicting RFS in patients **without approved molecular MRD markers** who achieved cCR1 (n = 32), comparing standard-of-care (SOC) multiparameter flow cytometry (MFC) and personalized ctDNA-MRD. **(B)** Univariate Cox proportional hazards models for RFS in this subgroup, summarizing hazard ratios (HRs), 95% confidence intervals (CIs), model discrimination (C-index), likelihood ratio statistics, and Akaike Information Criterion (AIC) values for each MRD strategy. **(C)** Nested Cox regression analyses assessing the incremental prognostic value of adding personalized ctDNA-MRD beyond SOC MFC. **(D)** Kaplan–Meier curves depicting RFS in patients **with approved molecular MRD markers** who achieved cCR1 (n = 17), comparing SOC combined MRD assessment (MFC ± single-variant molecular testing) and personalized ctDNA-MRD. **(E)** Univariate Cox proportional hazards models for RFS in patients with approved molecular MRD markers, summarizing HRs, 95% CIs, and model performance metrics for each MRD strategy. **(F)** Nested Cox regression analyses evaluating the incremental prognostic value of personalized ctDNA-MRD beyond SOC combined MRD in this subgroup.

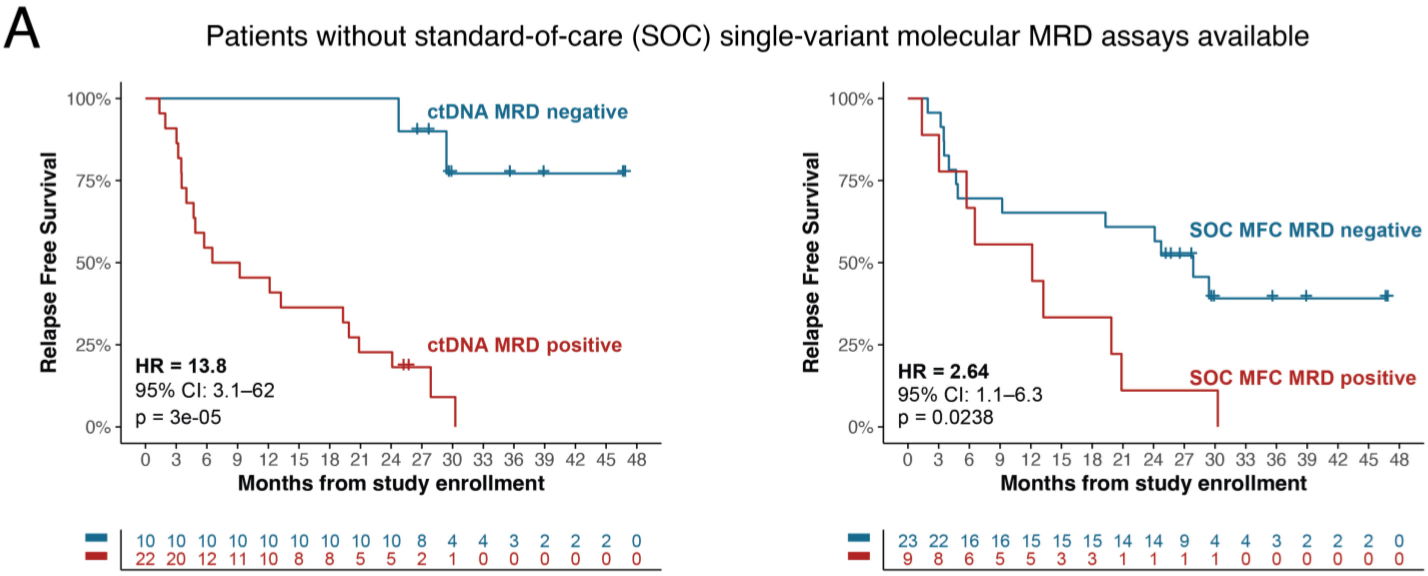


**Panel B. Univariate Cox proportional hazards models for RFS in patients without SOC molecular MRD**

| MRD strategy | N | Events | Hazard Ratio (95% CI) | p-value | C-index | LR χ² | AIC | ΔAIC vs ctDNA |
|---|---|---|---|---|---|---|---|---|
| SOC MFC MRD | 32 | 22 | 2.64 (1.10–6.33) | $2.9 \times 10^{-2}$ | 0.588 | 4.39 | 127.39 | +13.79 |
| ctDNA MRD | 32 | 22 | 13.78 (3.06–62.12) | $6.4 \times 10^{-4}$ | 0.713 | 20.18 | 113.60 | 0 (reference) |

**Panel C. Incremental prognostic value of personalized ctDNA MRD for RFS in patients without SOC molecular MRD**

| Base model | Added variable | ΔLR χ² | Δdf | LRT p-value | ΔAIC |
|---|---|---|---|---|---|
| SOC MFC MRD | ctDNA MRD | 15.79 | 1 | $7.1 \times 10^{-5}$ | −13.79 |

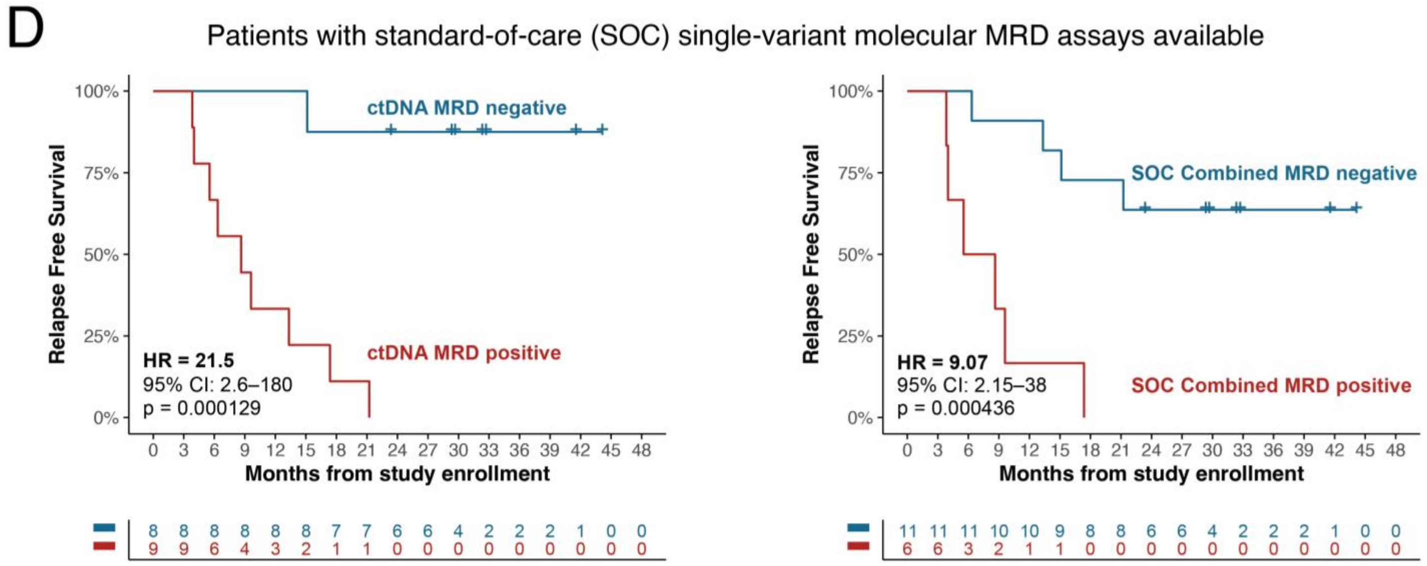


### Panel E. Univariate Cox proportional hazards models for RFS in patients with SOC molecular MRD

| MRD strategy | N | Events | Hazard Ratio (95% CI) | p-value | C-index | LR χ² | AIC | ΔAIC vs ctDNA (all) |
|---|---|---|---|---|---|---|---|---|
| SOC Combined MRD | 17 | 10 | 9.07 (2.15–38.23) | $2.7 \times 10^{-3}$ | 0.743 | 9.57 | 42.39 | +10.39 |
| **ctDNA MRD** | **17** | **10** | **21.51 (2.60–177.6)** | **$4.4 \times 10^{-3}$** | **0.796** | **14.68** | **32.00** | **0 (reference)** |

### Panel F. Incremental prognostic value of personalized ctDNA MRD for RFS in patients with SOC molecular MRD

| Base model | Added variable | ΔLR χ² | Δdf | LRT p-value | ΔAIC |
|---|---|---|---|---|---|
| SOC Combined MRD | ctDNA MRD | 6.39 | 1 | 0.011 | −4.39 |

**Figure S11. Swimmer plots comparing ctDNA-MRD with standard-of-care molecular MRD assessments.** Swimmer plots depicting longitudinal disease course and MRD status for the subset of AML patients with approved standard-of-care (SOC) single-variant molecular MRD testing available (n = 18). Bars represent time on study for each patient and are color-coded to indicate SOC MRD status, with distinct shading reflecting results from multiparameter flow cytometry (MFC) versus single-variant molecular MRD assays when performed. Superimposed points denote serial plasma ctDNA levels measured by AML-CAPP-Seq over time.

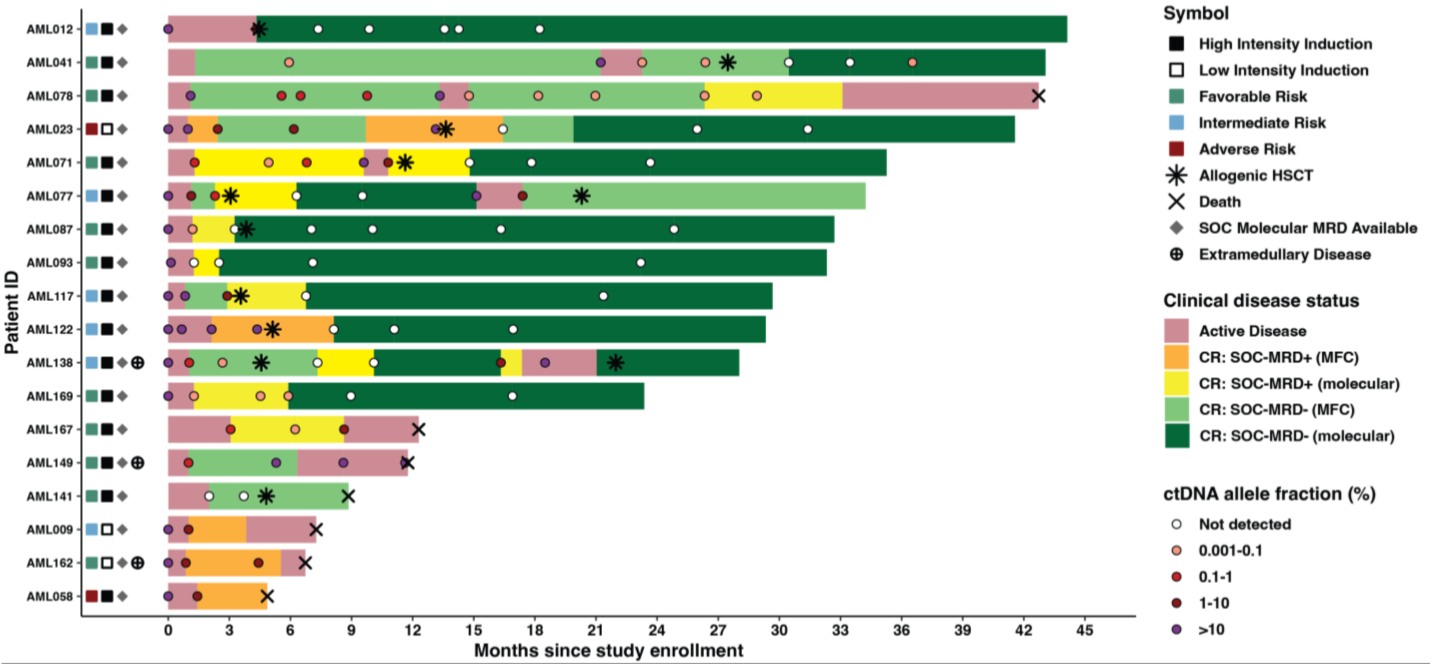

**Figure S12. Prognostic performance of ctDNA-MRD in patients with adverse-risk AML.** Kaplan–Meier analyses and multivariable modeling of RFS and OS in patients with adverse-risk AML stratified by MRD status. (A–C) Kaplan–Meier curves depicting RFS among adverse-risk patients who achieved an initial composite complete remission (n = 26), stratified by MRD status assessed using personalized circulating tumor DNA–based MRD incorporating all tumor-specific variants (A), standard-of-care (SOC) combined MRD assessment (multiparameter flow cytometry ± approved molecular assays, when available) (B), and multiparameter flow cytometry (MFC) alone (C). (D–F) Kaplan–Meier curves depicting OS among all adverse-risk patients (n = 30), stratified by MRD status assessed using personalized ctDNA-MRD (D), SOC combined MRD assessment (E), and MFC alone (F). (G) Univariate Cox proportional hazards models for RFS comparing SOC MFC MRD, SOC combined MRD, ctDNA-MRD based on canonical AML driver mutations only, and personalized ctDNA-MRD incorporating all tumor-specific variants. Hazard ratios (HRs), 95% confidence intervals (CIs), concordance indices (C-indices), likelihood ratio (LR) statistics, and Akaike Information Criterion (AIC) values are shown. (H) Nested Cox regression analyses evaluating the incremental prognostic value of personalized ctDNA-MRD beyond SOC MRD approaches and canonical ctDNA-MRD for RFS. Improvements in model fit are quantified using likelihood ratio tests (LRTs) and changes in AIC (ΔAIC). (I) Univariate Cox proportional hazards models for OS comparing SOC MFC MRD, SOC combined MRD, canonical ctDNA-MRD, and personalized ctDNA-MRD. (J) Nested Cox regression analyses evaluating the incremental prognostic value of personalized ctDNA-MRD beyond SOC MRD approaches and canonical ctDNA-MRD for OS.

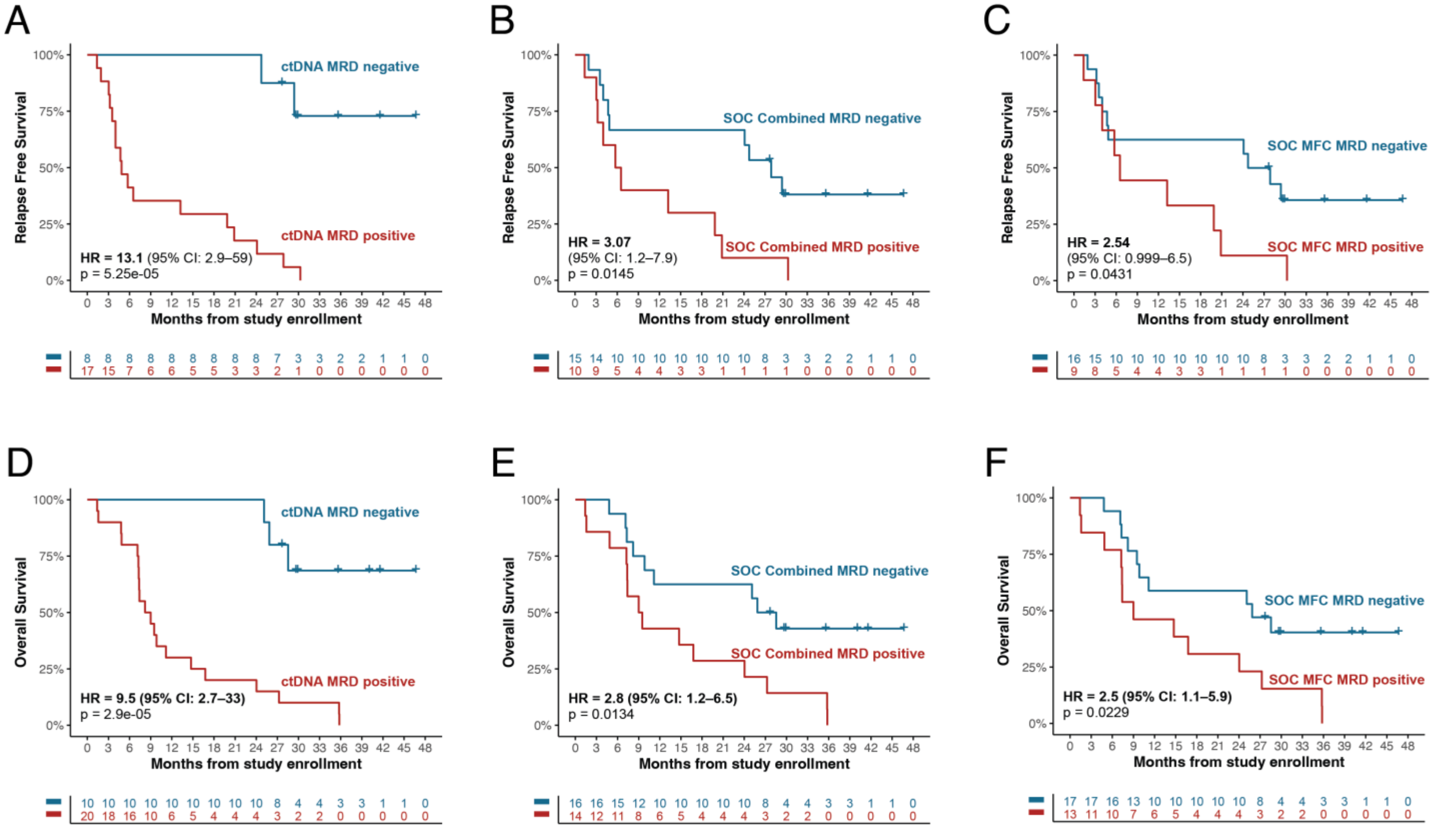

### Panel G. Univariate Cox proportional hazards models for RFS in adverse-risk AML

| MRD strategy | N | Events | Hazard Ratio (95% CI) | p-value | C-index | LR χ² | AIC | ΔAIC vs ctDNA (all) |
|---|---|---|---|---|---|---|---|---|
| SOC MFC MRD | 25 | 19 | 2.54 (1.00–6.48) | $5.0 \times 10^{-2}$ | 0.593 | 3.69 | 99.53 | +13.08 |
| SOC Combined MRD | 25 | 19 | 3.07 (1.20–7.87) | $1.9 \times 10^{-2}$ | 0.625 | 5.38 | 97.85 | +11.40 |
| ctDNA MRD (canonical mutations only) | 25 | 19 | 7.90 (2.45–25.51) | $5.5 \times 10^{-4}$ | 0.724 | 15.06 | 88.16 | +1.71 |
| **ctDNA MRD (all mutations)** | **25** | **19** | **13.05 (2.87–59.30)** | **$8.8 \times 10^{-4}$** | **0.726** | **18.17** | **86.45** | **0 (reference)** |

### Panel H. Incremental prognostic value of personalized ctDNA-MRD for RFS in adverse-risk AML (nested Cox models)

| Base model | Added variable | ΔLR χ² | Δdf | LRT p-value | ΔAIC |
|---|---|---|---|---|---|
| SOC MFC MRD | ctDNA MRD (all mutations) | 14.92 | 1 | $1.1 \times 10^{-4}$ | −12.92 |
| SOC Combined MRD | ctDNA MRD (all mutations) | 12.88 | 1 | $3.3 \times 10^{-4}$ | −10.88 |
| ctDNA MRD (canonical only) | ctDNA MRD (all mutations) | 3.71 | 1 | $5.4 \times 10^{-2}$ | −1.71 |

### Panel I. Univariate Cox proportional hazards models for OS in adverse-risk AML

| MRD strategy | N | Events | Hazard Ratio (95% CI) | p-value | C-index | LR χ² | AIC | ΔAIC vs ctDNA (all) |
|---|---|---|---|---|---|---|---|---|
| SOC MFC MRD | 30 | 23 | 2.55 (1.11–5.85) | $2.8 \times 10^{-2}$ | 0.607 | 4.86 | 126.63 | +14.58 |
| SOC Combined MRD | 30 | 23 | 2.80 (1.20–6.54) | $1.8 \times 10^{-2}$ | 0.616 | 5.82 | 125.67 | +13.62 |
| ctDNA MRD (canonical mutations only) | 30 | 23 | 1.03 (1.00–1.06) | $2.8 \times 10^{-2}$ | 0.657 | 4.04 | 127.46 | +15.41 |
| **ctDNA MRD (all mutations)** | **30** | **23** | **9.50 (2.74–32.91)** | **$3.9 \times 10^{-4}$** | **0.717** | **18.94** | **112.05** | **0 (reference)** |

### Panel J. Incremental prognostic value of personalized ctDNA-MRD for OS in adverse-risk AML (nested Cox models)

| Base model | Added variable | ΔLR χ² | Δdf | LRT p-value | ΔAIC |
|---|---|---|---|---|---|
| SOC MFC MRD | ctDNA MRD (all mutations) | 16.58 | 1 | $4.7 \times 10^{-5}$ | −14.58 |
| SOC Combined MRD | ctDNA MRD (all mutations) | 15.50 | 1 | $8.3 \times 10^{-5}$ | −13.50 |
| ctDNA MRD (canonical only) | ctDNA MRD (all mutations) | 15.42 | 1 | $8.6 \times 10^{-5}$ | −13.41 |

**Figure S13. ctDNA-MRD outperforms cellular compartments for predicting clinical outcomes.** (A) Kaplan–Meier curves depicting relapse-free survival (RFS) for patients who achieved an initial composite complete remission (CR), stratified by MRD status per AML-CAPP-Seq using plasma ctDNA (left) or PBMCs (right), among those with paired samples available (n = 48). (B) RFS for the subset of patients with paired plasma ctDNA and BMMC samples available (n = 28), stratified by MRD status in each compartment. (C) Kaplan–Meier curves showing overall survival (OS) for all patients in the cohort (n = 56), stratified by MRD negativity at any timepoint based on plasma ctDNA, PBMCs, or BMMCs. Hazard ratios (HR), 95% confidence intervals (CI), and p-values were calculated using the log-rank test.

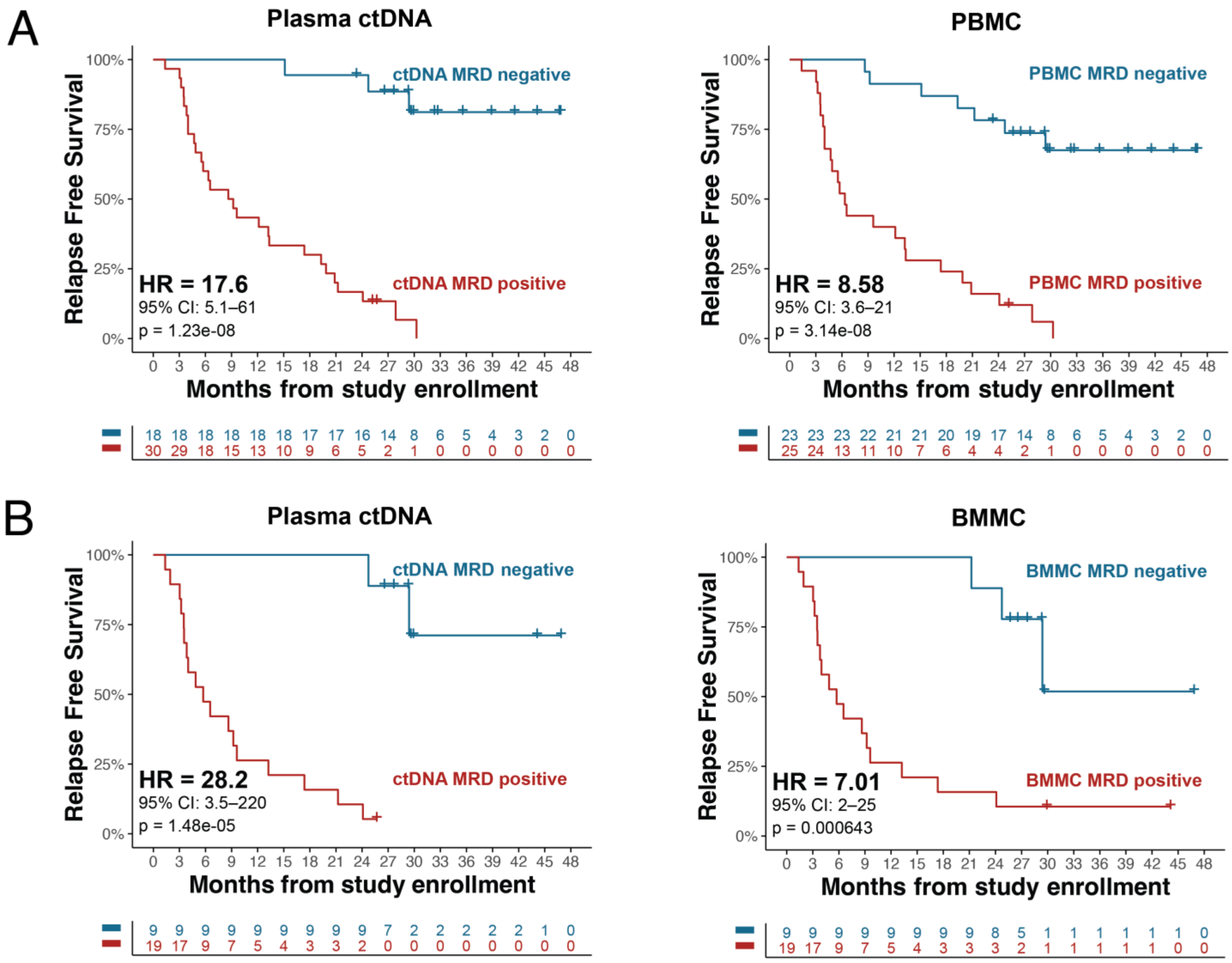

C

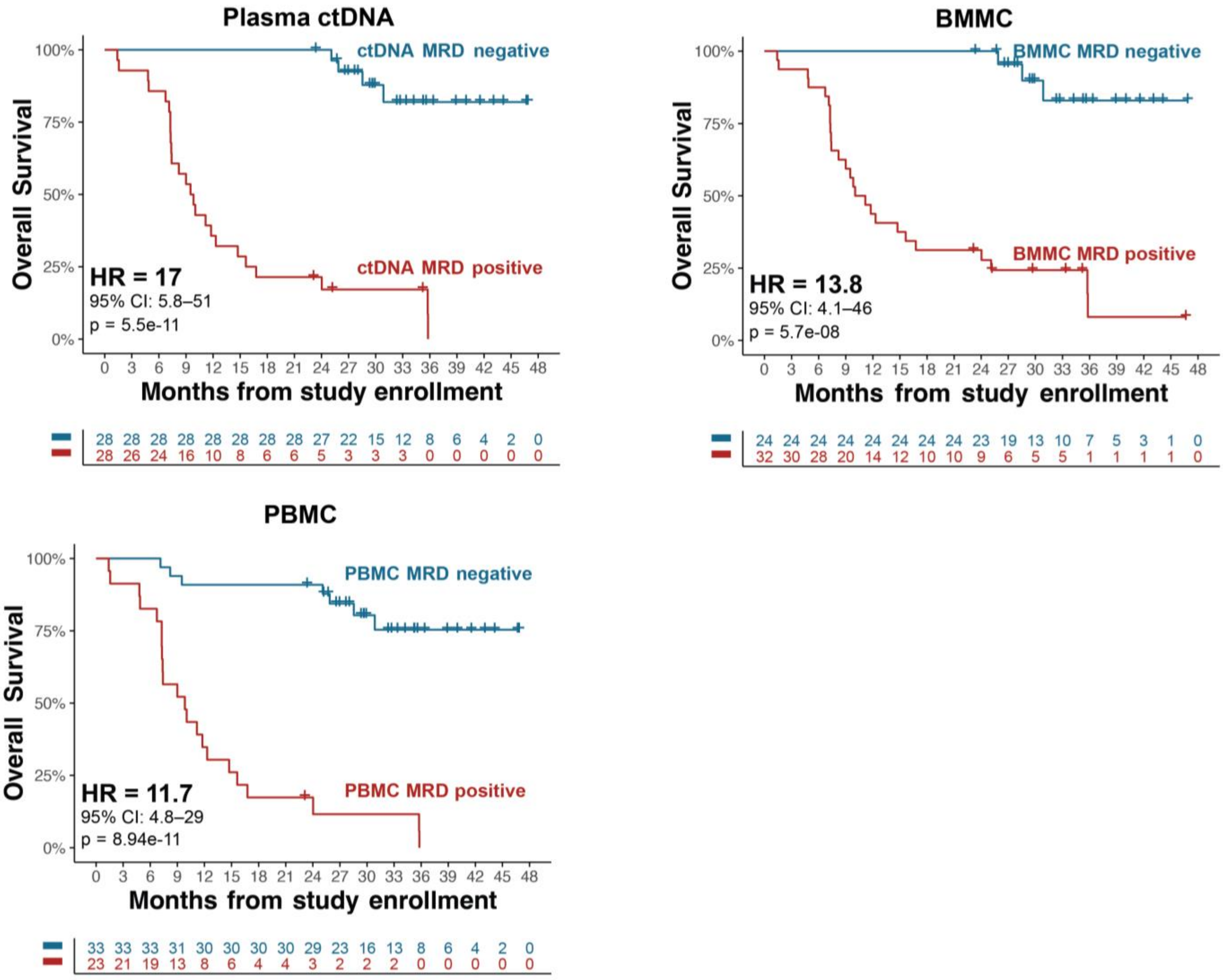

Plasma ctDNA
ctDNA MRD negative
ctDNA MRD positive
HR = 17
95% CI: 5.8–51
p = 5.5e-11
Overall Survival
Months from study enrollment
100%
75%
50%
25%
0%
0 3 6 9 12 15 18 21 24 27 30 33 36 39 42 45 48
28 28 28 28 28 28 28 28 27 22 15 12 8 6 4 2 0
28 26 24 16 10 8 6 6 5 3 3 3 0 0 0 0 0
BMMC
BMMC MRD negative
BMMC MRD positive
HR = 13.8
95% CI: 4.1–46
p = 5.7e-08
24 24 24 24 24 24 24 24 23 19 13 10 7 5 3 1 0
32 30 28 20 14 12 10 10 9 6 5 5 1 1 1 1 0
PBMC
PBMC MRD negative
PBMC MRD positive
HR = 11.7
95% CI: 4.8–29
p = 8.94e-11
33 33 33 31 30 30 30 30 29 23 16 13 8 6 4 2 0
23 21 19 13 8 6 4 4 3 2 2 2 0 0 0 0 0

**Figure S14. Association of ctDNA clearance kinetics and durability with RFS.** (A) Kaplan–Meier analysis of RFS stratified by timing of initial ctDNA MRD clearance, comparing early (<180 days) versus late (≥180 days) clearance among patients who achieved ctDNA negativity. (B) Kaplan–Meier analysis of RFS stratified by duration of ctDNA negativity. Hazard ratios were estimated using Cox proportional hazards regression, with p-values derived from log-rank tests.

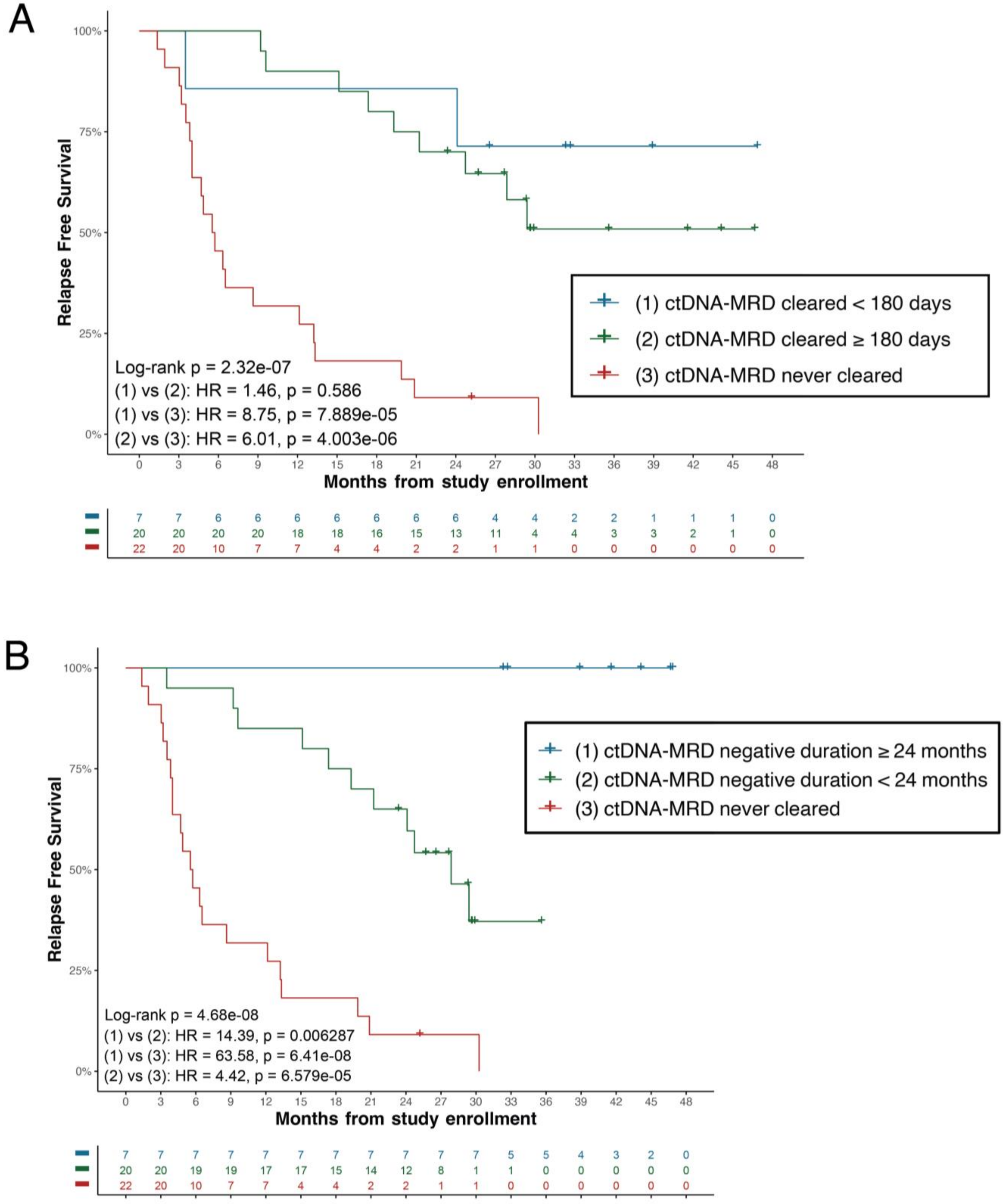

**Figure S15. Comparison of ctDNA-MRD and conventional marrow-based NGS for post-transplant relapse risk stratification.** Kaplan–Meier analyses of RFS among patients who underwent allogeneic HSCT (n = 26) with at least one evaluable molecular marker by clinical NGS and/or single-variant assays, comparing conventional bone marrow-based MRD assessment using the latter versus ctDNA–based MRD assessment. (A) RFS survival stratified by pre-transplant MRD status assessed by conventional marrow-based NGS (left) vs ctDNA-MRD (right). (B) RFS stratified by combined pre- and post-transplant MRD status assessed by conventional marrow-based NGS (left) vs ctDNA-MRD (right). Small subgroup sizes limited precise hazard ratio estimation. Hazard ratios (HRs), 95% confidence intervals (CIs), and p-values derived from log-rank tests are shown where applicable.

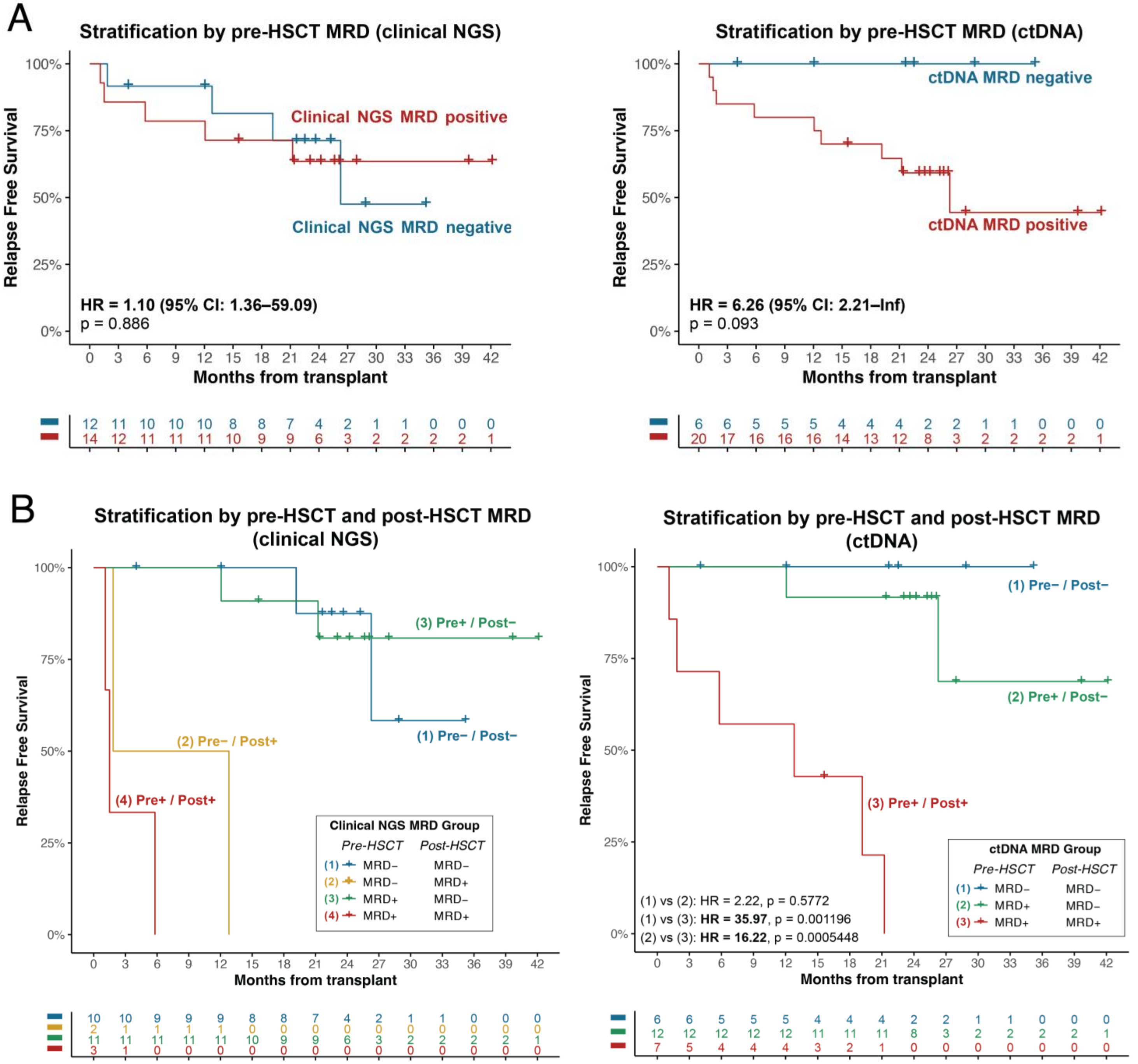

**Figure S16. AML-CAPP-Seq-based chimerism quantification.** Number of informative donor SNPs detected per patient using AML-CAPP-Seq.

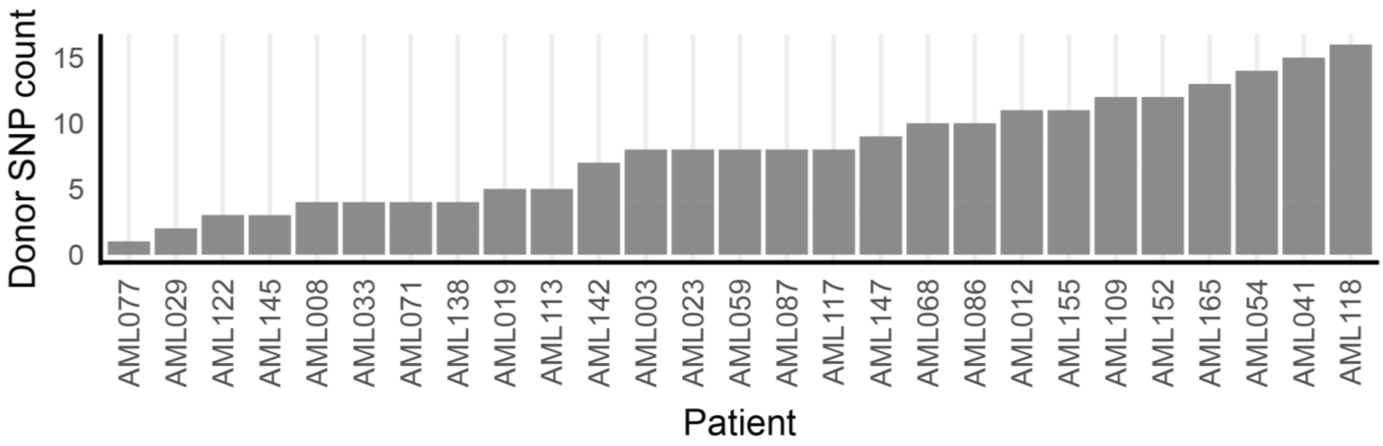

**Figure S17. AML-CAPP-Seq enables interrogation of clonal dynamics at relapse.** Schematic summarizing clonal evolution of canonical AML gene mutations between diagnosis and relapse grouped in 31 relapsed patients with paired samples, grouped by mutation class.

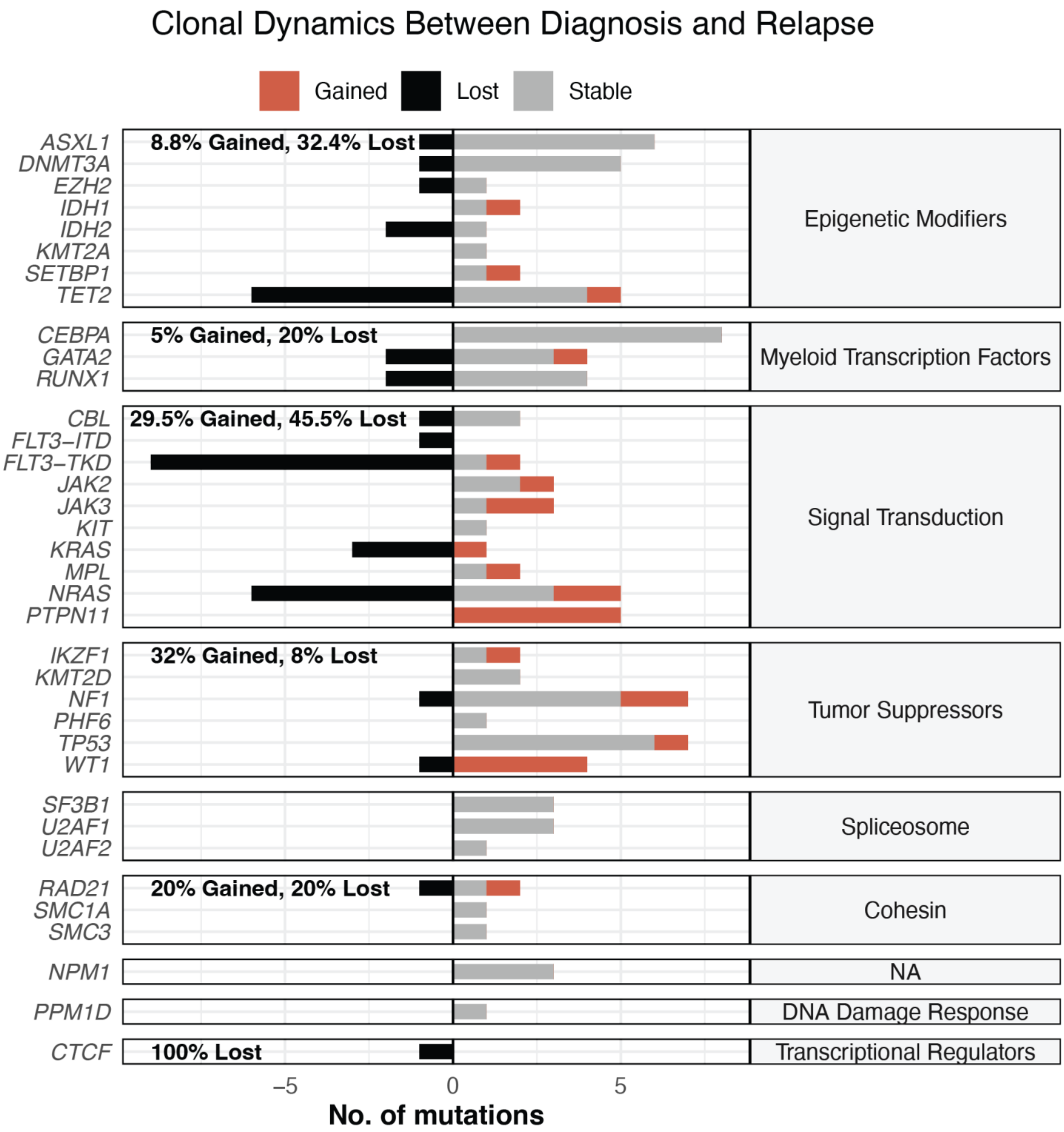
